# Preconception health among migrant women in England: a cross-sectional analysis of maternity services data 2018-2019

**DOI:** 10.1101/2023.01.26.23284338

**Authors:** Majel McGranahan, Elizabeth Augarde, Danielle Schoenaker, Helen Duncan, Sue Mann, Debra Bick, Felicity Boardman, Oyinlola Oyebode

**Affiliations:** University of Warwick Medical School, Medical School Building, Coventry CV4 7HL; Office for Health Improvement and Disparities (OHID), 39 Victoria Street, Westminster, London, SW1H 0EU; School of Primary Care, Population Sciences and Medical Education, Faculty of Medicine, University of Southampton, Tremona Road, SO16 6YD, Southampton, UK; NIHR Southampton Biomedical Research Centre, University of Southampton and University Hospital Southampton NHS Foundation Trust, Tremona Road, SO16 6YD, Southampton, UK; NHS England and Improvement, Wellington House, London SE1 8UG; Maternal Health Warwick Clinical Trials Unit, University of Warwick, Gibbet Hill Road, Coventry, CV4 7AL; Social Science as applied to Medicine University of Warwick Medical School, Medical School Building, Coventry CV4 7HL; Public Health and Lead, Centre for Public Health & Policy Wolfson Institute of Population Health, Barts and The London School of Medicine and Dentistry, Queen Mary University of London, EC1M 6BQ

**Keywords:** Prenatal care, Public health, Reproductive medicine

## Abstract

**Objective:** To examine inequalities in preconception health between migrant women in potentially vulnerable situations and non-migrant women.

**Design:** National cross-sectional study.

**Setting:** Data from the National Health Service (NHS) Maternity Services Data Set (MSDS) version 1.5, using data submitted by NHS maternity services in England.

**Participants:** All 652,880 women with an antenatal booking appointment between 1/4/2018 and 31/3/2019 were included. Data regarding migration status were available for 66.2% of the study population (n=432,022).

**Outcome measures:** Prevalence of preconception indicators were compared between probable migrants (those with complex social factors and English not their first language), possible migrants due to English not being a first language (without complex social factors), possible migrants due to complex social factors (who speak English as their first language) and unlikely migrants (those who speak English as their first language without complex social factors). Complex social factors include recent migrants, asylum seekers or refugees, difficulty reading or speaking English; alcohol and/or drugs misuse; all those aged under 20; and/or experiencing domestic abuse. Odds ratios were calculated comparing preconception indicators among those identified as migrants compared to unlikely migrants.

**Results:** Women identified as probable migrants (n=25,070) had over twice the odds of not taking folic acid before pregnancy and of having their first antenatal booking appointment after the recommended 10 weeks gestation compared to unlikely migrants (n=303,737), after adjusting for area-based deprivation level, mother’s age at booking, number of previous live births and ethnicity (odds ratio 2.15 (95% confidence interval 2.06 to 2.25) and 2.25 (2.18 to 2.32) respectively). Probable migrants had increased odds of previous obstetric complications and being underweight at booking, but lower odds of recorded physical and mental health conditions (apart from diabetes and hepatitis b), smoking and obesity in unadjusted and adjusted analyses.

**Conclusions:** Inequalities between migrant women in potentially vulnerable situations and non-migrants exist across many preconception indicators. Findings highlight the opportunity to improve preconception health in this population in order to reduce health inequalities and improve perinatal and neonatal outcomes.

**What is already known on this topic:**

- Nearly a third of live births in the UK are to migrant women (women born outside the UK).
- Compared with UK-born women, migrant women experience worse perinatal outcomes.
- The health of a woman before conception influences pregnancy outcomes, but little is known about inequalities between migrant and non-migrant women preconception.

**What this study adds:**

- This study showed that migrant women in potentially vulnerable situations are less likely to take folic acid before pregnancy, are more likely to be underweight, to have pre-existing diabetes or hepatitis b, and are more likely to have their first antenatal booking appointment after the recommended 10 weeks gestation, compared with non-migrants.
- Findings highlight the opportunity for more comprehensive preconception care for migrant women in potentially vulnerable situations.

## INTRODUCTION

Nearly a third (28.8%) of women giving birth in England and Wales in 2021 were migrants (born abroad themselves)^1^. Maternal mortality in the UK for women from certain countries is higher than for UK-born women; for example, women born in Bangladesh were over three times more likely to die during or after childbirth than UK-born women between 2018-2020^2^. Certain migrant groups have particularly complex social and health needs, as a result of poverty, trauma^3^, and financial and structural barriers to healthcare^4,5^, and migrant women are sometimes victims of forced labour, trafficking and sexual assault^6^.

Recent umbrella systematic reviews have highlighted the link between preconception risks and health behaviours with maternal and neonatal outcomes^7,8^. For example, high-quality evidence has shown being overweight or obese preconception increases the risk of miscarriage and adverse maternal and neonatal outcomes such as pre-eclampsia and stillbirth^7^, whilst maternal folic acid supplementation preconception is associated with fewer anomaly-related terminations and a reduction in neural tube defects^8^.

Migrant women are at particular risk of poor preconception and maternal health. Fear of deportation and charging prevents some women from accessing antenatal care^9,10^. In addition, refugees from certain countries have higher rates of tuberculosis, hepatitis B and HIV^11^, and may not be fully vaccinated^12^. A recent report by Doctors of the World (DOTW) found that approximately one in twenty (6%) migrant women accessing the London DOTW clinic were taking folic acid before pregnancy^10^, compared to one in four in the general population nationally (25.9%)^13^. Nearly half (45%) of migrant women did not have their first antenatal appointment until after 16 weeks^10^, in contrast to 10% of women in the general population^13^. National Institute for Health and Care Excellence (NICE) guidelines recommend that all pregnant women are seen for their first antenatal appointment within the first ten weeks of pregnancy^14^. A UK rapid review of barriers and facilitators to preconception care identified limited evidence among migrant women, with almost all studies undertaken in non-migrant populations^15^. Despite the potential for higher preconception risks among migrant women, and known links between preconception risks and pregnancy outcomes, no previous studies have compared migrant and non-migrant women’s preconception health in the UK. It is therefore not known where the biggest preconception health inequalities lie (if any) and where policy or health interventions could be targeted.

Our objective was to examine inequalities in preconception health between migrant women in potentially vulnerable situations and non-migrant women. For the purposes of this paper, migrant women in potentially vulnerable situations include asylum seekers and refugees^16^, recent migrants and those with difficulty speaking or reading English^17^.

*\*The terms women/woman are used throughout this manuscript, but it is recognised that not all pregnant people or those who give birth identify as women*.

## METHODS

### Setting

This national cross-sectional study used data from the National Health Service (NHS) Maternity Services Data Set (MSDS) version 1.5. The MSDS re-uses operational and clinical data, and is used for examining health inequalities, monitoring health outcomes, commissioning and planning of services^18^. All NHS-funded maternity units in England are required to submit their data to MSDS, and data are available at the patient-level: from a woman’s first antenatal appointment to discharge from maternity services^18^. Some data items within MSDS are mandatory and must be reported, some are required, and some are optional. For this study, only data collected at the first antenatal appointment (the ‘booking appointment’) were accessed. Women who had their booking appointment during the period 1 April 2018 to 31 March 2019 were included.

As per the Health and Social Care Act 2012, consent from participants was not required. Data were accessed through the Office for Health Improvement and Disparities (OHID). Data were anonymised and provided to OHID by NHS Digital, for the purposes of population health surveillance, with a relevant data sharing agreement.

### Participants

For this study, a variable called “complex social factors” was used to help identify probable migrants. The MSDS complex social factors variable is based on NICE Guidance CG110 and includes women who are recent migrants, asylum seekers or refugees, or have difficulty reading or speaking English; women who misuse alcohol and/or drugs; all women aged under 20; and/or women who experience domestic abuse^17^. It was not possible to identify the reason that a woman was assigned ‘complex social factors’ from the data available. This was used alongside the variable describing whether women speak English as their first language to allow preconception and maternal health indicators to be compared between:

1. Women identified as probable migrants (those with complex social factors who do not speak English as their first language)
2. Women identified as possible migrants because they do not speak English as their first language (but who do not have complex social factors)
3. Women identified as possible migrants due to having complex social factors (but who speak English as their first language)
4. Women identified as unlikely migrants (women without complex social factors who speak English as their first language).

By combining information on women for whom English is not a first language with complex social factors, the exposure group is likely to be a group of migrant women in potentially vulnerable situations: either asylum seekers or refugees^16^, recent migrants, or those who have difficulty reading or speaking English^17^. Uncertainty due to the method of identifying migrant women is a risk (because some women in this group may not be migrants); however, the majority (99%) of UK-born people speak English as a first language at home and fewer than 0.1% of UK-born individuals have difficulty speaking English^19^.

### Variables collected

Preconception indicators identified from a previous review and report card of priority preconception indicators for national surveillance using MSDS were included^20,21^ (Table 2). We also included data regarding late antenatal booking (after 10, 16 and 20 weeks), because late booking has been identified as an issue for migrant women in previous studies^10,22,23^, may reflect engagement with services preconception, and booking by 10 weeks gestation is recommended by NICE guidelines^14^. Data regarding alcohol consumption and substance use were included because they were considered relevant to the classification of migration status (because they would also lead to a woman being flagged as having complex social factors), although they were not included in the previous study mentioned above due to poor data quality^20^. Data regarding mother’s age at booking (years), ethnicity, number of previous live births and Index of Multiple Deprivation 2015 (IMD) were also obtained, in order to be able to address possible confounding.

Most data available from the booking antenatal appointment within MSDS are obtained retrospectively, including self-reported health behaviours such as folic acid use, alcohol intake and substance use, as well as medical, family, and obstetric history. Where possible, during the first face-to-face antenatal appointment, height and weight are measured by a midwife or other healthcare professional, and Body Mass Index (BMI) calculated^14^. IMD relates to the lower super output area (LSOA) of residence which is determined from a woman’s postcode.

### Variable exclusions due to spurious results

Some variables within MSDS record previous pregnancy and delivery events. For the previous caesarean sections variable, records were recoded as ‘missing’ if more than 10 caesarean sections were reported (based on discussion with experts). For the previous live births variable, records were recoded as ‘missing’ if more than 25 live births were reported (based on 38 years of fertility with one baby born per 18 months). BMI was recoded as ‘missing’ if it was reported as above 80 or below 13, or if the booking appointment was after 14 weeks (in line with OHID methodology for national Child and Maternal Health statistics^24^). For preconception indicators related to previous obstetric conditions, only women with at least one previous pregnancy were included in analyses.

### Statistical analysis

Descriptive statistics were used to describe women’s age at booking (years), number of previous live births, ethnicity, and deprivation decile (IMD) across the whole study population, and by migration category. The prevalence of each preconception indicator was calculated, together with 95% confidence intervals for the whole study population, and by migration category.

Odds ratios and 95% confidence intervals were calculated using binary logistic regression for each preconception indicator according to migration status, with unlikely migration status as the reference category. Age, ethnicity and IMD were not included as preconception indicator outcome variables in logistic regression analyses but were included as covariates in multivariable models. ‘Complex social factors’ was not included as a preconception indicator outcome variable because it contributed to migration status classification. Age (years), ethnicity (categorised as white, mixed, Asian, black, other or ‘missing’ ethnicity), number of previous live births, and most deprived IMD decile (as a binary variable) were included as covariates in multivariable models. Ethnicity was included because ethnicity is known to impact maternal outcomes^2,25^ and may have a relationship with migration status^26^; mother’s age and deprivation are also linked to maternal outcomes^2^ and may be associated with migration status. Number of previous live births were included within models for previous obstetric complications (because more live births results in more opportunity for previous obstetric complications), health behaviours (because they may have received more pregnancy-related health behaviour advice through previous pregnancies), underweight, overweight and obesity (because weight gain frequently occurs between pregnancies^27^) and late antenatal booking (because women with more previous live births may be more aware of the antenatal care system and the importance of booking by 10 weeks gestation, or may have childcare commitments that preclude early booking). Pre-existing physical and mental conditions and social support were not considered to be confounded by the number of previous live births.

Five logistic regression models were calculated for each preconception indicator. Forced entry model fitting was used. **Model 1**, a univariate model, included migration status as the exposure variable and the relevant preconception indicator as the outcome variable. Participants with missing data for complex social factors and/or English as a first language were categorised as ‘missing’ migration status and included in the model. **Model 2a** adjusted for most deprived IMD decile, mother’s age at booking (years) and ethnicity. **Model 2b** additionally adjusted for number of previous live births.

Sensitivity analyses were conducted for each preconception indicator (Models 3a/b, 4a/b and 5a/b). To examine the contribution of adjustment for ethnicity on the findings, **Model 3a** adjusted for most deprived IMD decile and mother’s age at booking (years) but not ethnicity, and **Model 3b** additionally adjusted for number of previous live births. To examine the effect of excluding individuals with missing data, **Model 4a** excluded participants with missing data for the migration status variable and/or the ethnicity variable (because these variables had a reasonable amount of missing data at 33.8% and 15.8% respectively), and **Model 4b** additionally adjusted for number of previous live births. To assess potential bias due to misclassification of probable or possible migrant due to age or history of substance abuse resulting in complex social factors being recorded, **Models 5a** and **5b** replicated Models 2a and 2b respectively, but excluded those aged under 20 years and/or those with a recorded history of substance use.

Data management and analysis were undertaken using R version 4.1.3.

### Ethical approval

Ethical approval was obtained from the Biomedical and Scientific Research Ethics Committee of the University of Warwick Research Governance and Ethics Committee (Reference BSREC 158/21-22).

### Patient and public involvement

A community advisory group, including migrant women, healthcare professionals and other stakeholders commented on the research plan but were unable to contribute to interpretation of the initial results because of data sharing agreements.

## RESULTS

### Participants

Data regarding migration status (both English as a first language and complex social factors) were available for 66.2% of the study population (432,022 women), of whom 5.8% (25,070 women) were identified as probable migrants, 15.5% (66,783 women) as possible migrants due to English not being their first language (but without complex social factors), 8.4% (36,433 women) as possible migrants due to having complex social factors recorded (but speaking English as their first language), and 70.3% (303,737 women) as unlikely migrants (Table 1).

**Table 1.** Participant Characteristics. Complex social factors include women who misuse alcohol and/or drugs; women who are recent migrants, asylum seekers or refugees, or have difficulty reading or speaking English; all women aged under 20; and/or women who experience domestic abuse.

| Characteristic | Overall<br>N = 652,880 | Probable migrant<br>(English not their first<br>language and complex<br>social factors)<br>N = 25,070 | Possible migrant (only<br>English not their first<br>language)<br>N = 66,783 | Possible migrant (only<br>complex social<br>factors)<br>N = 36,433 | Unlikely migrant<br>(English as first<br>language without<br>complex social<br>factors)<br>N = 303,737 | Missing migration<br>status (first language<br>and/or complex social<br>factor data missing)<br>N = 220,858 |
| --- | --- | --- | --- | --- | --- | --- |
| **Mother's age at booking (years)** |  |  |  |  |  |  |
| Mean (SD) | 29.8 (5.7) | 28.1 (6.4) | 30.9 (5.2) | 24.4 (6.9) | 30.4 (5.3) | 29.8 (5.6) |
| Median (IQR) | 30 (26 to 24) | 28 (23 to 32) | 31 (27 to 34) | 22 (19 to 30) | 30 (27 to 34) | 30 (26 to 34) |
| Missing, n | 9 | 0 | 0 | 0 | 0 | 9 |
| **Number of previous live births** |  |  |  |  |  |  |
| Mean (SD) | 0.9 (1.1) | 1.0 (1.3) | 0.9 (1.1) | 0.8 (1.3) | 0.9 (1.1) | 0.9 (1.2) |
| Median (IQR) | 1 (0 to 1) | 1 (0 to 1) | 1 (0 to 1) | 0 (0 to 1) | 1 (0 to 1) | 1 (0 to 1) |
| Missing, n | 128,308 | 1,733 | 4,293 | 4,757 | 30,311 | 87,214 |
| **Ethnicity of mother, n (%)** |  |  |  |  |  |  |
| White | 424,453 (77.2) | 8,944 (45.0) | 28,261 (52.0) | 27,656 (85.2) | 219,683 (82.4) | 139,909 (79.4) |
| Mixed | 11,695 (2.1) | 438 (2.2) | 1,094 (2.0) | 848 (2.6) | 5,547 (2.1) | 3,768 (2.1) |
| Asian | 66,444 (12.1) | 5,638 (28.4) | 16,299 (30.0) | 1,685 (5.2) | 23,932 (9.0) | 18,890 (10.7) |
| Black | 26,172 (4.8) | 1,876 (9.4) | 3,691 (6.8) | 1,514 (4.7) | 11,100 (4.2) | 7,991 (4.5) |
| Other | 20,788 (3.8) | 2,987 (15.0) | 4,994 (9.2) | 776 (2.4) | 6,404 (2.4) | 5,627 (3.2) |
| Missing, n | 103,328 | 5,187 | 12,443 | 3,954 | 37,071 | 44,673 |
| **Index of Multiple Deprivation 2015 (IMD), n (%)** |  |  |  |  |  |  |
| IMD 1 | 92,528 (14.2) | 7,130 (28.4) | 11,408 (17.1) | 8,280 (22.7) | 31,389 (10.3) | 34,321 (15.5) |
| IMD 2 | 82,397 (12.6) | 5,011 (20.0) | 9,876 (14.8) | 5,834 (16.0) | 33,844 (11.1) | 27,832 (12.6) |
| IMD 3 | 77,043 (11.8) | 3,900 (15.6) | 9,402 (14.1) | 4,975 (13.7) | 33,866 (11.1) | 24,900 (11.3) |
| IMD 4 | 70,7697 (10.8) | 2,705 (10.8) | 8,046 (12.0) | 3,995 (11.0) | 33,484 (11.0) | 20,689 (10.2) |
| IMD 5 | 63,917 (9.8) | 2,026 (8.1) | 6,672 (10.0) | 3,255 (8.9) | 31,275 (10.3) | 20,689 (9.4) |
| IMD 6 | 60,930 (9.3) | 1,561 (6.2) | 6,038 (9.0) | 2,686 (7.4) | 31,238 (10.3) | 19,407 (8.8) |
| IMD 7 | 56,858 (8.7) | 1,089 (4.3) | 4,854 (7.3) | 2,442 (6.7) | 29,558 (9.7) | 18,917 (8.6) |
| IMD 8 | 54,125 (8.3) | 786 (3.1) | 4,262 (6.4) | 2,035 (5.6) | 28,391 (9.3) | 18,651 (8.4) |
| IMD 9 | 50,586 (7.7) | 564 (2.2) | 3,588 (5.4) | 1,788 (4.9) | 27,267 (9.0) | 17,379 (7.9) |
| IMD 10 | 43,799 (6.7) | 298 (1.2) | 2,638 (4.0) | 1,143 (3.1) | 23,425 (7.7) | 16,295 (7.4) |
| Missing, n | 0 | 0 | 0 | 0 | 0 | 0 |

### Descriptive data

As shown in Table 1, the mean age for study population overall was 29.8 (standard deviation (SD) 5.7), ranging from 24.4 for possible migrants (only complex social factors) to 30.9 for possible migrants (only English not their first language). The mean number of previous live births was 0.9 (SD 1.1), with the mean ranging from 0.8 (among possible migrants due to only having complex social factors and those with missing migration status) to 1.0 (among probable migrants). The majority (77.8%) of the study population overall were of white ethnicity, 12.8% of Asian ethnicity, 4.8% black ethnicity and 2.1% mixed ethnicity, whereas in the probable migrant group, 45.0% were of white ethnicity, 28.4% of Asian ethnicity, 9.4% of black ethnicity and 2.2% mixed ethnicity. There was a higher proportion of individuals in the lowest IMD decile in the probable migrant group (28.4%) compared with overall (14.2%), unlikely migrant (10.3%) and possible migrant due to English not being their first language (17.1%) or complex social factors (22.7%).

### Preconception indicators

Table 2 shows unadjusted numbers and proportions of participants with each preconception indicator according to migration category. There were proportionally more participants with known previous obstetric complications in the probable migrant group (34.4%, 95% Confidence Interval (CI) 33.6-35.2) compared to unlikely migrants (26.9%, 95% CI 26.7-27.1). A higher proportion of participants in the probable migrant group did not take folic acid supplementation before pregnancy (87.6%, 95% CI 87.2-88.1) compared to all other groups. Proportionally fewer participants in the probable migrant group smoked around conception (16.5%, 95% CI 16.0-16.9) compared to other groups apart from possible migrant due to English not being their first language (13.9%, 95% CI 13.7-14.2). A higher proportion of participants in the probable migrant group were underweight (BMI<18.5) than other groups (5.2%, 95% CI 4.9-5.6), similar proportions were overweight (27.8%, 95% CI 27.1-28.5) in the probable migrant group compared with the overall cohort (27.8%, 95% CI 27), and fewer were obese (17.2%, 95% CI 16.7-17.8) in the probable migrant group compared with 27.8% (95% CI 27.7-28.0) overall and (27.9%, 95% CI 27.7-28.1) in the unlikely migrant group). A smaller proportion of probable migrants had a diagnosed mental or physical health condition, although more had diabetes (1.6%, 95% CI 1.5-1.8 compared with 1.1%, 95% CI 1.1-1.2 in the unlikely migrant group) and hepatitis b (0.7%, 95% CI 0.6-0.8 compared to 0.1%, 95% CI 0.1-0.1). Late antenatal booking (after 10, 16 and 20 weeks gestation) occurred more often in the probable migrant group compared with all other groups.

**Table 2:** Preconception indicators by migration status. Complex social factors include women who misuse alcohol and/or drugs; women who are recent migrants, asylum seekers or refugees, or have difficulty reading or speaking English; all women aged under 20; and/or women who experience domestic abuse. Probable migrant: English not their first language and complex social factors. Possible migrant (EAFL): only English not their first language. Possible migrant (CSF): only complex social factors. Unlikely migrant: English as first language without complex social factors. Missing migration status: first language and/or complex social factor data missing.

| Preconception indicator | Overall<br>N = 652,880 | Probable migrant<br>N = 25,070 | Possible migrant (EAFL)<br>N = 66,783 | Possible migrant (CSF)<br>N = 36,433 | Unlikely migrant<br>N = 303,737 | Missing migrant status<br>N = 220,858 |
| --- | --- | --- | --- | --- | --- | --- |
|  | n<br>% (95% CI) | n<br>% (95% CI) | n<br>% (95% CI) | n<br>% (95% CI) | n<br>% (95% CI) | n<br>% (95% CI) |
| **Ethnic minority (n=549,552)** | 549,552 | 10,939 | 26,078 | 4,823 | 46,983.0 | 36,276 |
|  | 22.8 (22.7-22.9) | 55.0 (54.3-55.7) | 48.0 (47.6-48.4) | 14.8 (14.5-15.2) | 17.6 (17.5-17.8) | 20.6 (20.4-20.8) |
| **Unemployed and seeking work (n=472,181)** | 26,849 | 2,295 | 2,770 | 5,601 | 11,897 | 4,286 |
|  | 5.7 (5.6-5.8) | 10.6 (10.2-11.0) | 4.5 (4.4-4.7%) | 19.0 (18.6-19.5) | 4.4 (4.3-4.5) | 4.8 (4.6-4.9) |
| **Living in most deprived area (bottom 10%)**<br>**(n=652,880)** | 92,528 | 7,130 | 11,408 | 8,280 | 31,389 | 34,321 |
|  | 14.2 (14.1-14.3) | 28.4 (27.9-29.0) | 17.1 (16.8-17.4) | 22.7 (22.3-23.2) | 10.3 (10.2-10.4) | 15.5 (15.4-15.7) |
| **No adequate support available during and**<br>**after pregnancy (n=449,884)** | 26,590 | 686 | 3,027 | 1,463 | 9,191 | 12,223 |
|  | 5.9 (5.8-6.0) | 3.2 (3.0-3.5) | 5.2 (5.0-5.4) | 4.9 (4.6-5.1) | 3.6 (3.5-3.7) | 14.3 (14.1-14.5) |
| **Advanced maternal age (greater than or**<br>**equal to 35 years) (n=652,871)** | 139,661 | 4,181 | 16,501 | 3,713 | 69,901 | 45,365 |
|  | 21.4 (21.3-21.5) | 16.7 (16.2-17.1) | 24.7 (24.4-25.0) | 10.2 (9.9-10.5) | 23.0 (22.9-23.2) | 20.5 (20.4-20.7) |
| **Teenage pregnancy (less than 20 years)**<br>**(n=652,871)** | 24,675 | 2,820 | 0 | 15,612 | 0 | 6,243 |
|  | 3.8 (3.7-3.8) | 11.2 (10.9-11.6) | 0.0 (0.0-0.0) | 42.9 (42.3-43.4) | 0.0 (0.0-0.0) | 2.8 (2.8-2.9) |
| **Known previous obstetric complication**<br>**(n=329,228)** | 80,694 | 4,722 | 13,136 | 4,136 | 46,885 | 11,815 |
|  | 24.5 (24.4-24.7) | 34.4 (33.6-35.2) | 32.9 (32.5-33.4) | 24.3 (23.7-25.0) | 26.9 (26.7-27.1) | 14.0 (13.7-14.2) |
| **Previous pre-eclampsia, eclampsia,**<br>**gestational proteinuria, HELLP (n=329,228)** | 3,469 | 127 | 458 | 141 | 2,054 | 689 |
|  | 1.1 (1.0-1.1) | 0.9 (0.8-1.1) | 1.1 (1.0-1.3) | 0.8 (0.7-1.0) | 1.2 (1.1-1.2) | 0.8 (0.8-0.9) |
| **Previous gestational hypertension**<br>**(n=329,228)** | 5,208 | 255 | 662 | 339 | 3,170 | 782 |
|  | 1.6 (1.5-1.6) | 1.9 (1.6-2.1) | 1.7 (1.5-1.8) | 2.0 (1.8-2.2) | 1.8 (1.8-1.9) | 0.9 (0.9-1.0) |
| **Previous gestational diabetes mellitus**<br>**(n=329,228)** | 7,506 | 500 | 1,735 | 243 | 3,918 | 1,110 |
|  | 2.3 (2.2-2.3) | 3.6 (3.3-4.0) | 4.3 (4.2-4.6) | 1.4 (1.3-1.6) | 2.3 (2.2-2.3) | 1.3 (1.2-1.4) |
| **Previous caesarean section (n=306,429)** | 69,989 | 3,542 | 10,132 | 3,226 | 37,446 | 15,643 |
|  | 22.8 (22.7-23.0) | 26.3 (25.5-27.0) | 25.9 (25.5-26.3) | 19.9 (19.3-20.5) | 22.5 (22.3-22.7) | 22.1 (21.8-22.4) |
| **Previous pregnancy loss (n=301,168)** | 117,258 | 4,389 | 11,941 | 7,459 | 60,274 | 33,195 |
|  | 38.9 (38.8-39.1) | 34.8 (33.9-35.6) | 31.6 (31.2-32.1) | 48.8 (48.0-49.6) | 36.9 (36.7-37.1) | 46.0 (45.6-46.3) |
| **Previous live birth (n=524,572)** | 288,073 | 12,293 | 35,677 | 14,045 | 152,511 | 73,547 |
|  | 54.9 (54.8-55.1) | 52.7 (52.0-52.3) | 57.1 (56.7-57.5) | 44.3 (43.8-44.9) | 55.8 (55.6-56.0) | 55.0 (54.8-55.3) |
| **Not taken folic acid supplementation before**<br>**pregnancy (488,987)** | 355,648 | 19,673 | 46,049 | 27,737 | 186,997 | 75,192 |
|  | 72.7 (72.6-72.9) | 87.6 (87.2-88.1) | 75.1 (74.8-75.4) | 84.8 (84.4-85.1) | 70.0 (69.8-70.2) | 71.3 (71.0-71.6) |
| **Smoking around conception (n=604,514)** | 117,602 | 4,068 | 9,051 | 14,538 | 53,559 | 36,386 |
|  | 19.5 (19.5-19.7) | 16.5 (16.0-16.9) | 13.9 (13.7-14.2) | 40.9 (40.4-41.4) | 18.1 (18.0-18.3) | 19.8 (19.6-20.0) |
| **Smokers who did not quit smoking during**<br>**year before pregnancy (n=138,422)** | 117,602 | 4,068 | 9,051 | 14,538 | 53,559 | 36,386 |
|  | 85.0 (84.9-85.2) | 88.3 (87.3-89.2) | 82.2 (81.5-82.9) | 89.7 (89.2-90.1) | 82.2 (81.9-82.5) | 87.8 (87.5-88.1) |
| **Underweight at booking (BMI less than 18.5**<br>**kg per m2) (n=496,267)** | 15,345 | 871 | 1,929 | 1,846 | 6,792 | 3,907 |
|  | 3.1 (3.0-3.1) | 5.2 (4.9-5.6) | 3.5 (3.3-3.6) | 6.4 (6.2-6.7) | 2.6 (2.5-2.7) | 2.9 (2.8-3.0) |
| **Overweight at booking (BMI 25 to 29.9 kg**<br>**per m2) (n=496,267)** | 138,155 | 4,622 | 15,602 | 7,204 | 72,797 | 37,930 |
|  | 27.8 (27.7-28.0) | 27.8 (27.1-28.5) | 28.2 (27.9-28.6) | 25.1 (24.6-25.7) | 27.9 (27.7-28.1) | 28.1 (27.9-28.3) |
| **Obesity at booking (BMI greater than or** | 110,596 | 2,866 | 9,537 | 6,654 | 59,884 | 31,655 |
| equal to 30 kg per m2) (n=496,267) | 22.3 (22.2-22.4) | 17.2 (16.7-17.8) | 17.3 (16.9-17.6) | 23.2 (22.7-23.7) | 23.0 (22.8-23.1) | 23.5 (23.2-23.7) |
| Substance use at booking (n=526,446) | 6,143 | 110 | 65 | 1,405 | 957 | 3,606 |
|  | 1.2 (1.1-1.2) | 0.5 (0.4-0.6) | 0.1 (0.1-0.1) | 4.1 (3.9-4.3) | 0.3 (0.3-0.4) | 3.1 (3.0-3.2) |
| Alcohol drinking at booking (greater than or equal to 1 unit per week) (n=398,959) | 10,400 | 170 | 527 | 826 | 4,064 | 4,813 |
|  | 2.6 (2.6-2.7) | 1.1 (0.9-1.3) | 1.1 (1.0-1.2) | 3.4 (3.2-3.6) | 1.8 (1.8-1.9) | 5.5 (5.4-5.7) |
| At least one mental or physical health condition (n=652,880) | 158,839 | 4,424 | 17,361 | 11,761 | 99,456 | 25,837 |
|  | 24.3 (24.2-24.4) | 17.6 (17.2-18.1) | 26.0 (25.7-26.3) | 32.3 (31.8-32.8) | 32.7 (32.6-32.9) | 11.7 (11.6-11.8) |
| Mental health condition (n=652,880) | 60,973 | 1,079 | 3,890 | 7,244 | 38,831 | 9,929 |
|  | 9.3 (9.3-9.4) | 4.3 (4.1-4.6) | 5.8 (5.6-6.0) | 19.9 (19.5-20.3) | 12.8 (12.7-12.9) | 4.5 (4.4-4.6) |
| Physical health condition (n=652,880) | 124,705 | 3,743 | 15,289 | 7,064 | 78,817 | 19,792 |
|  | 19.1 (19.0-19.2) | 14.9 (14.5-15.4) | 22.9 (22.6-23.2) | 19.4 (19.0-19.8) | 25.9 (25.8-26.1) | 9.0 (8.8-9.1) |
| Diabetes (n=652,880) | 6,343 | 411 | 1,073 | 383 | 3,434 | 1,042 |
|  | 1.0 (0.9-1.0) | 1.6 (1.5-1.8) | 1.6 (1.5-1.7) | 1.1 (1.0-1.2) | 1.1 (1.1-1.2) | 0.5 (0.4-0.5) |
| Hypertension (n=652,880) | 6,696 | 216 | 850 | 326 | 4,358 | 946 |
|  | 1.0 (1.0-1.1) | 0.9 (0.8-1.0) | 1.3 (1.2-1.4) | 0.9 (0.8-1.0) | 1.4 (1.4-1.5) | 0.4 (0.4-0.5) |
| Cardiac disease (n=652,880) | 5,184 | 132 | 579 | 323 | 3,201 | 949 |
|  | 0.8 (0.8-0.8) | 0.5 (0.4-0.6) | 0.9 (0.8-0.9) | 0.9 (0.8-1.0) | 1.1 (1.0-1.1) | 0.4 (0.4-0.5) |
| Thromboembolic condition (n=652,880) | 3,885 | 120 | 519 | 205 | 2,473 | 568 |
|  | 0.6 (0.6-0.6) | 0.5 (0.4-0.6) | 0.8 (0.7-0.8) | 0.6 (0.5-0.6) | 0.8 (0.8-0.8) | 0.3 (0.2-0.3) |
| Renal disease (n=652,880) | 5,126 | 183 | 625 | 291 | 2,791 | 1,236 |
|  | 0.8 (0.8-0.8) | 0.7 (0.6-0.8) | 0.9 (0.9-1.0) | 0.8 (0.7-0.9) | 0.9 (0.9-1.0) | 0.6 (0.5-0.6) |
| Hepatitis B (n=652,880) | 998 | 177 | 376 | 35 | 292 | 118 |
|  | 0.2 (0.1-0.2) | 0.7 (0.6-0.8) | 0.6 (0.5-0.6) | 0.1 (0.1-0.1) | 0.1 (0.1-0.1) | 0.1 (0.1-0.1) |
| Cancer (n=652,880) | 1,085 | 20 | 102 | 52 | 742 | 169 |
|  | 0.2 (0.2-0.2) | 0.1 (0.1-0.1) | 0.2 (0.1-0.2) | 0.1 (0.1-0.2) | 0.2 (0.2-0.3) | 0.1 (0.1-0.1) |
| Inherited conditions (n=652,880) | 13,323 | 228 | 1,192 | 1,040 | 7,592 | 3,271 |
|  | 2.0 (2.0-2.1) | 0.9 (0.8-1.0) | 1.8 (1.7-1.9) | 2.9 (2.7-3.0) | 2.5 (2.4-2.6) | 1.5 (1.4-1.5) |
| Family history of diabetes (n=652,880) | 134,398 | 7,124 | 20,513 | 9,213 | 78,802 | 18,746 |
|  | 20.6 (20.5-20.7) | 28.4 (27.9-29.0) | 30.7 (30.4-31.1) | 25.3 (24.8-25.7) | 25.9 (25.8-26.1) | 8.5 (8.4-8.6) |
| Booking after 10 weeks gestation (n=652,309) | 275,191 | 15,665 | 30,676 | 17,282 | 117,063 | 94,505 |
|  | 42.2 (42.1-42.3) | 62.5 (61.9-63.1) | 46.0 (45.6-46.3) | 47.5 (47.0-48.0) | 38.6 (38.4-38.7) | 42.9 (42.6-43.1) |
| Booking after 16 weeks gestation (n=652,309) | 52,516 | 5,483 | 5,506 | 4,332 | 15,970 | 21,225 |
|  | 8.1 (8.0-8.1) | 21.9 (21.4-22.4) | 8.2 (8.0-8.5) | 11.9 (11.6-12.2) | 5.3 (5.2-5.3) | 9.6 (9.5-9.7) |
| Booking after 20 weeks gestation (n=652,309) | 36,459 | 3,630 | 3,320 | 2,893 | 10,590 | 16,026 |
|  | 5.6 (5.5-5.6) | 14.5 (14.1-14.9) | 5.0 (4.8-5.1) | 7.9 (7.7-8.2) | 3.5 (3.4-3.6) | 7.3 (7.2-7.4) |

### Logistic regression models

Table 3 shows the unadjusted and fully adjusted logistic regression models (Models 1 and 2a/b respectively). Logistic regression models including sensitivity analyses (Models 3a/b-5a/b) and odds ratios for all covariates are included in Supplementary File 1.

**Table 3:** Logistic regression models for preconception indicators. Complex social factors include women who misuse alcohol and/or drugs; women who are recent migrants, asylum seekers or refugees, or have difficulty reading or speaking English; all women aged under 20; and/or women who experience domestic abuse. Unlikely migrant: English as first language without complex social factors. Probable migrant: English not their first language and complex social factors. Possible migrant (EAFL): only English not their first language. Possible migrant (CSF): only complex social factors. Missing migration status: first language and/or complex social factor data missing. Model 1 is unadjusted. Model 2a adjusts for mother’s age at booking (years), most deprived Index of Multiple Deprivation (IMD) decile and ethnicity; Model 2b additionally adjusts for number of previous live births.

| Indicator | Model | Unlikely migrant | Probable migrant Odds ratio (95% confidence interval) | Possible migrant (EAFL) Odds ratio (95% confidence interval) | Possible migrant (CSF) Odds ratio (95% confidence interval) | Missing migrant status Odds ratio (95% confidence interval) |
| --- | --- | --- | --- | --- | --- | --- |
| **Unemployed and seeking work** | Model 1 (n=472,181) | Reference | 2.58 (2.46-2.70) | 1.03 (0.98-1.07) | 5.10 (4.92-5.28) | 1.09 (1.05-1.13) |
|  | Model 2a (n=472,175) | Reference | 1.74 (1.66-1.83) | 0.96 (0.92-1.01) | 2.84 (2.73-2.95) | 1.03 (0.99-1.07) |
| **No adequate support available during and after pregnancy** | Model 1 (n=449,884) | Reference | 0.89 (0.82-0.96) | 1.47 (1.41-1.53) | 1.36 (1.29-1.44) | 4.46 (4.33-4.59) |
|  | Model 2a (n=449,877) | Reference | 1.10 (1.02-1.19) | 1.75 (1.67-1.82) | 1.37 (1.29-1.45) | 4.56 (4.43-4.69) |
| **Known previous obstetric complication** | Model 1 (n=329,228) | Reference | 1.42 (1.37-1.47) | 1.33 (1.30-1.36) | 0.87 (0.84-0.90) | 0.44 (0.43-0.45) |
|  | Model 2b (n=325,619) | Reference | 1.26 (1.21-1.31) | 1.18 (1.15-1.21) | 0.91 (0.88-0.95) | 0.43 (0.42-0.44) |
| **Previous pre-eclampsia, eclampsia, gestational proteinuria, HELLP** | Model 1 (n=329,228) | Reference | 0.78 (0.65-0.93) | 0.97 (0.88-1.08) | 0.70 (0.59-0.83) | 0.69 (0.63-0.75) |
|  | Model 2b (n=325,619) | Reference | 0.69 (0.57-0.83) | 0.94 (0.84-1.04) | 0.64 (0.54-0.76) | 0.67 (0.62-0.73) |
| **Previous gestational hypertension** | Model 1 (n=329,228) | Reference | 1.02 (0.89-1.16) | 0.91 (0.83-0.99) | 1.10 (0.98-1.22) | 0.50 (0.47-0.54) |
|  | Model 2b (n=325,619) | Reference | 0.97 (0.85-1.11) | 0.89 (0.81-0.97) | 1.07 (0.95-1.20) | 0.50 (0.47-0.55) |
| **Previous gestational diabetes mellitus** | Model 1 (n=329,228) | Reference | 1.64 (1.49-1.80) | 1.97 (1.86-2.09) | 0.63 (0.55-0.71) | 0.58 (0.54-0.62) |
|  | Model 2b (n=325,619) | Reference | 1.21 (1.01-1.24) | 1.36 (1.28-1.45) | 0.68 (0.59-0.77) | 0.55 (0.51-0.59) |
| **Previous caesarean section** | Model 1 (n=306,429) | Reference | 1.23 (1.18-1.28) | 1.21 (1.18-1.24) | 0.86 (0.82-0.89) | 0.98 (0.96-1.00) |
|  | Model 2b (n=305,932) | Reference | 1.13 (1.08-1.18) | 1.07 (1.04-1.10) | 1.02 (0.98-1.07) | 0.97 (0.95-0.99) |
| **Previous loss** | Model 1 (n=301,168) | Reference | 0.91 (0.88-0.95) | 0.79 (0.77-0.81) | 1.63 (1.58-1.69) | 1.45 (1.43-1.48) |
|  | Model 2b (n=297,559) | Reference | 1.06 (1.02-1.11) | 0.85 (0.83-0.87) | 1.78 (1.72-1.86) | 1.45 (1.42-1.47) |
| **Not taken folic acid supplementation before pregnancy** | Model 1 (n=488,987) | Reference | 3.04 (2.92-3.16) | 1.29 (1.27-1.32) | 2.38 (2.31-2.46) | 1.06 (1.05-1.08) |
|  | Model 2b (n=431,990) | Reference | 2.15 (2.06-2.25) | 1.22 (1.19-1.25) | 1.49 (1.44-1.55) | 1.09 (1.07-1.11) |
| **Smoking around conception** | Model 1 (n=604,514) | Reference | 0.89 (0.86-0.92) | 0.73 (0.71-0.75) | 3.12 (3.05-3.20) | 1.11 (1.10-1.13) |
|  | Model 2b (n=526,809) | Reference | 0.86 (0.82-0.90) | 1.03 (1.00-1.06) | 1.80 (1.74-1.87) | 1.05 (1.03-1.07) |
| **Smokers who did not quit smoking during year before pregnancy** | Model 1 (n=138,422) | Reference | 1.63 (1.49-1.79) | 1.00 (0.95-1.05) | 1.88 (1.78-1.99) | 1.56 (1.50-1.61) |
|  | Model 2b (n=121,676) | Reference | 1.56 (1.41-1.73) | 1.21 (1.15-1.28) | 1.33 (1.23-1.44) | 1.32 (1.27-1.38) |
| **Underweight at booking (BMI less than 18.5 kg per m2)** | Model 1 (n=496,267) | Reference | 2.07 (1.92-2.22) | 1.35 (1.28-1.42) | 2.58 (2.44-2.71) | 1.11 (1.07-1.16) |
|  | Model 2b (n=440,681) | Reference | 1.55 (1.44-1.68) | 1.26 (1.19-1.33) | 1.60 (1.51-1.70) | 1.07 (1.03-1.12) |
| **Overweight at booking (BMI 25 to 29.9 kg per m2)** | Model 1 (n=496,267) | Reference | 0.99 (0.96-1.03) | 1.02 (1.00-1.04) | 0.87 (0.84-0.89) | 1.01 (0.99-1.02) |
|  | Model 2b (n=440,681) | Reference | 0.95 (0.92-0.99) | 0.96 (0.94-0.98) | 0.94 (0.91-0.96) | 1.01 (1.00-1.03) |
| **Obesity at booking (BMI greater than or equal to 30 kg per m2)** | Model 1 (n=496,267) | Reference | 0.70 (0.67-0.73) | 0.70 (0.68-0.72) | 1.01 (0.99-1.04) | 1.03 (1.01-1.04) |
|  | Model 2b (n=440,681) | Reference | 0.66 (0.63-0.69) | 0.71 (0.69-0.73) | 0.91 (0.88-0.94) | 1.00 (0.99-1.02) |
| **At least one mental or physical health condition** | Model 1 (n=652,880) | Reference | 0.44 (0.43-0.45) | 0.72 (0.71-0.74) | 0.98 (0.96-1.00) | 0.27 (0.27-0.28) |
|  | Model 2a (n=652,871) | Reference | 0.54 (0.52-0.56) | 0.83 (0.82-0.85) | 1.06 (1.03-1.08) | 0.28 (0.28-0.28) |
| **At least one mental health condition** | Model 1 (n=652,880) | Reference | 0.31 (0.29-0.33) | 0.42 (0.41-0.44) | 1.69 (1.65-1.74) | 0.32 (0.31-0.33) |
|  | Model 2a (n=652,871) | Reference | 0.36 (0.34-0.39) | 0.53 (0.51-0.54) | 1.41 (1.37-1.46) | 0.32 (0.31-0.33) |
| **At least one physical health condition** | Model 1 (n=652,880) | Reference | 0.50 (0.48-0.52) | 0.85 (0.83-0.86) | 0.69 (0.67-0.71) | 0.28 (0.28-0.29) |
|  | Model 2a (n=652,871) | Reference | 0.62 (0.60-0.64) | 0.94 (0.92-0.96) | 0.82 (0.80-0.84) | 0.29 (0.29-0.30) |
| **Diabetes** | Model 1 (n=652,880) | Reference | 1.46 (1.31-1.61) | 1.43 (1.33-1.53) | 0.93 (0.83-1.03) | 0.41 (0.39-0.44) |
|  | Model 2a (n=652,871) | Reference | 1.30 (1.17-1.45) | 1.18 (1.10-1.27) | 1.21 (1.09-1.35) | 0.41 (0.39-0.44) |
| **Hypertension** | Model 1 (n=652,880) | Reference | 0.60 (0.52-0.68) | 0.89 (0.82-0.95) | 0.62 (0.55-0.69) | 0.30 (0.28-0.32) |
|  | Model 2a (n=652,871) | Reference | 0.67 (0.58-0.77) | 0.89 (0.82-0.95) | 0.83 (0.74-0.93) | 0.31 (0.29-0.33) |
| **Cardiac disease** | Model 1 (n=652,880) | Reference | 0.50 (0.42-0.59) | 0.82 (0.75-0.90) | 0.84 (0.75-0.94) | 0.41 (0.38-0.44) |
|  | Model 2a (n=652,871) | Reference | 0.61 (0.51-0.72) | 1.00 (0.91-1.09) | 0.79 (0.70-0.89) | 0.42 (0.39-0.45) |
| **Thromboembolic condition** | Model 1 (n=652,880) | Reference | 0.59 (0.49-0.70) | 0.95 (0.87-1.05) | 0.69 (0.60-0.79) | 0.31 (0.29-0.34) |
|  | Model 2a (n=652,871) | Reference | 0.80 (0.66-0.96) | 1.12 (1.02-1.24) | 0.88 (0.76-1.02) | 0.34 (0.31-0.37) |
| **Renal disease** | Model 1 (n=652,880) | Reference | 0.79 (0.68-0.92) | 1.02 (0.93-1.11) | 0.87 (0.77-0.98) | 0.61 (0.57-0.65) |
|  | Model 2a (n=652,871) | Reference | 0.97 (0.83-1.13) | 1.25 (1.14-1.36) | 0.79 (0.69-0.89) | 0.63 (0.59-0.68) |
| **Hepatitis B** | Model 1 (n=652,880) | Reference | 7.39 (6.12-8.90) | 5.88 (5.05-6.86) | 1.00 (0.69-1.40) | 0.56 (0.45-0.69) |
|  | Model 2a (n=652,871) | Reference | 5.40 (4.43-6.58) | 4.44 (3.78-5.21) | 1.20 (0.83-1.69) | 0.54 (0.44-0.67) |
| **Cancer** | Model 1 (n=652,880) | Reference | 0.33 (0.20-0.49) | 0.62 (0.50-0.76) | 0.58 (0.44-0.76) | 0.31 (0.26-0.37) |
|  | Model 2a (n=652,871) | Reference | 0.53 (0.33-0.81) | 0.77 (0.62-0.95) | 0.87 (0.64-1.14) | 0.35 (0.29-0.41) |
| **Inherited conditions** | Model 1 (n=652,880) | Reference | 0.36 (0.31-0.41) | 0.71 (0.67-0.75) | 1.15 (1.07-1.22) | 0.59 (0.56-0.61) |
|  | Model 2a (n=652,871) | Reference | 0.44 (0.39-0.51) | 0.85 (0.80-0.91) | 1.08 (1.00-1.15) | 0.61 (0.58-0.64) |
| **Family history of diabetes** | Model 1 (n=652,880) | Reference | 1.13 (1.10-1.17) | 1.27 (1.24-1.29) | 0.97 (0.94-0.99) | 0.26 (0.26-0.27) |
|  | Model 2a (n=652,871) | Reference | 0.89 (0.86-0.92) | 1.01 (0.99-1.03) | 1.00 (0.97-1.02) | 0.25 (0.25-0.26) |
| **Booking after 10 weeks gestation** | Model 1 (n=652,309) | Reference | 2.66 (2.59-2.73) | 1.35 (1.33-1.38) | 1.44 (1.41-1.47) | 1.19 (1.18-1.21) |
|  | Model 2b (n=548,368) | Reference | 2.25 (2.18-2.32) | 1.22 (1.20-1.24) | 1.38 (1.34-1.42) | 1.15 (1.14-1.17) |
| **Booking after 16 weeks gestation** | Model 1 (n=652,309) | Reference | 5.04 (4.88-5.22) | 1.62 (1.57-1.67) | 2.43 (2.35-2.52) | 1.92 (1.88-1.96) |
|  | Model 2b (n=548,368) | Reference | 3.69 (3.54-3.83) | 1.43 (1.38-1.48) | 2.23 (2.12-2.34) | 1.65 (1.61-1.70) |
| **Booking after 20 weeks gestation** | Model 1 (n=652,309) | Reference | 4.69 (4.50-4.88) | 1.45 (1.39-1.51) | 2.39 (2.29-2.49) | 2.17 (2.11-2.22) |
|  | Model 2b (n=548,368) | Reference | 3.69 (3.53-3.86) | 1.30 (1.24-1.35) | 2.13 (2.03-2.23) | 1.84 (1.78-1.89) |

### Social factors

Probable migrants had higher odds of being unemployed and seeking work compared to unlikely migrants in all models including unadjusted (Odds Ratio (OR) 2.58, 95% CI 2.46-2.70) and fully adjusted models (OR 1.74, 95% CI 1.66-1.83). Probable migrants had slightly lower odds of not having adequate support available during and after pregnancy in the unadjusted model (OR 0.89, 95% CI 0.82-0.96) but slightly increased odds in the fully adjusted model (OR 1.10, 95% CI 1.02-1.19). Possible migrants (due to English not being their first language or complex social factors) had higher odds of no adequate support available during and after pregnancy in both unadjusted and adjusted models.

### Reproductive health

Probable migrants had higher odds of previous obstetric complications compared to unlikely migrants in all analyses including the unadjusted (OR 1.42, 95% CI 1.37-1.47) and fully adjusted (OR 1.26, 95% CI 1.21-1.31) models. Higher odds of previous obstetric complications were also seen for possible migrants due to English not being their first language. Conversely, the group with missing migration status had around half the odds of previous obstetric complications compared to unlikely migrants in unadjusted and adjusted models. Probable migrants had lower odds of previous pregnancy loss in the unadjusted model (OR 0.91 95% CI 0.88-0.95), but this difference reversed in the fully adjusted model (OR 1.06, 95% CI 1.02-1.11). Those with missing migration status had higher odds of previous pregnancy loss in adjusted and unadjusted models.

Probable migrants had higher odds of previous gestational diabetes mellitus and previous caesarean section in unadjusted and adjusted models, and lower odds of previous pre-eclampsia, eclampsia, gestational proteinuria or HELLP (haemolysis, elevated liver enzyme levels, and low platelet levels) in both unadjusted and adjusted models.

### Health behaviours

Probable migrants had over double the odds of not taking folic acid supplements before pregnancy compared to unlikely migrants in both unadjusted (OR 3.04, 95% CI 2.92-3.16) and fully adjusted (OR 2.15, 95% CI 2.06-2.25) models. Increased odds of not taking folic acid preconception were also found for both groups of possible migrants, although the magnitude was smaller. Those with missing migration status had very slightly higher odds of not taking folic acid before pregnancy.

Probable migrants had lower odds of smoking around conception compared to the unlikely migrant group in all models including the unadjusted (OR 0.89, 95% CI 0.86.0.92) and fully adjusted (OR 0.86, 95% CI 0.82-0.90) models. Possible migrants due to only complex social factors had higher odds of smoking in all models. Probable migrants who smoked had higher odds of not quitting smoking during the year before pregnancy than unlikely migrants who smoked in both unadjusted (OR 1.63, 95% CI 1.49-1.79) and adjusted (OR 1.56, 95% CI 1.41-1.73) models.

### Weight and pre-existing conditions

Probable migrants had increased odds of being underweight in both unadjusted (OR 2.07, 95% CI 1.92-2.22) and fully adjusted (OR 1.55, 95% CI 1.44-1.68) models, as did both groups of possible migrants. Probable migrants had similar odds of being overweight at booking in the unadjusted model, but borderline decreased odds in the fully adjusted model (OR 0.95, 95% CI 0.92-0.99). Probable migrants had lower odds of obesity at booking in all models including the unadjusted (OR 0.70, 95% CI 0.67-0.73) and fully adjusted models (OR 0.66, 95% CI 0.63-0.69), and were also consistently less likely to have at least one mental health condition (OR 0.31, 95% CI 0.29-0.33 and OR 0.36, 95% CI 0.34-0.39 for unadjusted and fully adjusted models respectively) or at least one physical health condition (OR 0.50, 95% CI 0.48-0.52 and OR 0.62, 95% CI 0.60-0.64 in unadjusted and adjusted models respectively).

Probable migrants had higher odds of diabetes in unadjusted (OR 1.46, 95% CI 1.31-1.61) and fully adjusted (OR .30, 95% CI 1.17-1.45) models, and higher odds of hepatitis b in unadjusted (OR 7.39, 95% CI 6.12-8.90) and fully adjusted models (OR 5.40, 95% CI 4.43-5.58). Probable migrants had lower odds of hypertension, cardiac disease, thromboembolic conditions, renal disease, cancer and inherited conditions in all models. In terms of family history of diabetes, probable migrants had higher odds in the unadjusted model (OR 1.13, 95% CI 1.10-1.17) but slightly reduced odds in the fully adjusted model (OR 0.89, 95% CI 0.86-0.92).

### Late antenatal booking

Probable migrants had over double the odds of booking after 10 weeks gestation compared to unlikely migrants in all models (OR 2.66, 95% CI 2.59-2.73 in unadjusted model and OR 2.25, 95% CI 2.18-2.32 in fully adjusted model), and over three times the odds of booking after 16 and 20 weeks gestation in all models (OR 5.04, 95% CI 4.88-5.22 and OR 4.69, 95% CI 4.50-4.88 respectively for unadjusted models and OR 3.69, 95% CI 3.54-3.83 and OR 3.69, 95% CI 3.53-3.86 respectively for fully adjusted models). Increased odds of late booking of a lower magnitude were also identified for both groups of possible migrants and for those with missing migration status data.

### Sensitivity analysis

Excluding ethnicity as a covariate, participants with missing migration status and/or ethnicity, and participants who were under 20 and/or substance users (Models 3a/b, 4a/b and 5a/b respectively in Supplementary Table 1) from regression analyses did not have a substantial influence on most preconception indicators. The exception to this included the preconception indicator of ‘no adequate support available during and after pregnancy’, for which model 2 (including adjustment for ethnicity) found very slightly higher odds for probable migrants compared with unlikely migrants, model 3a (without adjustment for ethnicity) found slightly lower odds, and models 4a (fully adjusted model excluding participants with missing migration status and/or ethnicity) and 5a (fully adjusted model excluding under 20s and substance misusers) found no significant difference. Similarly, for ‘family history of diabetes’, only the models not adjusting for ethnicity (models 1 and 3a) showed slightly higher odds of family history of diabetes for probable migrants compared with unlikely migrants, whereas those adjusting for ethnicity (models 2a, 4a and 5a) showed slightly lower odds. For the ‘overweight’ indicator, Models 1 and 3b showed no increase in odds among probable migrants, whereas Models 2b, 4b and 5b (all additionally adjusting for ethnicity), showed borderline lower odds of being overweight among probable migrants. Slightly lower odds of previous pregnancy loss in unadjusted analysis reversed in adjusted analyses with increased odds in models adjusting for ethnicity (Models 2b, 4b and 5b) and no difference in odds in Model 3b. Lower odds of renal disease among probable migrants in unadjusted analysis and Model 3a became non-significant in models 2a, 4a and 5a.

## DISCUSSION

This is the first study to describe preconception indicators according to migration status in the UK. We have found that women identified as probable migrants have over twice the odds of not taking folic acid before pregnancy and of having their first antenatal booking appointment after the recommended 10 weeks gestation^14^ compared to unlikely migrants, even after adjusting for most deprived deprivation decile, age of mother at booking, number of previous live births and ethnicity. This suggests that migration status has an independent association with these indicators. Probable migrants also have increased odds of previous obstetric complications and of being underweight at booking, but lower odds of recorded physical and mental health conditions (apart from diabetes and hepatitis b), smoking and obesity (in unadjusted and adjusted analyses). These findings highlight areas of interest for policy and practice aiming to reduce inequalities and improve migrant women’s preconception health.

Higher odds of pre-existing diabetes among women identified as probable migrants, even after adjusting for ethnicity, most deprived decile, and age at booking, indicates that diabetes is an area of particular importance for migrant women’s preconception health. Ensuring women have good blood glucose control preconception and during pregnancy reduces risks of miscarriage, congenital anomalies, neonatal death and stillbirth^28,29^, and NICE guidelines reiterate the importance of pregnancy planning for women with diabetes^29^. Despite this, just 17.5% of women with type 1 diabetes and 38.4% of women with type 2 diabetes have the recommended HbA1c (blood glucose) levels in the first trimester, indicating inadequate diabetic control preconception^30^. However, little is known about pregnancy planning among women with diabetes who are migrants.

Lower odds of folic acid supplementation in the preconception period is also a serious concern for migrant maternal health: maternal folate supplementation preconception is associated with fewer anomaly-related terminations and a reduction in neural tube defects^8^. Moreover, being underweight preconception increases odds of small for gestational age, low birth weight and preterm birth^7^. Conversely, lower odds of many physical and mental health conditions among probable migrants may not be a positive finding: it could indicate lack of diagnosis due to, for example, lower primary care access rates among migrants compared to non-migrants^26^; lack of diagnosis could mean preconception care that could support a healthy pregnancy does not happen.

### Comparison with other studies

This study provides the first description of preconception indicators among migrant women compared to non-migrant women in England. In contrast to this study’s findings of lower odds of obesity in the probable migrant group, a Swedish population-based study found that individuals born in North Africa and Middle East and Sub-Saharan Africa were more likely to be obese during pregnancy than Swedish-born women^31^, and a Finnish population-based study found that women of Kurdish, Somali and Russian origin had higher BMIs before pregnancy than women in the general Finnish population^32^. Migrants from countries with lower levels of obesity often gain unhealthy weight after migration^33,34^, which may explain why lower levels of obesity among probable migrants were identified in this study. Moreover, BMI-based cardiovascular risk varies between ethnicities^35^ and lower BMI levels are associated with type 2 diabetes among south Asian, black, Chinese and Arab populations^36^. Thus, obesity remains an important preconception risk factor in this population.

Late antenatal booking among migrant women has been identified in several previous studies^10,22,23^. A narrative synthesis systematic review of access to and interventions to improve maternity care services for migrant women found that language barriers, poor awareness of service availability, not understanding the reason for antenatal appointments and barriers due to immigration status and income meant that migrant women were more likely to access antenatal care late^23^. As well as improving healthy behaviours and understanding of the importance of antenatal care before pregnancy, preconception care could help women navigate a complex health system.

### Strengths

A major strength of this study is the comprehensive nature of MSDS, representing the majority of pregnant women in England within the study period, and resulting in a study population largely representative of the English population of pregnant people^37,38^. Migration status is rarely recorded within electronic health records^6^, so the presence of a recorded complex social factors variable as well as English not a first language within MSDS creates a unique opportunity to examine the influence of migration status on preconception indicators, focusing on migrants in potentially vulnerable situations.

### Limitations

The main limitation of this study is uncertainty in the definition of migration status due to missing data for this exposure (missing data were present for 33.8% of women for either complex social factors and/or English as a first language). However, sensitivity analysis undertaken during logistic regression analyses indicated that missing data did not have a substantial impact on outcomes. There were also missing data for most preconception indicators, ranging from 0% of records for pre-existing health conditions to 38.9% of records with missing data for alcohol drinking at booking, which may have impacted results. The true level of missing data relating to pre-existing conditions is likely to be higher because those with missing diagnosis data were recorded in the dataset as not having the diagnosis, so it was not possible for us to differentiate between missing data or true absence of a diagnosis. For variables related to pre-existing health conditions, it is unclear whether lower prevalence rates in the probable migrant group compared to unlikely migrant group for many conditions are because of a true difference in prevalence, underdiagnosis or non-disclosure (missing data were assumed to be ‘no’). Those with ‘missing’ migration status had the lowest odds of having mental or physical health conditions, indicating that in some participants’ records, there may be low reporting of several variables and not just those related to migration status (either at an individual level, or systematic under-reporting at hospital, NHS Trust or IT-system level). Further research is needed to identify whether and how differences in pre-existing mental and physical health conditions preconception exist between migrant women and UK-born women.

Uncertainty due to the method of identifying migrant women is a risk; some women who experience domestic abuse and do not speak English as their first language, but were born in the UK, may have been included in the probable migrant group. Similarly, there may be some women born in the UK who do not speak English as their first language and were identified in our analysis as ‘probable migrants’ due to their age being under 20 or their misuse of alcohol and/or drugs (rather than being asylum seekers, refugees or recent migrants); however, sensitivity analysis was undertaken to exclude most of these individuals in logistic regression analyses, with no substantial effect on outcomes. Finally, by including multiple preconception indicators, there is a risk of identifying false-positive (or negative) findings due to multiple testing^39^. However, due to the large sample size of this study, levels of precision are high.

In conclusion, we have shown that migrant women in potentially vulnerable situations are less likely to take folic acid during the preconception period than non-migrants, are more likely to have certain underlying health issues including diabetes, hepatitis b and being underweight at booking, and are more likely to have had previous obstetric complications than non-migrants. They are also less likely to have their first antenatal appointment within the recommended 10 weeks. These findings highlight the opportunity for more comprehensive preconception care for migrant women in potentially vulnerable situations, who are already known to have worse perinatal outcomes^4^. Further research could explore inequalities at more granular levels of migration status (e.g. asylum seeker, refugee or region of origin), and identify barriers and facilitators to improving preconception health in this population.

## Data Availability

Data used within this study were collected for the national Maternity Services Dataset. Data are available on request to NHS Digital (https://digital.nhs.uk/services/data-access-request-service-dars).

## Acknowledgements

The authors would like to thank Helen Smith (OHID) for her support in data quality assessment, as well as Kate Thurland, Professor Robert Aldridge and Dr Ines Campos-Matos for their role in the project advisory group. We would also like to thank the community advisory group for their invaluable input into the planning of this study.

## Competing interests

All authors have completed the ICMJE uniform disclosure form at www.icmje.org/coi_disclosure.pdf and declare: MM has support from the Medical Research Council [Grant number MR/W01498X/1]; DS is supported by NIHR Southampton Biomedical Research Centre; HD is National lead for lifecourse intelligence at the Office for Health Improvement and Disparities, Department of Health and Social Care; DB reports grants from the NIHR RfPB Programme, the NIHR HS&DS Programme and the NIHR HTA Programme and is Chair of Trustees of the MASIC Charity; FB reports grants from MRC, NIHR, Novartis Gene Therapies, Wellcome Trust and Public Health England and is a member of the Foetal, Maternal and Child Health Reference Group of the UK National Screening Committee and a Member of Bloodspot Advisory Group; SM reports being on the Advisory Group on Contraception and on the NHSEI Advisory Group on postnatal care; no financial relationships with any organisations that might have an interest in the submitted work in the previous three years; no other relationships or activities that could appear to have influenced the submitted work.

## Contributors

MM, OO, HD and EA designed the study. MM undertook the data analysis, which EA quality assessed. OO provided advisory support regarding data analysis with input from DS and SM. MM wrote the draft manuscript. All authors contributed to the editing of the final manuscript. The corresponding author attests that all listed authors meet authorship criteria and that no others meeting the criteria have been omitted. MM is the guarantor.

## Funding

MM is supported by the Medical Research Council [Grant number MR/W01498X/1]. For the purpose of open access, the author has applied a Creative Commons Attribution (CC BY) licence. DS is supported by the National Institute for Health and Social Care Research (NIHR) Southampton Biomedical Research Centre [IS-BRC-1215-20004]. The views expressed are those of the author(s) and not necessarily those of MRC, UKRI, NIHR or the Department of Health and Social Care.

The lead author (the manuscript’s guarantor) affirms that the manuscript is an honest, accurate, and transparent account of the study being reported; that no important aspects of the study have been omitted; and that any discrepancies from the study as originally planned (and, if relevant, registered) have been explained.

Dissemination to participants and related patient and public communities: The results of this research will be presented to migrant women and other stakeholders at workshops run in partnership with third sector organisations. Findings will be shared with relevant royal colleges and professional associations, including the UK Preconception Partnership, the Faculty of Public Health, the Royal College of GPs, the Royal College of Midwives and the Royal College of Nursing. Findings will also be shared through social media. A link to the study will be available on the gov.uk website via the Office for Health Improvement and Disparities.

