## Supplementary File 1 for "Preconception health among migrant women in England: a cross-sectional analysis of maternity services data 2018-2019"

Supplementary Table 1: Full logistic regression models for all preconception health indicators

*Model 1 is unadjusted. Model 2a adjusts for most deprived Index of Multiple Deprivation (IMD) decile, mother’s age at booking and ethnicity; Model 2b additionally adjusts for number of previous live births. Model 3a adjusts for most deprived IMD decile and mother’s age at booking; Model 3b additionally adjusts for number of previous live births. Model 4a adjusts for most deprived IMD decile, mother’s age at booking and ethnicity but excludes those with missing data for complex social factors and/or English as a first language and/or ethnicity; Model 4b additionally adjusts for number of previous live births. Model 5a adjusts for most deprived IMD decile, mother’s age at booking and ethnicity but excludes those under age 20 and those with recorded substance misuse; Model 5b additionally adjusts for number of previous live births. LCI = lower confidence interval. UCI = upper confidence interval.* *Complex social factors include women who misuse alcohol and/or drugs; women who are recent migrants, asylum seekers or refugees, or have difficulty reading or speaking English; women aged under 20; and/or women who experience domestic abuse.*

| **Outcome: mother unemployed and seeking work** | | | | | | |
| --- | --- | --- | --- | --- | --- | --- |
| **Migration status** | **Odds ratio** | **95% LCI** | **95% UCI** | **P value** | **Number of observations** | **Pseudo R squared (Hosmer-Lemeshow)** |
| **Model 1** | | | | | | |
| Unlikely migrant (English as first language without complex social factors) | Reference | Reference | Reference | Reference | 472,181 | 0.04 |
| Probable migrant (English not their first language and complex social factors) | 2.58 | 2.46 | 2.70 | 0.000 |  |  |
| Possible migrant (only English not their first language) | 1.03 | 0.98 | 1.07 | 0.235 |  |  |
| Possible migrant (only complex social factors) | 5.10 | 4.92 | 5.28 | 0.000 |  |  |
| Missing migration status (first language or complex social factor data missing) | 1.09 | 1.05 | 1.13 | 0.000 |  |  |
| **Model 2a** | | | | | | |
| Unlikely migrant (English as first language without complex social factors) | Reference | Reference | Reference | Reference | 472,175 | 0.084 |
| Probable migrant (English not their first language and complex social factors) | 1.74 | 1.66 | 1.83 | 0.000 |  |  |
| Possible migrant (only English not their first language) | 0.96 | 0.92 | 1.01 | 0.093 |  |  |
| Possible migrant (only complex social factors) | 2.84 | 2.73 | 2.95 | 0.000 |  |  |
| Missing migration status (first language or complex social factor data missing) | 1.03 | 0.99 | 1.07 | 0.116 |  |  |
| Not most deprived | Reference | Reference | Reference | Reference |  |  |
| Most deprived | 2.39 | 2.32 | 2.46 | 0.000 |  |  |
| Mother's age at booking (years) | 0.92 | 0.92 | 0.92 | 0.000 |  |  |
| White | Reference | Reference | Reference | Reference |  |  |
| Mixed | 1.56 | 1.44 | 1.68 | 0.000 |  |  |
| Asian | 1.07 | 1.02 | 1.12 | 0.003 |  |  |
| Black | 1.70 | 1.61 | 1.79 | 0.000 |  |  |
| Other | 1.25 | 1.16 | 1.33 | 0.000 |  |  |
| Missing ethnicity | 0.87 | 0.84 | 0.91 | 0.000 |  |  |
| **Model 3a** | | | | | | |
| Unlikely migrant (English as first language without complex social factors) | Reference | Reference | Reference | Reference | 472,175 | 0.082 |
| Probable migrant (English not their first language and complex social factors) | 1.81 | 1.72 | 1.90 | 0.000 |  |  |
| Possible migrant (only English not their first language) | 0.99 | 0.95 | 1.03 | 0.570 |  |  |
| Possible migrant (only complex social factors) | 2.87 | 2.76 | 2.98 | 0.000 |  |  |
| Missing migration status (first language or complex social factor data missing) | 1.04 | 1.00 | 1.08 | 0.037 |  |  |
| Not most deprived | Reference | Reference | Reference | Reference |  |  |
| Most deprived | 2.44 | 2.37 | 2.51 | 0.000 |  |  |
| Mother's age at booking (years) | 0.92 | 0.92 | 0.93 | 0.000 |  |  |
| **Model 4a** | | | | | | |
| Unlikely migrant (English as first language without complex social factors) | Reference | Reference | Reference | Reference | 330,963 | 0.091 |
| Probable migrant (English not their first language and complex social factors) | 1.79 | 1.69 | 1.89 | 0.000 |  |  |
| Possible migrant (only English not their first language) | 0.99 | 0.95 | 1.04 | 0.772 |  |  |
| Possible migrant (only complex social factors) | 2.86 | 2.75 | 2.98 | 0.000 |  |  |
| Not most deprived | Reference | Reference | Reference | Reference |  |  |
| Most deprived | 2.34 | 2.26 | 2.43 | 0.000 |  |  |
| Mother's age at booking (years) | 0.92 | 0.92 | 0.93 | 0.000 |  |  |
| White | Reference | Reference | Reference | Reference |  |  |
| Mixed | 1.56 | 1.43 | 1.70 | 0.000 |  |  |
| Asian | 0.94 | 0.89 | 0.98 | 0.011 |  |  |
| Black | 1.63 | 1.53 | 1.72 | 0.000 |  |  |
| Other | 1.14 | 1.06 | 1.23 | 0.001 |  |  |
| **Model 5a** | | | | | | |
| Unlikely migrant (English as first language without complex social factors) | Reference | Reference | Reference | Reference | 450,819 | 0.065 |
| Probable migrant (English not their first language and complex social factors) | 1.74 | 1.65 | 1.84 | 0.000 |  |  |
| Possible migrant (only English not their first language) | 0.97 | 0.93 | 1.01 | 0.156 |  |  |
| Possible migrant (only complex social factors) | 3.11 | 2.96 | 3.27 | 0.000 |  |  |
| Missing migration status (first language or complex social factor data missing) | 0.95 | 0.91 | 0.98 | 0.006 |  |  |
| Not most deprived | Reference | Reference | Reference | Reference |  |  |
| Most deprived | 2.50 | 2.42 | 2.58 | 0.000 |  |  |
| Mother's age at booking (years) | 0.91 | 0.91 | 0.92 | 0.000 |  |  |
| White | Reference | Reference | Reference | Reference |  |  |
| Mixed | 1.66 | 1.52 | 1.80 | 0.000 |  |  |
| Asian | 1.07 | 1.02 | 1.12 | 0.006 |  |  |
| Black | 1.77 | 1.68 | 1.87 | 0.000 |  |  |
| Other | 1.29 | 1.20 | 1.39 | 0.000 |  |  |
| Missing ethnicity | 0.88 | 0.85 | 0.92 | 0.000 |  |  |

| **Outcome: no adequate support available during and after pregnancy** | | | | | | |
| --- | --- | --- | --- | --- | --- | --- |
| **Migration status** | **Odds ratio** | **95% LCI** | **95% UCI** | **P value** | **Number of observations** | **Pseudo R squared (Hosmer-Lemeshow)** |
| **Model 1** | | | | | | |
| Unlikely migrant (English as first language without complex social factors) | Reference | Reference | Reference | Reference | 449,884 | 0.055 |
| Probable migrant (English not their first language and complex social factors) | 0.89 | 0.82 | 0.96 | 0.003 |  |  |
| Possible migrant (only English not their first language) | 1.47 | 1.41 | 1.53 | 0.000 |  |  |
| Possible migrant (only complex social factors) | 1.36 | 1.29 | 1.44 | 0.000 |  |  |
| Missing migration status (first language or complex social factor data missing) | 4.46 | 4.33 | 4.59 | 0.000 |  |  |
| **Model 2a** | | | | | | |
| Unlikely migrant (English as first language without complex social factors) | Reference | Reference | Reference | Reference | 449,877 | 0.062 |
| Probable migrant (English not their first language and complex social factors) | 1.10 | 1.02 | 1.19 | 0.018 |  |  |
| Possible migrant (only English not their first language) | 1.75 | 1.67 | 1.82 | 0.000 |  |  |
| Possible migrant (only complex social factors) | 1.37 | 1.29 | 1.45 | 0.000 |  |  |
| Missing migration status (first language or complex social factor data missing) | 4.56 | 4.43 | 4.69 | 0.000 |  |  |
| Not most deprived | Reference | Reference | Reference | Reference |  |  |
| Most deprived | 0.85 | 0.82 | 0.88 | 0.000 |  |  |
| Mother's age at booking (years) | 1.00 | 1.00 | 1.00 | 0.678 |  |  |
| White | Reference | Reference | Reference | Reference |  |  |
| Mixed | 0.73 | 0.66 | 0.81 | 0.000 |  |  |
| Asian | 0.44 | 0.41 | 0.46 | 0.000 |  |  |
| Black | 0.57 | 0.53 | 0.61 | 0.000 |  |  |
| Other | 0.63 | 0.58 | 0.68 | 0.000 |  |  |
| Missing ethnicity | 0.82 | 0.79 | 0.85 | 0.000 |  |  |
| **Model 3a** | | | | | | |
| Unlikely migrant (English as first language without complex social factors) | Reference | Reference | Reference | Reference | 449,877 | 0.056 |
| Probable migrant (English not their first language and complex social factors) | 0.91 | 0.84 | 0.99 | 0.022 |  |  |
| Possible migrant (only English not their first language) | 1.49 | 1.43 | 1.55 | 0.000 |  |  |
| Possible migrant (only complex social factors) | 1.37 | 1.29 | 1.45 | 0.000 |  |  |
| Missing migration status (first language or complex social factor data missing) | 4.47 | 4.35 | 4.60 | 0.000 |  |  |
| Not most deprived | Reference | Reference | Reference | Reference |  |  |
| Most deprived | 0.82 | 0.78 | 0.85 | 0.000 |  |  |
| Mother's age at booking (years) | 1.00 | 0.99 | 1.00 | 0.005 |  |  |
| **Model 4a** | | | | | | |
| Unlikely migrant (English as first language without complex social factors) | Reference | Reference | Reference | Reference | 316,493 | 0.014 |
| Probable migrant (English not their first language and complex social factors) | 1.05 | 0.96 | 1.15 | 0.264 |  |  |
| Possible migrant (only English not their first language) | 1.81 | 1.73 | 1.90 | 0.000 |  |  |
| Possible migrant (only complex social factors) | 1.45 | 1.36 | 1.54 | 0.000 |  |  |
| Not most deprived | Reference | Reference | Reference | Reference |  |  |
| Most deprived | 0.58 | 0.54 | 0.61 | 0.000 |  |  |
| Mother's age at booking (years) | 1.02 | 1.02 | 1.02 | 0.000 |  |  |
| White | Reference | Reference | Reference | Reference |  |  |
| Mixed | 0.86 | 0.76 | 0.98 | 0.020 |  |  |
| Asian | 0.50 | 0.47 | 0.53 | 0.000 |  |  |
| Black | 0.74 | 0.68 | 0.80 | 0.000 |  |  |
| Other | 0.60 | 0.54 | 0.66 | 0.000 |  |  |
| **Model 5a** | | | | | | |
| Unlikely migrant (English as first language without complex social factors) | Reference | Reference | Reference | Reference | 431,079 | 0.062 |
| Probable migrant (English not their first language and complex social factors) | 1.02 | 0.93 | 1.11 | 0.638 |  |  |
| Possible migrant (only English not their first language) | 1.75 | 1.67 | 1.82 | 0.000 |  |  |
| Possible migrant (only complex social factors) | 1.33 | 1.23 | 1.43 | 0.000 |  |  |
| Missing migration status (first language or complex social factor data missing) | 4.49 | 4.36 | 4.62 | 0.000 |  |  |
| Not most deprived | Reference | Reference | Reference | Reference |  |  |
| Most deprived | 0.84 | 0.81 | 0.88 | 0.000 |  |  |
| Mother's age at booking (years) | 1.00 | 1.00 | 1.01 | 0.000 |  |  |
| White | Reference | Reference | Reference | Reference |  |  |
| Mixed | 0.74 | 0.66 | 0.82 | 0.000 |  |  |
| Asian | 0.44 | 0.41 | 0.46 | 0.000 |  |  |
| Black | 0.56 | 0.52 | 0.60 | 0.000 |  |  |
| Other | 0.63 | 0.58 | 0.68 | 0.000 |  |  |
| Missing ethnicity | 0.81 | 0.78 | 0.84 | 0.000 |  |  |

| **Outcome: previous obstetric complication** | | | | | | |
| --- | --- | --- | --- | --- | --- | --- |
| **Migration status** | **Odds ratio** | **95% LCI** | **95% UCI** | **P value** | **Number of observations** | **Pseudo R squared (Hosmer-Lemeshow)** |
| **Model 1** | | | | | | |
| Unlikely migrant (English as first language without complex social factors) | Reference | Reference | Reference | Reference | 329,228 | 0.023 |
| Probable migrant (English not their first language and complex social factors) | 1.42 | 1.37 | 1.47 | 0.000 |  |  |
| Possible migrant (only English not their first language) | 1.33 | 1.30 | 1.36 | 0.000 |  |  |
| Possible migrant (only complex social factors) | 0.87 | 0.84 | 0.90 | 0.000 |  |  |
| Missing migration status (first language or complex social factor data missing) | 0.44 | 0.43 | 0.45 | 0.000 |  |  |
| **Model 2b** | | | | | | |
| Unlikely migrant (English as first language without complex social factors) | Reference | Reference | Reference | Reference | 325,619 | 0.041 |
| Probable migrant (English not their first language and complex social factors) | 1.26 | 1.21 | 1.31 | 0.000 |  |  |
| Possible migrant (only English not their first language) | 1.18 | 1.15 | 1.21 | 0.000 |  |  |
| Possible migrant (only complex social factors) | 0.91 | 0.88 | 0.95 | 0.000 |  |  |
| Missing migration status (first language or complex social factor data missing) | 0.43 | 0.42 | 0.44 | 0.000 |  |  |
| Not most deprived | Reference | Reference | Reference | Reference |  |  |
| Most deprived | 1.09 | 1.06 | 1.11 | 0.000 |  |  |
| Mother's age at booking (years) | 1.03 | 1.03 | 1.03 | 0.000 |  |  |
| Number of previous live births | 1.22 | 1.21 | 1.23 | 0.000 |  |  |
| White | Reference | Reference | Reference | Reference |  |  |
| Mixed | 1.12 | 1.06 | 1.19 | 0.000 |  |  |
| Asian | 1.46 | 1.42 | 1.50 | 0.000 |  |  |
| Black | 1.21 | 1.17 | 1.25 | 0.000 |  |  |
| Other | 0.99 | 0.95 | 1.04 | 0.807 |  |  |
| Missing ethnicity | 0.94 | 0.91 | 0.96 | 0.000 |  |  |
| **Model 3b** | | | | | | |
| Unlikely migrant (English as first language without complex social factors) | Reference | Reference | Reference | Reference | 325,619 | 0.039 |
| Probable migrant (English not their first language and complex social factors) | 1.35 | 1.30 | 1.40 | 0.000 |  |  |
| Possible migrant (only English not their first language) | 1.27 | 1.24 | 1.31 | 0.000 |  |  |
| Possible migrant (only complex social factors) | 0.90 | 0.87 | 0.94 | 0.000 |  |  |
| Missing migration status (first language or complex social factor data missing) | 0.44 | 0.43 | 0.44 | 0.000 |  |  |
| Not most deprived | Reference | Reference | Reference | Reference |  |  |
| Most deprived | 1.11 | 1.08 | 1.14 | 0.000 |  |  |
| Mother's age at booking (years) | 1.03 | 1.03 | 1.03 | 0.000 |  |  |
| Number of previous live births | 1.23 | 1.22 | 1.23 | 0.000 |  |  |
| **Model 4b** | | | | | | |
| Unlikely migrant (English as first language without complex social factors) | Reference | Reference | Reference | Reference | 216,001 | 0.028 |
| Probable migrant (English not their first language and complex social factors) | 1.22 | 1.17 | 1.27 | 0.000 |  |  |
| Possible migrant (only English not their first language) | 1.17 | 1.14 | 1.20 | 0.000 |  |  |
| Possible migrant (only complex social factors) | 0.94 | 0.90 | 0.98 | 0.002 |  |  |
| Not most deprived | Reference | Reference | Reference | Reference |  |  |
| Most deprived | 1.15 | 1.12 | 1.19 | 0.000 |  |  |
| Mother's age at booking (years) | 1.03 | 1.03 | 1.03 | 0.000 |  |  |
| Number of previous live births | 1.24 | 1.23 | 1.25 | 0.000 |  |  |
| White | Reference | Reference | Reference | Reference |  |  |
| Mixed | 1.22 | 1.14 | 1.30 | 0.000 |  |  |
| Asian | 1.54 | 1.50 | 1.58 | 0.000 |  |  |
| Black | 1.32 | 1.27 | 1.37 | 0.000 |  |  |
| Other | 1.02 | 0.96 | 1.07 | 0.551 |  |  |
| **Model 5b** | | | | | | |
| Unlikely migrant (English as first language without complex social factors) | Reference | Reference | Reference | Reference | 317,443 | 0.039 |
| Probable migrant (English not their first language and complex social factors) | 1.27 | 1.22 | 1.32 | 0.000 |  |  |
| Possible migrant (only English not their first language) | 1.17 | 1.14 | 1.20 | 0.000 |  |  |
| Possible migrant (only complex social factors) | 0.97 | 0.93 | 1.01 | 0.168 |  |  |
| Missing migration status (first language or complex social factor data missing) | 0.44 | 0.43 | 0.45 | 0.000 |  |  |
| Not most deprived | Reference | Reference | Reference | Reference |  |  |
| Most deprived | 1.10 | 1.08 | 1.13 | 0.000 |  |  |
| Mother's age at booking (years) | 1.03 | 1.03 | 1.03 | 0.000 |  |  |
| Number of previous live births | 1.22 | 1.21 | 1.23 | 0.000 |  |  |
| White | Reference | Reference | Reference | Reference |  |  |
| Mixed | 1.13 | 1.06 | 1.20 | 0.000 |  |  |
| Asian | 1.47 | 1.43 | 1.51 | 0.000 |  |  |
| Black | 1.22 | 1.17 | 1.26 | 0.000 |  |  |
| Other | 0.99 | 0.95 | 1.04 | 0.754 |  |  |
| Missing ethnicity | 0.94 | 0.91 | 0.96 | 0.000 |  |  |

| **Outcome: previous pre-eclampsia, HELLP, eclampsia or gestational proteinuria** | | | | | | |
| --- | --- | --- | --- | --- | --- | --- |
| **Migration status** | **Odds ratio** | **95% LCI** | **95% UCI** | **P value** | **Number of observations** | **Pseudo R squared (Hosmer-Lemeshow)** |
| **Model 1** | | | | | | |
| Unlikely migrant (English as first language without complex social factors) | Reference | Reference | Reference | Reference | 329,228 | 0.002 |
| Probable migrant (English not their first language and complex social factors) | 0.78 | 0.65 | 0.93 | 0.007 |  |  |
| Possible migrant (only English not their first language) | 0.97 | 0.88 | 1.08 | 0.585 |  |  |
| Possible migrant (only complex social factors) | 0.70 | 0.59 | 0.83 | 0.000 |  |  |
| Missing migration status (first language or complex social factor data missing) | 0.69 | 0.63 | 0.75 | 0.000 |  |  |
| **Model 2b** | | | | | | |
| Unlikely migrant (English as first language without complex social factors) | Reference | Reference | Reference | Reference | 325,619 | 0.012 |
| Probable migrant (English not their first language and complex social factors) | 0.69 | 0.57 | 0.83 | 0.000 |  |  |
| Possible migrant (only English not their first language) | 0.94 | 0.84 | 1.04 | 0.250 |  |  |
| Possible migrant (only complex social factors) | 0.64 | 0.54 | 0.76 | 0.000 |  |  |
| Missing migration status (first language or complex social factor data missing) | 0.67 | 0.62 | 0.73 | 0.000 |  |  |
| Not most deprived | Reference | Reference | Reference | Reference |  |  |
| Most deprived | 1.32 | 1.20 | 1.44 | 0.000 |  |  |
| Mother's age at booking (years) | 1.00 | 0.99 | 1.01 | 0.827 |  |  |
| Number of previous live births | 1.21 | 1.18 | 1.24 | 0.000 |  |  |
| White | Reference | Reference | Reference | Reference |  |  |
| Mixed | 1.12 | 0.88 | 1.41 | 0.330 |  |  |
| Asian | 0.93 | 0.82 | 1.04 | 0.205 |  |  |
| Black | 1.58 | 1.39 | 1.79 | 0.000 |  |  |
| Other | 0.67 | 0.52 | 0.84 | 0.001 |  |  |
| Missing ethnicity | 1.09 | 0.98 | 1.21 | 0.119 |  |  |
| **Model 3b** | | | | | | |
| Unlikely migrant (English as first language without complex social factors) | Reference | Reference | Reference | Reference | 325,619 | 0.01 |
| Probable migrant (English not their first language and complex social factors) | 0.68 | 0.57 | 0.82 | 0.000 |  |  |
| Possible migrant (only English not their first language) | 0.93 | 0.84 | 1.03 | 0.161 |  |  |
| Possible migrant (only complex social factors) | 0.64 | 0.54 | 0.76 | 0.000 |  |  |
| Missing migration status (first language or complex social factor data missing) | 0.67 | 0.62 | 0.74 | 0.000 |  |  |
| Not most deprived | Reference | Reference | Reference | Reference |  |  |
| Most deprived | 1.34 | 1.23 | 1.46 | 0.000 |  |  |
| Mother's age at booking (years) | 1.00 | 1.00 | 1.01 | 0.560 |  |  |
| Number of previous live births | 1.21 | 1.18 | 1.24 | 0.000 |  |  |
| **Model 4b** | | | | | | |
| Unlikely migrant (English as first language without complex social factors) | Reference | Reference | Reference | Reference | 216,001 | 0.013 |
| Probable migrant (English not their first language and complex social factors) | 0.70 | 0.57 | 0.85 | 0.000 |  |  |
| Possible migrant (only English not their first language) | 0.95 | 0.85 | 1.07 | 0.410 |  |  |
| Possible migrant (only complex social factors) | 0.67 | 0.56 | 0.80 | 0.000 |  |  |
| Not most deprived | Reference | Reference | Reference | Reference |  |  |
| Most deprived | 1.33 | 1.20 | 1.48 | 0.000 |  |  |
| Mother's age at booking (years) | 1.00 | 1.00 | 1.01 | 0.213 |  |  |
| Number of previous live births | 1.21 | 1.17 | 1.24 | 0.000 |  |  |
| White | Reference | Reference | Reference | Reference |  |  |
| Mixed | 1.29 | 1.00 | 1.64 | 0.045 |  |  |
| Asian | 0.99 | 0.87 | 1.12 | 0.868 |  |  |
| Black | 1.82 | 1.59 | 2.08 | 0.000 |  |  |
| Other | 0.69 | 0.52 | 0.88 | 0.005 |  |  |
| **Model 5b** | | | | | | |
| Unlikely migrant (English as first language without complex social factors) | Reference | Reference | Reference | Reference | 317,443 | 0.012 |
| Probable migrant (English not their first language and complex social factors) | 0.70 | 0.58 | 0.84 | 0.000 |  |  |
| Possible migrant (only English not their first language) | 0.94 | 0.84 | 1.04 | 0.223 |  |  |
| Possible migrant (only complex social factors) | 0.58 | 0.47 | 0.70 | 0.000 |  |  |
| Missing migration status (first language or complex social factor data missing) | 0.69 | 0.63 | 0.75 | 0.000 |  |  |
| Not most deprived | Reference | Reference | Reference | Reference |  |  |
| Most deprived | 1.32 | 1.21 | 1.44 | 0.000 |  |  |
| Mother's age at booking (years) | 1.00 | 0.99 | 1.01 | 0.722 |  |  |
| Number of previous live births | 1.21 | 1.18 | 1.23 | 0.000 |  |  |
| White | Reference | Reference | Reference | Reference |  |  |
| Mixed | 1.14 | 0.89 | 1.43 | 0.272 |  |  |
| Asian | 0.94 | 0.83 | 1.05 | 0.271 |  |  |
| Black | 1.61 | 1.41 | 1.82 | 0.000 |  |  |
| Other | 0.67 | 0.52 | 0.84 | 0.001 |  |  |
| Missing ethnicity | 1.08 | 0.97 | 1.20 | 0.149 |  |  |

| **Outcome: previous gestational hypertension** | | | | | | |
| --- | --- | --- | --- | --- | --- | --- |
| **Migration status** | **Odds ratio** | **95% LCI** | **95% UCI** | **P value** | **Number of observations** | **Pseudo R squared (Hosmer-Lemeshow)** |
| **Model 1** | | | | | | |
| Unlikely migrant (English as first language without complex social factors) | Reference | Reference | Reference | Reference | 329,228 | 0.007 |
| Probable migrant (English not their first language and complex social factors) | 1.02 | 0.89 | 1.16 | 0.768 |  |  |
| Possible migrant (only English not their first language) | 0.91 | 0.83 | 0.99 | 0.027 |  |  |
| Possible migrant (only complex social factors) | 1.10 | 0.98 | 1.22 | 0.115 |  |  |
| Missing migration status (first language or complex social factor data missing) | 0.50 | 0.47 | 0.54 | 0.000 |  |  |
| **Model 2b** | | | | | | |
| Unlikely migrant (English as first language without complex social factors) | Reference | Reference | Reference | Reference | 325,619 | 0.017 |
| Probable migrant (English not their first language and complex social factors) | 0.97 | 0.85 | 1.11 | 0.689 |  |  |
| Possible migrant (only English not their first language) | 0.89 | 0.81 | 0.97 | 0.008 |  |  |
| Possible migrant (only complex social factors) | 1.07 | 0.95 | 1.20 | 0.243 |  |  |
| Missing migration status (first language or complex social factor data missing) | 0.50 | 0.47 | 0.55 | 0.000 |  |  |
| Not most deprived | Reference | Reference | Reference | Reference |  |  |
| Most deprived | 1.17 | 1.09 | 1.26 | 0.000 |  |  |
| Mother's age at booking (years) | 1.02 | 1.01 | 1.02 | 0.000 |  |  |
| Number of previous live births | 1.21 | 1.18 | 1.23 | 0.000 |  |  |
| White | Reference | Reference | Reference | Reference |  |  |
| Mixed | 0.98 | 0.80 | 1.19 | 0.827 |  |  |
| Asian | 1.02 | 0.93 | 1.11 | 0.676 |  |  |
| Black | 1.12 | 1.00 | 1.25 | 0.051 |  |  |
| Other | 0.75 | 0.62 | 0.89 | 0.001 |  |  |
| Missing ethnicity | 0.70 | 0.63 | 0.77 | 0.000 |  |  |
| **Model 3b** | | | | | | |
| Unlikely migrant (English as first language without complex social factors) | Reference | Reference | Reference | Reference | 325,619 | 0.016 |
| Probable migrant (English not their first language and complex social factors) | 1.08 | 0.95 | 1.23 | 0.257 |  |  |
| Possible migrant (only English not their first language) | 1.15 | 1.06 | 1.25 | 0.001 |  |  |
| Possible migrant (only complex social factors) | 0.93 | 0.83 | 1.05 | 0.219 |  |  |
| Missing migration status (first language or complex social factor data missing) | 2.00 | 1.85 | 2.17 | 0.000 |  |  |
| Not most deprived | Reference | Reference | Reference | Reference |  |  |
| Most deprived | 0.85 | 0.79 | 0.92 | 0.000 |  |  |
| Mother's age at booking (years) | 0.98 | 0.98 | 0.99 | 0.000 |  |  |
| Number of previous live births | 0.82 | 0.81 | 0.84 | 0.000 |  |  |
| **Model 4b** | | | | | | |
| Unlikely migrant (English as first language without complex social factors) | Reference | Reference | Reference | Reference | 216,001 | 0.011 |
| Probable migrant (English not their first language and complex social factors) | 0.96 | 0.83 | 1.11 | 0.620 |  |  |
| Possible migrant (only English not their first language) | 0.89 | 0.81 | 0.98 | 0.019 |  |  |
| Possible migrant (only complex social factors) | 1.06 | 0.94 | 1.19 | 0.355 |  |  |
| Not most deprived | Reference | Reference | Reference | Reference |  |  |
| Most deprived | 1.21 | 1.11 | 1.31 | 0.000 |  |  |
| Mother's age at booking (years) | 1.02 | 1.01 | 1.02 | 0.000 |  |  |
| Number of previous live births | 1.21 | 1.18 | 1.24 | 0.000 |  |  |
| White | Reference | Reference | Reference | Reference |  |  |
| Mixed | 1.04 | 0.84 | 1.28 | 0.698 |  |  |
| Asian | 1.03 | 0.93 | 1.13 | 0.542 |  |  |
| Black | 1.17 | 1.03 | 1.32 | 0.013 |  |  |
| Other | 0.76 | 0.63 | 0.92 | 0.005 |  |  |
| **Model 5b** | | | | | | |
| Unlikely migrant (English as first language without complex social factors) | Reference | Reference | Reference | Reference | 317,443 | 0.017 |
| Probable migrant (English not their first language and complex social factors) | 1.00 | 0.87 | 1.14 | 0.946 |  |  |
| Possible migrant (only English not their first language) | 0.89 | 0.81 | 0.97 | 0.007 |  |  |
| Possible migrant (only complex social factors) | 1.17 | 1.04 | 1.32 | 0.009 |  |  |
| Missing migration status (first language or complex social factor data missing) | 0.52 | 0.48 | 0.56 | 0.000 |  |  |
| Not most deprived | Reference | Reference | Reference | Reference |  |  |
| Most deprived | 1.19 | 1.10 | 1.28 | 0.000 |  |  |
| Mother's age at booking (years) | 1.02 | 1.01 | 1.02 | 0.000 |  |  |
| Number of previous live births | 1.20 | 1.18 | 1.23 | 0.000 |  |  |
| White | Reference | Reference | Reference | Reference |  |  |
| Mixed | 0.99 | 0.81 | 1.20 | 0.936 |  |  |
| Asian | 1.02 | 0.93 | 1.11 | 0.673 |  |  |
| Black | 1.12 | 1.00 | 1.26 | 0.044 |  |  |
| Other | 0.75 | 0.63 | 0.90 | 0.002 |  |  |
| Missing ethnicity | 0.69 | 0.62 | 0.76 | 0.000 |  |  |

| **Outcome: previous gestational diabetes** | | | | | | |
| --- | --- | --- | --- | --- | --- | --- |
| **Migration status** | **Odds ratio** | **95% LCI** | **95% UCI** | **P value** | **Number of observations** | **Pseudo R squared (Hosmer-Lemeshow)** |
| **Model 1** | | | | | | |
| Unlikely migrant (English as first language without complex social factors) | Reference | Reference | Reference | Reference | 329,228 | 0.017 |
| Probable migrant (English not their first language and complex social factors) | 1.64 | 1.49 | 1.80 | 0.000 |  |  |
| Possible migrant (only English not their first language) | 1.97 | 1.86 | 2.09 | 0.000 |  |  |
| Possible migrant (only complex social factors) | 0.63 | 0.55 | 0.71 | 0.000 |  |  |
| Missing migration status (first language or complex social factor data missing) | 0.58 | 0.54 | 0.62 | 0.000 |  |  |
| **Model 2b** | | | | | | |
| Unlikely migrant (English as first language without complex social factors) | Reference | Reference | Reference | Reference | 325,619 | 0.053 |
| Probable migrant (English not their first language and complex social factors) | 1.12 | 1.01 | 1.24 | 0.024 |  |  |
| Possible migrant (only English not their first language) | 1.36 | 1.28 | 1.45 | 0.000 |  |  |
| Possible migrant (only complex social factors) | 0.68 | 0.59 | 0.77 | 0.000 |  |  |
| Missing migration status (first language or complex social factor data missing) | 0.55 | 0.51 | 0.59 | 0.000 |  |  |
| Not most deprived | Reference | Reference | Reference | Reference |  |  |
| Most deprived | 1.12 | 1.05 | 1.19 | 0.000 |  |  |
| Mother's age at booking (years) | 1.04 | 1.03 | 1.04 | 0.000 |  |  |
| Number of previous live births | 1.24 | 1.22 | 1.26 | 0.000 |  |  |
| White | Reference | Reference | Reference | Reference |  |  |
| Mixed | 1.26 | 1.06 | 1.49 | 0.008 |  |  |
| Asian | 2.97 | 2.80 | 3.15 | 0.000 |  |  |
| Black | 1.51 | 1.37 | 1.66 | 0.000 |  |  |
| Other | 1.37 | 1.21 | 1.55 | 0.000 |  |  |
| Missing ethnicity | 1.09 | 1.01 | 1.19 | 0.030 |  |  |
| **Model 3b** | | | | | | |
| Unlikely migrant (English as first language without complex social factors) | Reference | Reference | Reference | Reference | 325,619 | 0.035 |
| Probable migrant (English not their first language and complex social factors) | 1.49 | 1.35 | 1.64 | 0.000 |  |  |
| Possible migrant (only English not their first language) | 1.86 | 1.75 | 1.97 | 0.000 |  |  |
| Possible migrant (only complex social factors) | 0.65 | 0.56 | 0.73 | 0.000 |  |  |
| Missing migration status (first language or complex social factor data missing) | 0.57 | 0.53 | 0.61 | 0.000 |  |  |
| Not most deprived | Reference | Reference | Reference | Reference |  |  |
| Most deprived | 1.20 | 1.13 | 1.28 | 0.000 |  |  |
| Mother's age at booking (years) | 1.04 | 1.04 | 1.05 | 0.000 |  |  |
| Number of previous live births | 1.24 | 1.22 | 1.26 | 0.000 |  |  |
| **Model 4b** | | | | | | |
| Unlikely migrant (English as first language without complex social factors) | Reference | Reference | Reference | Reference | 216,001 | 0.051 |
| Probable migrant (English not their first language and complex social factors) | 1.11 | 0.99 | 1.23 | 0.065 |  |  |
| Possible migrant (only English not their first language) | 1.36 | 1.27 | 1.45 | 0.000 |  |  |
| Possible migrant (only complex social factors) | 0.69 | 0.60 | 0.79 | 0.000 |  |  |
| Not most deprived | Reference | Reference | Reference | Reference |  |  |
| Most deprived | 1.10 | 1.03 | 1.18 | 0.007 |  |  |
| Mother's age at booking (years) | 1.04 | 1.03 | 1.04 | 0.000 |  |  |
| Number of previous live births | 1.24 | 1.22 | 1.27 | 0.000 |  |  |
| White | Reference | Reference | Reference | Reference |  |  |
| Mixed | 1.34 | 1.11 | 1.60 | 0.002 |  |  |
| Asian | 3.01 | 2.82 | 3.21 | 0.000 |  |  |
| Black | 1.58 | 1.43 | 1.75 | 0.000 |  |  |
| Other | 1.30 | 1.13 | 1.49 | 0.000 |  |  |
| **Model 5b** | | | | | | |
| Unlikely migrant (English as first language without complex social factors) | Reference | Reference | Reference | Reference | 317,443 | 0.052 |
| Probable migrant (English not their first language and complex social factors) | 1.13 | 1.02 | 1.24 | 0.019 |  |  |
| Possible migrant (only English not their first language) | 1.36 | 1.28 | 1.45 | 0.000 |  |  |
| Possible migrant (only complex social factors) | 0.68 | 0.59 | 0.78 | 0.000 |  |  |
| Missing migration status (first language or complex social factor data missing) | 0.57 | 0.53 | 0.61 | 0.000 |  |  |
| Not most deprived | Reference | Reference | Reference | Reference |  |  |
| Most deprived | 1.13 | 1.06 | 1.20 | 0.000 |  |  |
| Mother's age at booking (years) | 1.04 | 1.03 | 1.04 | 0.000 |  |  |
| Number of previous live births | 1.24 | 1.22 | 1.26 | 0.000 |  |  |
| White | Reference | Reference | Reference | Reference |  |  |
| Mixed | 1.27 | 1.07 | 1.50 | 0.006 |  |  |
| Asian | 2.98 | 2.81 | 3.16 | 0.000 |  |  |
| Black | 1.52 | 1.38 | 1.67 | 0.000 |  |  |
| Other | 1.37 | 1.21 | 1.55 | 0.000 |  |  |
| Missing ethnicity | 1.10 | 1.01 | 1.19 | 0.028 |  |  |

| **Outcome: previous caesarean section** | | | | | | |
| --- | --- | --- | --- | --- | --- | --- |
| **Migration status** | **Odds ratio** | **95% LCI** | **95% UCI** | **P value** | **Number of observations** | **Pseudo R squared (Hosmer-Lemeshow)** |
| **Model 1** | | | | | | |
| Unlikely migrant (English as first language without complex social factors) | Reference | Reference | Reference | Reference | 306,429 | 0.001 |
| Probable migrant (English not their first language and complex social factors) | 1.23 | 1.18 | 1.28 | 0.000 |  |  |
| Possible migrant (only English not their first language) | 1.21 | 1.18 | 1.24 | 0.000 |  |  |
| Possible migrant (only complex social factors) | 0.86 | 0.82 | 0.89 | 0.000 |  |  |
| Missing migration status (first language or complex social factor data missing) | 0.98 | 0.96 | 1.00 | 0.055 |  |  |
| **Model 2b** | | | | | | |
| Unlikely migrant (English as first language without complex social factors) | Reference | Reference | Reference | Reference | 298,229 | 0.02 |
| Probable migrant (English not their first language and complex social factors) | 1.13 | 1.08 | 1.18 | 0.000 |  |  |
| Possible migrant (only English not their first language) | 1.07 | 1.04 | 1.10 | 0.000 |  |  |
| Possible migrant (only complex social factors) | 1.02 | 0.98 | 1.07 | 0.363 |  |  |
| Missing migration status (first language or complex social factor data missing) | 0.97 | 0.95 | 0.99 | 0.002 |  |  |
| Not most deprived | Reference | Reference | Reference | Reference |  |  |
| Most deprived | 0.98 | 0.95 | 1.00 | 0.068 |  |  |
| Mother's age at booking (years) | 1.05 | 1.04 | 1.05 | 0.000 |  |  |
| Number of previous live births | 1.14 | 1.13 | 1.15 | 0.000 |  |  |
| White | Reference | Reference | Reference | Reference |  |  |
| Mixed | 1.03 | 0.97 | 1.10 | 0.372 |  |  |
| Asian | 1.36 | 1.32 | 1.40 | 0.000 |  |  |
| Black | 1.42 | 1.36 | 1.47 | 0.000 |  |  |
| Other | 1.22 | 1.16 | 1.28 | 0.000 |  |  |
| Missing ethnicity | 1.05 | 1.02 | 1.08 | 0.000 |  |  |
| **Model 3b** | | | | | | |
| Unlikely migrant (English as first language without complex social factors) | Reference | Reference | Reference | Reference | 298,229 | 0.017 |
| Probable migrant (English not their first language and complex social factors) | 1.24 | 1.19 | 1.29 | 0.000 |  |  |
| Possible migrant (only English not their first language) | 1.16 | 1.13 | 1.19 | 0.000 |  |  |
| Possible migrant (only complex social factors) | 1.02 | 0.97 | 1.06 | 0.472 |  |  |
| Missing migration status (first language or complex social factor data missing) | 0.98 | 0.96 | 1.00 | 0.037 |  |  |
| Not most deprived | Reference | Reference | Reference | Reference |  |  |
| Most deprived | 1.01 | 0.98 | 1.03 | 0.637 |  |  |
| Mother's age at booking (years) | 1.05 | 1.05 | 1.05 | 0.000 |  |  |
| Number of previous live births | 1.14 | 1.13 | 1.15 | 0.000 |  |  |
| **Model 4b** | | | | | | |
| Unlikely migrant (English as first language without complex social factors) | Reference | Reference | Reference | Reference | 208,684 | 0.021 |
| Probable migrant (English not their first language and complex social factors) | 1.11 | 1.06 | 1.16 | 0.000 |  |  |
| Possible migrant (only English not their first language) | 1.07 | 1.04 | 1.10 | 0.000 |  |  |
| Possible migrant (only complex social factors) | 0.97 | 0.93 | 1.01 | 0.165 |  |  |
| Not most deprived | Reference | Reference | Reference | Reference |  |  |
| Most deprived | 1.00 | 0.97 | 1.03 | 0.883 |  |  |
| Mother's age at booking (years) | 1.05 | 1.04 | 1.05 | 0.000 |  |  |
| Number of previous live births | 1.14 | 1.13 | 1.15 | 0.000 |  |  |
| White | Reference | Reference | Reference | Reference |  |  |
| Mixed | 1.02 | 0.95 | 1.10 | 0.498 |  |  |
| Asian | 1.35 | 1.31 | 1.39 | 0.000 |  |  |
| Black | 1.29 | 1.23 | 1.34 | 0.000 |  |  |
| Other | 1.17 | 1.11 | 1.24 | 0.000 |  |  |
| **Model 5b** |  |  |  |  |  |  |
| Unlikely migrant (English as first language without complex social factors) | Reference | Reference | Reference | Reference | 298,229 | 0.02 |
| Probable migrant (English not their first language and complex social factors) | 1.13 | 1.08 | 1.18 | 0.000 |  |  |
| Possible migrant (only English not their first language) | 1.07 | 1.04 | 1.10 | 0.000 |  |  |
| Possible migrant (only complex social factors) | 1.02 | 0.98 | 1.07 | 0.363 |  |  |
| Missing migration status (first language or complex social factor data missing) | 0.97 | 0.95 | 0.99 | 0.002 |  |  |
| Not most deprived | Reference | Reference | Reference | Reference |  |  |
| Most deprived | 0.98 | 0.95 | 1.00 | 0.068 |  |  |
| Mother's age at booking (years) | 1.05 | 1.04 | 1.05 | 0.000 |  |  |
| Number of previous live births | 1.14 | 1.13 | 1.15 | 0.000 |  |  |
| White | Reference | Reference | Reference | Reference |  |  |
| Mixed | 1.03 | 0.97 | 1.10 | 0.372 |  |  |
| Asian | 1.36 | 1.32 | 1.40 | 0.000 |  |  |
| Black | 1.42 | 1.36 | 1.47 | 0.000 |  |  |
| Other | 1.22 | 1.16 | 1.28 | 0.000 |  |  |
| Missing ethnicity | 1.05 | 1.02 | 1.08 | 0.000 |  |  |

| **Outcome: previous pregnancy loss** | | | | | | |
| --- | --- | --- | --- | --- | --- | --- |
| **Migration status** | **Odds ratio** | **95% LCI** | **95% UCI** | **P value** | **Number of observations** | **Pseudo R squared (Hosmer-Lemeshow)** |
| **Model 1** | | | | | | |
| Unlikely migrant (English as first language without complex social factors) | Reference | Reference | Reference | Reference | 301,168 | 0.008 |
| Probable migrant (English not their first language and complex social factors) | 0.91 | 0.88 | 0.95 | 0.000 |  |  |
| Possible migrant (only English not their first language) | 0.79 | 0.77 | 0.81 | 0.000 |  |  |
| Possible migrant (only complex social factors) | 1.63 | 1.58 | 1.69 | 0.000 |  |  |
| Missing migration status (first language or complex social factor data missing) | 1.45 | 1.43 | 1.48 | 0.000 |  |  |
| **Model 2b** | | | | | | |
| Unlikely migrant (English as first language without complex social factors) | Reference | Reference | Reference | Reference | 289,853 | 0.032 |
| Probable migrant (English not their first language and complex social factors) | 1.06 | 1.02 | 1.11 | 0.004 |  |  |
| Possible migrant (only English not their first language) | 0.85 | 0.83 | 0.87 | 0.000 |  |  |
| Possible migrant (only complex social factors) | 1.78 | 1.72 | 1.86 | 0.000 |  |  |
| Missing migration status (first language or complex social factor data missing) | 1.45 | 1.42 | 1.47 | 0.000 |  |  |
| Not most deprived | Reference | Reference | Reference | Reference |  |  |
| Most deprived | 1.13 | 1.11 | 1.16 | 0.000 |  |  |
| Mother's age at booking (years) | 1.03 | 1.02 | 1.03 | 0.000 |  |  |
| Number of previous live births | 0.69 | 0.69 | 0.70 | 0.000 |  |  |
| White | Reference | Reference | Reference | Reference |  |  |
| Mixed | 1.23 | 1.16 | 1.29 | 0.000 |  |  |
| Asian | 0.75 | 0.73 | 0.77 | 0.000 |  |  |
| Black | 1.16 | 1.12 | 1.20 | 0.000 |  |  |
| Other | 0.93 | 0.89 | 0.97 | 0.002 |  |  |
| Missing ethnicity | 0.87 | 0.85 | 0.89 | 0.000 |  |  |
| **Model 3b** | | | | | | |
| Unlikely migrant (English as first language without complex social factors) | Reference | Reference | Reference | Reference | 289,853 | 0.03 |
| Probable migrant (English not their first language and complex social factors) | 1.00 | 0.96 | 1.04 | 0.923 |  |  |
| Possible migrant (only English not their first language) | 0.80 | 0.78 | 0.82 | 0.000 |  |  |
| Possible migrant (only complex social factors) | 1.80 | 1.73 | 1.88 | 0.000 |  |  |
| Missing migration status (first language or complex social factor data missing) | 1.43 | 1.40 | 1.46 | 0.000 |  |  |
| Not most deprived | Reference | Reference | Reference | Reference |  |  |
| Most deprived | 1.13 | 1.10 | 1.15 | 0.000 |  |  |
| Mother's age at booking (years) | 1.02 | 1.02 | 1.03 | 0.000 |  |  |
| Number of previous live births | 0.69 | 0.69 | 0.70 | 0.000 |  |  |
| **Model 4b** | | | | | | |
| Unlikely migrant (English as first language without complex social factors) | Reference | Reference | Reference | Reference | 197,699 | 0.033 |
| Probable migrant (English not their first language and complex social factors) | 1.13 | 1.08 | 1.18 | 0.000 |  |  |
| Possible migrant (only English not their first language) | 0.85 | 0.83 | 0.88 | 0.000 |  |  |
| Possible migrant (only complex social factors) | 1.86 | 1.79 | 1.94 | 0.000 |  |  |
| Not most deprived | Reference | Reference | Reference | Reference |  |  |
| Most deprived | 1.20 | 1.17 | 1.24 | 0.000 |  |  |
| Mother's age at booking (years) | 1.03 | 1.03 | 1.03 | 0.000 |  |  |
| Number of previous live births | 0.67 | 0.66 | 0.67 | 0.000 |  |  |
| White | Reference | Reference | Reference | Reference |  |  |
| Mixed | 1.28 | 1.20 | 1.36 | 0.000 |  |  |
| Asian | 0.77 | 0.75 | 0.80 | 0.000 |  |  |
| Black | 1.18 | 1.13 | 1.23 | 0.000 |  |  |
| Other | 0.91 | 0.87 | 0.96 | 0.001 |  |  |
| **Model 5b** | | | | | | |
| Unlikely migrant (English as first language without complex social factors) | Reference | Reference | Reference | Reference | 289,853 | 0.032 |
| Probable migrant (English not their first language and complex social factors) | 1.06 | 1.02 | 1.11 | 0.004 |  |  |
| Possible migrant (only English not their first language) | 0.85 | 0.83 | 0.87 | 0.000 |  |  |
| Possible migrant (only complex social factors) | 1.78 | 1.72 | 1.86 | 0.000 |  |  |
| Missing migration status (first language or complex social factor data missing) | 1.45 | 1.42 | 1.47 | 0.000 |  |  |
| Not most deprived | Reference | Reference | Reference | Reference |  |  |
| Most deprived | 1.13 | 1.11 | 1.16 | 0.000 |  |  |
| Mother's age at booking (years) | 1.03 | 1.02 | 1.03 | 0.000 |  |  |
| Number of previous live births | 0.69 | 0.69 | 0.70 | 0.000 |  |  |
| White | Reference | Reference | Reference | Reference |  |  |
| Mixed | 1.23 | 1.16 | 1.29 | 0.000 |  |  |
| Asian | 0.75 | 0.73 | 0.77 | 0.000 |  |  |
| Black | 1.16 | 1.12 | 1.20 | 0.000 |  |  |
| Other | 0.93 | 0.89 | 0.97 | 0.002 |  |  |
| Missing ethnicity | 0.87 | 0.85 | 0.89 | 0.000 |  |  |

| **Outcome: not taking folic acid before conception** | | | | | | |
| --- | --- | --- | --- | --- | --- | --- |
| **Migration status** | **Odds ratio** | **95% LCI** | **95% UCI** | **P value** | **Number of observations** | **Pseudo R squared (Hosmer-Lemeshow)** |
| **Model 1** | | | | | | |
| Unlikely migrant (English as first language without complex social factors) | Reference | Reference | Reference | Reference | 488,987 | 0.012 |
| Probable migrant (English not their first language and complex social factors) | 3.04 | 2.92 | 3.16 | 0.000 |  |  |
| Possible migrant (only English not their first language) | 1.29 | 1.27 | 1.32 | 0.000 |  |  |
| Possible migrant (only complex social factors) | 2.38 | 2.31 | 2.46 | 0.000 |  |  |
| Missing migration status (first language or complex social factor data missing) | 1.06 | 1.05 | 1.08 | 0.000 |  |  |
| **Model 2b** |  |  |  |  |  |  |
| Unlikely migrant (English as first language without complex social factors) | Reference | Reference | Reference | Reference | 422,527 | 0.073 |
| Probable migrant (English not their first language and complex social factors) | 2.15 | 2.06 | 2.25 | 0.000 |  |  |
| Possible migrant (only English not their first language) | 1.22 | 1.19 | 1.25 | 0.000 |  |  |
| Possible migrant (only complex social factors) | 1.49 | 1.44 | 1.55 | 0.000 |  |  |
| Missing migration status (first language or complex social factor data missing) | 1.09 | 1.07 | 1.11 | 0.000 |  |  |
| Not most deprived | Reference | Reference | Reference | Reference |  |  |
| Most deprived | 1.67 | 1.63 | 1.71 | 0.000 |  |  |
| Mother's age at booking (years) | 0.91 | 0.91 | 0.92 | 0.000 |  |  |
| Number of previous live births | 1.54 | 1.52 | 1.55 | 0.000 |  |  |
| White | Reference | Reference | Reference | Reference |  |  |
| Mixed | 1.36 | 1.29 | 1.44 | 0.000 |  |  |
| Asian | 1.43 | 1.40 | 1.47 | 0.000 |  |  |
| Black | 1.95 | 1.87 | 2.03 | 0.000 |  |  |
| Other | 1.39 | 1.33 | 1.45 | 0.000 |  |  |
| Missing ethnicity | 1.18 | 1.15 | 1.20 | 0.000 |  |  |
| **Model 3b** | | | | | | |
| Unlikely migrant (English as first language without complex social factors) | Reference | Reference | Reference | Reference | 422,527 | 0.069 |
| Probable migrant (English not their first language and complex social factors) | 2.41 | 2.30 | 2.51 | 0.000 |  |  |
| Possible migrant (only English not their first language) | 1.32 | 1.30 | 1.35 | 0.000 |  |  |
| Possible migrant (only complex social factors) | 1.50 | 1.45 | 1.56 | 0.000 |  |  |
| Missing migration status (first language or complex social factor data missing) | 1.09 | 1.07 | 1.11 | 0.000 |  |  |
| Not most deprived | Reference | Reference | Reference | Reference |  |  |
| Most deprived | 1.74 | 1.69 | 1.78 | 0.000 |  |  |
| Mother's age at booking (years) | 0.92 | 0.92 | 0.92 | 0.000 |  |  |
| Number of previous live births | 1.54 | 1.52 | 1.55 | 0.000 |  |  |
| **Model 4b** |  |  |  |  |  |  |
| Unlikely migrant (English as first language without complex social factors) | Reference | Reference | Reference | Reference | 289,190 | 0.073 |
| Probable migrant (English not their first language and complex social factors) | 2.18 | 2.07 | 2.29 | 0.000 |  |  |
| Possible migrant (only English not their first language) | 1.20 | 1.18 | 1.23 | 0.000 |  |  |
| Possible migrant (only complex social factors) | 1.23 | 1.18 | 1.28 | 0.000 |  |  |
| Not most deprived | Reference | Reference | Reference | Reference |  |  |
| Most deprived | 1.62 | 1.57 | 1.67 | 0.000 |  |  |
| Mother's age at booking (years) | 0.91 | 0.91 | 0.91 | 0.000 |  |  |
| Number of previous live births | 1.57 | 1.55 | 1.58 | 0.000 |  |  |
| White | Reference | Reference | Reference | Reference |  |  |
| Mixed | 1.49 | 1.40 | 1.59 | 0.000 |  |  |
| Asian | 1.43 | 1.39 | 1.47 | 0.000 |  |  |
| Black | 2.11 | 2.02 | 2.21 | 0.000 |  |  |
| **Model 5b** |  |  |  |  |  |  |
| Unlikely migrant (English as first language without complex social factors) | Reference | Reference | Reference | Reference | 405,365 | 0.067 |
| Probable migrant (English not their first language and complex social factors) | 2.20 | 2.10 | 2.30 | 0.000 |  |  |
| Possible migrant (only English not their first language) | 1.22 | 1.19 | 1.25 | 0.000 |  |  |
| Possible migrant (only complex social factors) | 1.25 | 1.20 | 1.30 | 0.000 |  |  |
| Missing migration status (first language or complex social factor data missing) | 1.09 | 1.07 | 1.11 | 0.000 |  |  |
| Not most deprived | Reference | Reference | Reference | Reference |  |  |
| Most deprived | 1.69 | 1.64 | 1.73 | 0.000 |  |  |
| Mother's age at booking (years) | 0.92 | 0.92 | 0.92 | 0.000 |  |  |
| Number of previous live births | 1.54 | 1.53 | 1.55 | 0.000 |  |  |
| White | Reference | Reference | Reference | Reference |  |  |
| Mixed | 1.35 | 1.27 | 1.43 | 0.000 |  |  |
| Asian | 1.44 | 1.40 | 1.48 | 0.000 |  |  |
| Black | 1.95 | 1.87 | 2.04 | 0.000 |  |  |
| Other | 1.39 | 1.33 | 1.45 | 0.000 |  |  |
| Missing ethnicity | 1.18 | 1.15 | 1.20 | 0.000 |  |  |

| **Outcome: smoker at conception** | | | | | | |
| --- | --- | --- | --- | --- | --- | --- |
| **Migration status** | **Odds ratio** | **95% LCI** | **95% UCI** | **P value** | **Number of observations** | **Pseudo R squared (Hosmer-Lemeshow)** |
| **Model 1** | | | | | | |
| Unlikely migrant (English as first language without complex social factors) | Reference | Reference | Reference | Reference | 604,514 | 0.018 |
| Probable migrant (English not their first language and complex social factors) | 0.89 | 0.86 | 0.92 | 0.000 |  |  |
| Possible migrant (only English not their first language) | 0.73 | 0.71 | 0.75 | 0.000 |  |  |
| Possible migrant (only complex social factors) | 3.12 | 3.05 | 3.20 | 0.000 |  |  |
| Missing migration status (first language or complex social factor data missing) | 1.11 | 1.10 | 1.13 | 0.000 |  |  |
| **Model 2b** | | | | | | |
| Unlikely migrant (English as first language without complex social factors) | Reference | Reference | Reference | Reference | 481,999 | 0.11 |
| Probable migrant (English not their first language and complex social factors) | 0.86 | 0.82 | 0.90 | 0.000 |  |  |
| Possible migrant (only English not their first language) | 1.03 | 1.00 | 1.06 | 0.037 |  |  |
| Possible migrant (only complex social factors) | 1.80 | 1.74 | 1.87 | 0.000 |  |  |
| Missing migration status (first language or complex social factor data missing) | 1.05 | 1.03 | 1.07 | 0.000 |  |  |
| Not most deprived | Reference | Reference | Reference | Reference |  |  |
| Most deprived | 1.84 | 1.80 | 1.87 | 0.000 |  |  |
| Mother's age at booking (years) | 0.90 | 0.90 | 0.90 | 0.000 |  |  |
| Number of previous live births | 1.37 | 1.36 | 1.38 | 0.000 |  |  |
| White | Reference | Reference | Reference | Reference |  |  |
| Mixed | 0.80 | 0.76 | 0.85 | 0.000 |  |  |
| Asian | 0.12 | 0.11 | 0.13 | 0.000 |  |  |
| Black | 0.23 | 0.21 | 0.24 | 0.000 |  |  |
| Other | 0.43 | 0.41 | 0.46 | 0.000 |  |  |
| Missing ethnicity | 0.64 | 0.62 | 0.65 | 0.000 |  |  |
| **Model 3b** | | | | | | |
| Unlikely migrant (English as first language without complex social factors) | Reference | Reference | Reference | Reference | 481,999 | 0.073 |
| Probable migrant (English not their first language and complex social factors) | 0.57 | 0.55 | 0.59 | 0.000 |  |  |
| Possible migrant (only English not their first language) | 0.73 | 0.71 | 0.75 | 0.000 |  |  |
| Possible migrant (only complex social factors) | 1.76 | 1.70 | 1.82 | 0.000 |  |  |
| Missing migration status (first language or complex social factor data missing) | 1.01 | 0.99 | 1.03 | 0.423 |  |  |
| Not most deprived | Reference | Reference | Reference | Reference |  |  |
| Most deprived | 1.61 | 1.58 | 1.64 | 0.000 |  |  |
| Mother's age at booking (years) | 0.89 | 0.89 | 0.89 | 0.000 |  |  |
| Number of previous live births | 1.37 | 1.36 | 1.38 | 0.000 |  |  |
| **Model 4b** | | | | | | |
| Unlikely migrant (English as first language without complex social factors) | Reference | Reference | Reference | Reference | 317,047 | 0.122 |
| Probable migrant (English not their first language and complex social factors) | 0.90 | 0.86 | 0.95 | 0.000 |  |  |
| Possible migrant (only English not their first language) | 1.05 | 1.02 | 1.09 | 0.001 |  |  |
| Possible migrant (only complex social factors) | 1.80 | 1.73 | 1.87 | 0.000 |  |  |
| Not most deprived | Reference | Reference | Reference | Reference |  |  |
| Most deprived | 1.96 | 1.91 | 2.01 | 0.000 |  |  |
| Mother's age at booking (years) | 0.90 | 0.89 | 0.90 | 0.000 |  |  |
| Number of previous live births | 1.37 | 1.35 | 1.38 | 0.000 |  |  |
| White | Reference | Reference | Reference | Reference |  |  |
| Mixed | 0.80 | 0.75 | 0.86 | 0.000 |  |  |
| Asian | 0.11 | 0.10 | 0.12 | 0.000 |  |  |
| Black | 0.22 | 0.21 | 0.23 | 0.000 |  |  |
| Other | 0.43 | 0.41 | 0.46 | 0.000 |  |  |
| **Model 5b** | | | | | | |
| Unlikely migrant (English as first language without complex social factors) | Reference | Reference | Reference | Reference | 481,999 | 0.11 |
| Probable migrant (English not their first language and complex social factors) | 0.86 | 0.82 | 0.90 | 0.000 |  |  |
| Possible migrant (only English not their first language) | 1.03 | 1.00 | 1.06 | 0.037 |  |  |
| Possible migrant (only complex social factors) | 1.80 | 1.74 | 1.87 | 0.000 |  |  |
| Missing migration status (first language or complex social factor data missing) | 1.05 | 1.03 | 1.07 | 0.000 |  |  |
| Not most deprived | Reference | Reference | Reference | Reference |  |  |
| Most deprived | 1.84 | 1.80 | 1.87 | 0.000 |  |  |
| Mother's age at booking (years) | 0.90 | 0.90 | 0.90 | 0.000 |  |  |
| Number of previous live births | 1.37 | 1.36 | 1.38 | 0.000 |  |  |
| White | Reference | Reference | Reference | Reference |  |  |
| Mixed | 0.80 | 0.76 | 0.85 | 0.000 |  |  |
| Asian | 0.12 | 0.11 | 0.13 | 0.000 |  |  |
| Black | 0.23 | 0.21 | 0.24 | 0.000 |  |  |
| Other | 0.43 | 0.41 | 0.46 | 0.000 |  |  |
| Missing ethnicity | 0.64 | 0.62 | 0.65 | 0.000 |  |  |

| **Outcome: smokers who did not quit smoking during year before pregnancy** | | | | | | |
| --- | --- | --- | --- | --- | --- | --- |
| **Migration status** | **Odds ratio** | **95% LCI** | **95% UCI** | **P value** | **Number of observations** | **Pseudo R squared (Hosmer-Lemeshow)** |
| **Model 1** | | | | | | |
| Unlikely migrant (English as first language without complex social factors) | Reference | Reference | Reference | Reference | 138,422 | 0.009 |
| Probable migrant (English not their first language and complex social factors) | 1.63 | 1.49 | 1.79 | 0.000 |  |  |
| Possible migrant (only English not their first language) | 1.00 | 0.95 | 1.05 | 0.994 |  |  |
| Possible migrant (only complex social factors) | 1.88 | 1.78 | 1.99 | 0.000 |  |  |
| Missing migration status (first language or complex social factor data missing) | 1.56 | 1.50 | 1.61 | 0.000 |  |  |
| **Model 2b** | | | | | | |
| Unlikely migrant (English as first language without complex social factors) | Reference | Reference | Reference | Reference | 104,675 | 0.047 |
| Probable migrant (English not their first language and complex social factors) | 1.56 | 1.41 | 1.73 | 0.000 |  |  |
| Possible migrant (only English not their first language) | 1.21 | 1.15 | 1.28 | 0.000 |  |  |
| Possible migrant (only complex social factors) | 1.33 | 1.23 | 1.44 | 0.000 |  |  |
| Missing migration status (first language or complex social factor data missing) | 1.32 | 1.27 | 1.38 | 0.000 |  |  |
| Not most deprived | Reference | Reference | Reference | Reference |  |  |
| Most deprived | 1.96 | 1.86 | 2.07 | 0.000 |  |  |
| Mother's age at booking (years) | 0.95 | 0.94 | 0.95 | 0.000 |  |  |
| Number of previous live births | 1.45 | 1.42 | 1.47 | 0.000 |  |  |
| White | Reference | Reference | Reference | Reference |  |  |
| Mixed | 0.82 | 0.73 | 0.92 | 0.001 |  |  |
| Asian | 0.78 | 0.70 | 0.86 | 0.000 |  |  |
| Black | 0.66 | 0.59 | 0.74 | 0.000 |  |  |
| Other | 0.85 | 0.76 | 0.96 | 0.006 |  |  |
| Missing ethnicity | 0.80 | 0.77 | 0.85 | 0.000 |  |  |
| **Model 3b** | | | | | | |
| Unlikely migrant (English as first language without complex social factors) | Reference | Reference | Reference | Reference | 104,675 | 0.046 |
| Probable migrant (English not their first language and complex social factors) | 1.52 | 1.37 | 1.68 | 0.000 |  |  |
| Possible migrant (only English not their first language) | 1.19 | 1.12 | 1.26 | 0.000 |  |  |
| Possible migrant (only complex social factors) | 1.33 | 1.23 | 1.44 | 0.000 |  |  |
| Missing migration status (first language or complex social factor data missing) | 1.31 | 1.26 | 1.37 | 0.000 |  |  |
| Not most deprived | Reference | Reference | Reference | Reference |  |  |
| Most deprived | 1.94 | 1.84 | 2.05 | 0.000 |  |  |
| Mother's age at booking (years) | 0.94 | 0.94 | 0.95 | 0.000 |  |  |
| Number of previous live births | 1.46 | 1.43 | 1.48 | 0.000 |  |  |
| **Model 4b** | | | | | | |
| Unlikely migrant (English as first language without complex social factors) | Reference | Reference | Reference | Reference | 70,106 | 0.049 |
| Probable migrant (English not their first language and complex social factors) | 1.51 | 1.34 | 1.70 | 0.000 |  |  |
| Possible migrant (only English not their first language) | 1.23 | 1.15 | 1.31 | 0.000 |  |  |
| Possible migrant (only complex social factors) | 1.30 | 1.20 | 1.41 | 0.000 |  |  |
| Not most deprived | Reference | Reference | Reference | Reference |  |  |
| Most deprived | 2.06 | 1.93 | 2.20 | 0.000 |  |  |
| Mother's age at booking (years) | 0.94 | 0.94 | 0.95 | 0.000 |  |  |
| Number of previous live births | 1.45 | 1.42 | 1.48 | 0.000 |  |  |
| White | Reference | Reference | Reference | Reference |  |  |
| Mixed | 0.81 | 0.71 | 0.92 | 0.001 |  |  |
| Asian | 0.72 | 0.65 | 0.81 | 0.000 |  |  |
| Black | 0.61 | 0.54 | 0.69 | 0.000 |  |  |
| Other | 0.86 | 0.76 | 0.98 | 0.019 |  |  |
| **Model 5b** | | | | | | |
| Unlikely migrant (English as first language without complex social factors) | Reference | Reference | Reference | Reference | 104,675 | 0.047 |
| Probable migrant (English not their first language and complex social factors) | 1.56 | 1.41 | 1.73 | 0.000 |  |  |
| Possible migrant (only English not their first language) | 1.21 | 1.15 | 1.28 | 0.000 |  |  |
| Possible migrant (only complex social factors) | 1.33 | 1.23 | 1.44 | 0.000 |  |  |
| Missing migration status (first language or complex social factor data missing) | 1.32 | 1.27 | 1.38 | 0.000 |  |  |
| Not most deprived | Reference | Reference | Reference | Reference |  |  |
| Most deprived | 1.96 | 1.86 | 2.07 | 0.000 |  |  |
| Mother's age at booking (years) | 0.95 | 0.94 | 0.95 | 0.000 |  |  |
| Number of previous live births | 1.45 | 1.42 | 1.47 | 0.000 |  |  |
| White | Reference | Reference | Reference | Reference |  |  |
| Mixed | 0.82 | 0.73 | 0.92 | 0.001 |  |  |
| Asian | 0.78 | 0.70 | 0.86 | 0.000 |  |  |
| Black | 0.66 | 0.59 | 0.74 | 0.000 |  |  |
| Other | 0.85 | 0.76 | 0.96 | 0.006 |  |  |
| Missing ethnicity | 0.80 | 0.77 | 0.85 | 0.000 |  |  |

| **Outcome: underweight** | | | | | | |
| --- | --- | --- | --- | --- | --- | --- |
| **Migration status** | **Odds ratio** | **95% LCI** | **95% UCI** | **P value** | **Number of observations** | **Pseudo R squared (Hosmer-Lemeshow)** |
| **Model 1** | | | | | | |
| Unlikely migrant (English as first language without complex social factors) | Reference | Reference | Reference | Reference | 496,267 | 0.009 |
| Probable migrant (English not their first language and complex social factors) | 2.07 | 1.92 | 2.22 | 0.000 |  |  |
| Possible migrant (only English not their first language) | 1.35 | 1.28 | 1.42 | 0.000 |  |  |
| Possible migrant (only complex social factors) | 2.58 | 2.44 | 2.71 | 0.000 |  |  |
| Missing migration status (first language or complex social factor data missing) | 1.11 | 1.07 | 1.16 | 0.000 |  |  |
| **Model 2b** | | | | | | |
| Unlikely migrant (English as first language without complex social factors) | Reference | Reference | Reference | Reference | 421,658 | 0.031 |
| Probable migrant (English not their first language and complex social factors) | 1.55 | 1.44 | 1.68 | 0.000 |  |  |
| Possible migrant (only English not their first language) | 1.26 | 1.19 | 1.33 | 0.000 |  |  |
| Possible migrant (only complex social factors) | 1.60 | 1.51 | 1.70 | 0.000 |  |  |
| Missing migration status (first language or complex social factor data missing) | 1.07 | 1.03 | 1.12 | 0.002 |  |  |
| Not most deprived | Reference | Reference | Reference | Reference |  |  |
| Most deprived | 1.08 | 1.03 | 1.13 | 0.002 |  |  |
| Mother's age at booking (years) | 0.93 | 0.92 | 0.93 | 0.000 |  |  |
| Number of previous live births | 0.97 | 0.95 | 0.99 | 0.002 |  |  |
| White | Reference | Reference | Reference | Reference |  |  |
| Mixed | 1.04 | 0.91 | 1.18 | 0.596 |  |  |
| Asian | 1.55 | 1.47 | 1.63 | 0.000 |  |  |
| Black | 0.72 | 0.64 | 0.80 | 0.000 |  |  |
| Other | 1.09 | 0.98 | 1.21 | 0.113 |  |  |
| Missing ethnicity | 1.14 | 1.08 | 1.20 | 0.000 |  |  |
| **Model 3b** | | | | | | |
| Unlikely migrant (English as first language without complex social factors) | Reference | Reference | Reference | Reference | 421,658 | 0.028 |
| Probable migrant (English not their first language and complex social factors) | 1.70 | 1.58 | 1.84 | 0.000 |  |  |
| Possible migrant (only English not their first language) | 1.38 | 1.30 | 1.45 | 0.000 |  |  |
| Possible migrant (only complex social factors) | 1.58 | 1.49 | 1.68 | 0.000 |  |  |
| Missing migration status (first language or complex social factor data missing) | 1.08 | 1.04 | 1.13 | 0.001 |  |  |
| Not most deprived | Reference | Reference | Reference | Reference |  |  |
| Most deprived | 1.09 | 1.04 | 1.14 | 0.001 |  |  |
| Mother's age at booking (years) | 0.93 | 0.93 | 0.93 | 0.000 |  |  |
| Number of previous live births | 0.96 | 0.95 | 0.98 | 0.000 |  |  |
| **Model 4b** | | | | | | |
| Unlikely migrant (English as first language without complex social factors) | Reference | Reference | Reference | Reference | 286,331 | 0.033 |
| Probable migrant (English not their first language and complex social factors) | 1.54 | 1.41 | 1.68 | 0.000 |  |  |
| Possible migrant (only English not their first language) | 1.22 | 1.14 | 1.30 | 0.000 |  |  |
| Possible migrant (only complex social factors) | 1.61 | 1.50 | 1.72 | 0.000 |  |  |
| Not most deprived | Reference | Reference | Reference | Reference |  |  |
| Most deprived | 1.06 | 1.00 | 1.12 | 0.065 |  |  |
| Mother's age at booking (years) | 0.93 | 0.92 | 0.93 | 0.000 |  |  |
| Number of previous live births | 0.98 | 0.95 | 1.00 | 0.037 |  |  |
| White | Reference | Reference | Reference | Reference |  |  |
| Mixed | 1.03 | 0.89 | 1.19 | 0.667 |  |  |
| Asian | 1.45 | 1.36 | 1.54 | 0.000 |  |  |
| Black | 0.68 | 0.60 | 0.78 | 0.000 |  |  |
| Other | 1.05 | 0.94 | 1.18 | 0.388 |  |  |
| **Model 5b** | | | | | | |
| Unlikely migrant (English as first language without complex social factors) | Reference | Reference | Reference | Reference | 421,658 | 0.031 |
| Probable migrant (English not their first language and complex social factors) | 1.55 | 1.44 | 1.68 | 0.000 |  |  |
| Possible migrant (only English not their first language) | 1.26 | 1.19 | 1.33 | 0.000 |  |  |
| Possible migrant (only complex social factors) | 1.60 | 1.51 | 1.70 | 0.000 |  |  |
| Missing migration status (first language or complex social factor data missing) | 1.07 | 1.03 | 1.12 | 0.002 |  |  |
| Not most deprived | Reference | Reference | Reference | Reference |  |  |
| Most deprived | 1.08 | 1.03 | 1.13 | 0.002 |  |  |
| Mother's age at booking (years) | 0.93 | 0.92 | 0.93 | 0.000 |  |  |
| Number of previous live births | 0.97 | 0.95 | 0.99 | 0.002 |  |  |
| White | Reference | Reference | Reference | Reference |  |  |
| Mixed | 1.04 | 0.91 | 1.18 | 0.596 |  |  |
| Asian | 1.55 | 1.47 | 1.63 | 0.000 |  |  |
| Black | 0.72 | 0.64 | 0.80 | 0.000 |  |  |
| Other | 1.09 | 0.98 | 1.21 | 0.113 |  |  |
| Missing ethnicity | 1.14 | 1.08 | 1.20 | 0.000 |  |  |

| **Outcome: overweight** | | | | | | |
| --- | --- | --- | --- | --- | --- | --- |
| **Migration status** | **Odds ratio** | **95% LCI** | **95% UCI** | **P value** | **Number of observations** | **Pseudo R squared (Hosmer-Lemeshow)** |
| **Model 1** | | | | | | |
| Unlikely migrant (English as first language without complex social factors) | Reference | Reference | Reference | Reference | 496,267 | 0 |
| Probable migrant (English not their first language and complex social factors) | 0.99 | 0.96 | 1.03 | 0.693 |  |  |
| Possible migrant (only English not their first language) | 1.02 | 1.00 | 1.04 | 0.123 |  |  |
| Possible migrant (only complex social factors) | 0.87 | 0.84 | 0.89 | 0.000 |  |  |
| Missing migration status (first language or complex social factor data missing) | 1.01 | 0.99 | 1.02 | 0.212 |  |  |
| **Model 2b** | | | | | | |
| Unlikely migrant (English as first language without complex social factors) | Reference | Reference | Reference | Reference | 421,658 | 0.003 |
| Probable migrant (English not their first language and complex social factors) | 0.95 | 0.92 | 0.99 | 0.007 |  |  |
| Possible migrant (only English not their first language) | 0.96 | 0.94 | 0.98 | 0.000 |  |  |
| Possible migrant (only complex social factors) | 0.94 | 0.91 | 0.96 | 0.000 |  |  |
| Missing migration status (first language or complex social factor data missing) | 1.01 | 1.00 | 1.03 | 0.162 |  |  |
| Not most deprived | Reference | Reference | Reference | Reference |  |  |
| Most deprived | 1.02 | 1.00 | 1.04 | 0.044 |  |  |
| Mother's age at booking (years) | 1.01 | 1.01 | 1.01 | 0.000 |  |  |
| Number of previous live births | 1.04 | 1.03 | 1.05 | 0.000 |  |  |
| White | Reference | Reference | Reference | Reference |  |  |
| Mixed | 1.09 | 1.04 | 1.15 | 0.001 |  |  |
| Asian | 1.26 | 1.24 | 1.29 | 0.000 |  |  |
| Black | 1.35 | 1.31 | 1.40 | 0.000 |  |  |
| Other | 1.12 | 1.07 | 1.16 | 0.000 |  |  |
| Missing ethnicity | 1.00 | 0.98 | 1.02 | 0.972 |  |  |
| **Model 3b** |  |  |  |  |  |  |
| Unlikely migrant (English as first language without complex social factors) | Reference | Reference | Reference | Reference | 421,658 | 0.002 |
| Probable migrant (English not their first language and complex social factors) | 1.01 | 0.98 | 1.05 | 0.525 |  |  |
| Possible migrant (only English not their first language) | 1.01 | 0.99 | 1.03 | 0.386 |  |  |
| Possible migrant (only complex social factors) | 0.93 | 0.91 | 0.96 | 0.000 |  |  |
| Missing migration status (first language or complex social factor data missing) | 1.02 | 1.00 | 1.04 | 0.041 |  |  |
| Not most deprived | Reference | Reference | Reference | Reference |  |  |
| Most deprived | 1.04 | 1.02 | 1.06 | 0.000 |  |  |
| Mother's age at booking (years) | 1.01 | 1.01 | 1.01 | 0.000 |  |  |
| Number of previous live births | 1.04 | 1.04 | 1.05 | 0.000 |  |  |
| **Model 4b** | | | | | | |
| Unlikely migrant (English as first language without complex social factors) | Reference | Reference | Reference | Reference | 286,331 | 0.003 |
| Probable migrant (English not their first language and complex social factors) | 0.93 | 0.89 | 0.97 | 0.001 |  |  |
| Possible migrant (only English not their first language) | 0.97 | 0.95 | 0.99 | 0.014 |  |  |
| Possible migrant (only complex social factors) | 0.93 | 0.90 | 0.96 | 0.000 |  |  |
| Not most deprived | Reference | Reference | Reference | Reference |  |  |
| Most deprived | 1.00 | 0.98 | 1.03 | 0.765 |  |  |
| Mother's age at booking (years) | 1.01 | 1.01 | 1.01 | 0.000 |  |  |
| Number of previous live births | 1.04 | 1.03 | 1.05 | 0.000 |  |  |
| White | Reference | Reference | Reference | Reference |  |  |
| Mixed | 1.08 | 1.02 | 1.15 | 0.007 |  |  |
| Asian | 1.28 | 1.25 | 1.31 | 0.000 |  |  |
| Black | 1.38 | 1.33 | 1.43 | 0.000 |  |  |
| Other | 1.10 | 1.05 | 1.15 | 0.000 |  |  |
| **Model 5b** | | | | | | |
| Unlikely migrant (English as first language without complex social factors) | Reference | Reference | Reference | Reference | 421,658 | 0.003 |
| Probable migrant (English not their first language and complex social factors) | 0.95 | 0.92 | 0.99 | 0.007 |  |  |
| Possible migrant (only English not their first language) | 0.96 | 0.94 | 0.98 | 0.000 |  |  |
| Possible migrant (only complex social factors) | 0.94 | 0.91 | 0.96 | 0.000 |  |  |
| Missing migration status (first language or complex social factor data missing) | 1.01 | 1.00 | 1.03 | 0.162 |  |  |
| Not most deprived | Reference | Reference | Reference | Reference |  |  |
| Most deprived | 1.02 | 1.00 | 1.04 | 0.044 |  |  |
| Mother's age at booking (years) | 1.01 | 1.01 | 1.01 | 0.000 |  |  |
| Number of previous live births | 1.04 | 1.03 | 1.05 | 0.000 |  |  |
| White | Reference | Reference | Reference | Reference |  |  |
| Mixed | 1.09 | 1.04 | 1.15 | 0.001 |  |  |
| Asian | 1.26 | 1.24 | 1.29 | 0.000 |  |  |
| Black | 1.35 | 1.31 | 1.40 | 0.000 |  |  |
| Other | 1.12 | 1.07 | 1.16 | 0.000 |  |  |
| Missing ethnicity | 1.00 | 0.98 | 1.02 | 0.972 |  |  |

| **Outcome: obese** | | | | | | |
| --- | --- | --- | --- | --- | --- | --- |
| **Migration status** | **Odds ratio** | **95% LCI** | **95% UCI** | **P value** | **Number of observations** | **Pseudo R squared (Hosmer-Lemeshow)** |
| **Model 1** | | | | | | |
| Unlikely migrant (English as first language without complex social factors) | Reference | Reference | Reference | Reference | 496,267 | 0.002 |
| Probable migrant (English not their first language and complex social factors) | 0.70 | 0.67 | 0.73 | 0.000 |  |  |
| Possible migrant (only English not their first language) | 0.70 | 0.68 | 0.72 | 0.000 |  |  |
| Possible migrant (only complex social factors) | 1.01 | 0.99 | 1.04 | 0.321 |  |  |
| Missing migration status (first language or complex social factor data missing) | 1.03 | 1.01 | 1.04 | 0.001 |  |  |
| **Model 2b** | | | | | | |
| Unlikely migrant (English as first language without complex social factors) | Reference | Reference | Reference | Reference | 421,658 | 0.02 |
| Probable migrant (English not their first language and complex social factors) | 0.66 | 0.63 | 0.69 | 0.000 |  |  |
| Possible migrant (only English not their first language) | 0.71 | 0.69 | 0.73 | 0.000 |  |  |
| Possible migrant (only complex social factors) | 0.91 | 0.88 | 0.94 | 0.000 |  |  |
| Missing migration status (first language or complex social factor data missing) | 1.00 | 0.99 | 1.02 | 0.636 |  |  |
| Not most deprived | Reference | Reference | Reference | Reference |  |  |
| Most deprived | 1.38 | 1.35 | 1.41 | 0.000 |  |  |
| Mother's age at booking (years) | 0.99 | 0.98 | 0.99 | 0.000 |  |  |
| Number of previous live births | 1.25 | 1.24 | 1.25 | 0.000 |  |  |
| White | Reference | Reference | Reference | Reference |  |  |
| Mixed | 1.04 | 0.99 | 1.10 | 0.125 |  |  |
| Asian | 0.80 | 0.77 | 0.82 | 0.000 |  |  |
| Black | 1.62 | 1.57 | 1.67 | 0.000 |  |  |
| Other | 0.81 | 0.78 | 0.85 | 0.000 |  |  |
| Missing ethnicity | 0.83 | 0.81 | 0.85 | 0.000 |  |  |
| **Model 3b** | | | | | | |
| Unlikely migrant (English as first language without complex social factors) | Reference | Reference | Reference | Reference | 421,658 | 0.017 |
| Probable migrant (English not their first language and complex social factors) | 0.63 | 0.60 | 0.65 | 0.000 |  |  |
| Possible migrant (only English not their first language) | 0.68 | 0.66 | 0.69 | 0.000 |  |  |
| Possible migrant (only complex social factors) | 0.92 | 0.89 | 0.95 | 0.000 |  |  |
| Missing migration status (first language or complex social factor data missing) | 1.00 | 0.98 | 1.02 | 0.921 |  |  |
| Not most deprived | Reference | Reference | Reference | Reference |  |  |
| Most deprived | 1.39 | 1.36 | 1.42 | 0.000 |  |  |
| Mother's age at booking (years) | 0.99 | 0.98 | 0.99 | 0.000 |  |  |
| Number of previous live births | 1.26 | 1.25 | 1.27 | 0.000 |  |  |
| **Model 4bb** | | | | | | |
| Unlikely migrant (English as first language without complex social factors) | Reference | Reference | Reference | Reference | 286,331 | 0.02 |
| Probable migrant (English not their first language and complex social factors) | 0.66 | 0.63 | 0.69 | 0.000 |  |  |
| Possible migrant (only English not their first language) | 0.71 | 0.69 | 0.73 | 0.000 |  |  |
| Possible migrant (only complex social factors) | 0.91 | 0.88 | 0.94 | 0.000 |  |  |
| Not most deprived | Reference | Reference | Reference | Reference |  |  |
| Most deprived | 1.41 | 1.38 | 1.45 | 0.000 |  |  |
| Mother's age at booking (years) | 0.99 | 0.98 | 0.99 | 0.000 |  |  |
| Number of previous live births | 1.25 | 1.24 | 1.26 | 0.000 |  |  |
| White | Reference | Reference | Reference | Reference |  |  |
| Mixed | 1.06 | 1.00 | 1.13 | 0.063 |  |  |
| Asian | 0.83 | 0.80 | 0.85 | 0.000 |  |  |
| Black | 1.65 | 1.59 | 1.72 | 0.000 |  |  |
| Other | 0.85 | 0.80 | 0.89 | 0.000 |  |  |
| **Model 5b** | | | | | | |
| Unlikely migrant (English as first language without complex social factors) | Reference | Reference | Reference | Reference | 404,079 | 0.021 |
| Probable migrant (English not their first language and complex social factors) | 0.67 | 0.64 | 0.70 | 0.000 |  |  |
| Possible migrant (only English not their first language) | 0.71 | 0.69 | 0.73 | 0.000 |  |  |
| Possible migrant (only complex social factors) | 1.13 | 1.09 | 1.17 | 0.000 |  |  |
| Missing migration status (first language or complex social factor data missing) | 1.01 | 0.99 | 1.03 | 0.315 |  |  |
| Not most deprived | Reference | Reference | Reference | Reference |  |  |
| Most deprived | 1.40 | 1.37 | 1.43 | 0.000 |  |  |
| Mother's age at booking (years) | 0.98 | 0.98 | 0.98 | 0.000 |  |  |
| Number of previous live births | 1.24 | 1.23 | 1.25 | 0.000 |  |  |
| White | Reference | Reference | Reference | Reference |  |  |
| Mixed | 1.04 | 0.99 | 1.10 | 0.126 |  |  |
| Asian | 0.79 | 0.77 | 0.81 | 0.000 |  |  |
| Black | 1.63 | 1.57 | 1.68 | 0.000 |  |  |
| Other | 0.81 | 0.77 | 0.85 | 0.000 |  |  |
| Missing ethnicity | 0.83 | 0.81 | 0.85 | 0.000 |  |  |

| **Outcome: at least one mental or physical condition** | | | | | | |
| --- | --- | --- | --- | --- | --- | --- |
| **Migration status** | **Odds ratio** | **95% LCI** | **95% UCI** | **P value** | **Number of observations** | **Pseudo R squared (Hosmer-Lemeshow)** |
| **Model 1** | | | | | | |
| Unlikely migrant (English as first language without complex social factors) | Reference | Reference | Reference | Reference | 652,880 | 0.049 |
| Probable migrant (English not their first language and complex social factors) | 0.44 | 0.43 | 0.45 | 0.000 |  |  |
| Possible migrant (only English not their first language) | 0.72 | 0.71 | 0.74 | 0.000 |  |  |
| Possible migrant (only complex social factors) | 0.98 | 0.96 | 1.00 | 0.075 |  |  |
| Missing migration status (first language or complex social factor data missing) | 0.27 | 0.27 | 0.28 | 0.000 |  |  |
| **Model 2a** | | | | | | |
| Unlikely migrant (English as first language without complex social factors) | Reference | Reference | Reference | Reference | 652,871 | 0.059 |
| Probable migrant (English not their first language and complex social factors) | 0.54 | 0.52 | 0.56 | 0.000 |  |  |
| Possible migrant (only English not their first language) | 0.83 | 0.82 | 0.85 | 0.000 |  |  |
| Possible migrant (only complex social factors) | 1.06 | 1.03 | 1.08 | 0.000 |  |  |
| Missing migration status (first language or complex social factor data missing) | 0.28 | 0.28 | 0.28 | 0.000 |  |  |
| Not most deprived | Reference | Reference | Reference | Reference |  |  |
| Most deprived | 1.24 | 1.22 | 1.26 | 0.000 |  |  |
| Mother's age at booking (years) | 1.02 | 1.02 | 1.02 | 0.000 |  |  |
| White | Reference | Reference | Reference | Reference |  |  |
| Mixed | 0.88 | 0.85 | 0.92 | 0.000 |  |  |
| Asian | 0.53 | 0.52 | 0.54 | 0.000 |  |  |
| Black | 0.69 | 0.66 | 0.71 | 0.000 |  |  |
| Other | 0.52 | 0.50 | 0.54 | 0.000 |  |  |
| Missing ethnicity | 0.64 | 0.63 | 0.65 | 0.000 |  |  |
| **Model 3a** | | | | | | |
| Unlikely migrant (English as first language without complex social factors) | Reference | Reference | Reference | Reference | 652,871 | 0.051 |
| Probable migrant (English not their first language and complex social factors) | 0.44 | 0.43 | 0.46 | 0.000 |  |  |
| Possible migrant (only English not their first language) | 0.71 | 0.69 | 0.72 | 0.000 |  |  |
| Possible migrant (only complex social factors) | 1.07 | 1.04 | 1.09 | 0.000 |  |  |
| Missing migration status (first language or complex social factor data missing) | 0.27 | 0.27 | 0.28 | 0.000 |  |  |
| Not most deprived | Reference | Reference | Reference | Reference |  |  |
| Most deprived | 1.19 | 1.17 | 1.21 | 0.000 |  |  |
| Mother's age at booking (years) | 1.02 | 1.02 | 1.02 | 0.000 |  |  |
| **Model 4a** | | | | | | |
| Unlikely migrant (English as first language without complex social factors) | Reference | Reference | Reference | Reference | 373,367 | 0.016 |
| Probable migrant (English not their first language and complex social factors) | 0.54 | 0.52 | 0.56 | 0.000 |  |  |
| Possible migrant (only English not their first language) | 0.85 | 0.83 | 0.86 | 0.000 |  |  |
| Possible migrant (only complex social factors) | 1.07 | 1.04 | 1.09 | 0.000 |  |  |
| Not most deprived | Reference | Reference | Reference | Reference |  |  |
| Most deprived | 1.40 | 1.37 | 1.43 | 0.000 |  |  |
| Mother's age at booking (years) | 1.02 | 1.02 | 1.02 | 0.000 |  |  |
| White | Reference | Reference | Reference | Reference |  |  |
| Mixed | 0.87 | 0.83 | 0.91 | 0.000 |  |  |
| Asian | 0.55 | 0.54 | 0.57 | 0.000 |  |  |
| Black | 0.72 | 0.70 | 0.75 | 0.000 |  |  |
| Other | 0.53 | 0.51 | 0.55 | 0.000 |  |  |
| **Model 5a** | | | | | | |
| Unlikely migrant (English as first language without complex social factors) | Reference | Reference | Reference | Reference | 622,544 | 0.058 |
| Probable migrant (English not their first language and complex social factors) | 0.54 | 0.52 | 0.56 | 0.000 |  |  |
| Possible migrant (only English not their first language) | 0.83 | 0.82 | 0.85 | 0.000 |  |  |
| Possible migrant (only complex social factors) | 1.05 | 1.02 | 1.08 | 0.003 |  |  |
| Missing migration status (first language or complex social factor data missing) | 0.29 | 0.28 | 0.29 | 0.000 |  |  |
| Not most deprived | Reference | Reference | Reference | Reference |  |  |
| Most deprived | 1.26 | 1.24 | 1.28 | 0.000 |  |  |
| Mother's age at booking (years) | 1.02 | 1.02 | 1.02 | 0.000 |  |  |
| White | Reference | Reference | Reference | Reference |  |  |
| Mixed | 0.88 | 0.84 | 0.92 | 0.000 |  |  |
| Asian | 0.53 | 0.52 | 0.55 | 0.000 |  |  |
| Black | 0.69 | 0.66 | 0.71 | 0.000 |  |  |
| Other | 0.53 | 0.51 | 0.55 | 0.000 |  |  |
| Missing ethnicity | 0.64 | 0.63 | 0.65 | 0.000 |  |  |

| **Outcome: at least one mental health condition** | | | | | | |
| --- | --- | --- | --- | --- | --- | --- |
| **Migration status** | **Odds ratio** | **95% LCI** | **95% UCI** | **P value** | **Number of observations** | **Pseudo R squared (Hosmer-Lemeshow)** |
| **Model 1** | | | | | | |
| Unlikely migrant (English as first language without complex social factors) | Reference | Reference | Reference | Reference | 652,880 | 0.042 |
| Probable migrant (English not their first language and complex social factors) | 0.31 | 0.29 | 0.33 | 0.000 |  |  |
| Possible migrant (only English not their first language) | 0.42 | 0.41 | 0.44 | 0.000 |  |  |
| Possible migrant (only complex social factors) | 1.69 | 1.65 | 1.74 | 0.000 |  |  |
| Missing migration status (first language or complex social factor data missing) | 0.32 | 0.31 | 0.33 | 0.000 |  |  |
| **Model 2a** | | | | | | |
| Unlikely migrant (English as first language without complex social factors) | Reference | Reference | Reference | Reference | 652,871 | 0.06 |
| Probable migrant (English not their first language and complex social factors) | 0.36 | 0.34 | 0.39 | 0.000 |  |  |
| Possible migrant (only English not their first language) | 0.53 | 0.51 | 0.54 | 0.000 |  |  |
| Possible migrant (only complex social factors) | 1.41 | 1.37 | 1.46 | 0.000 |  |  |
| Missing migration status (first language or complex social factor data missing) | 0.32 | 0.31 | 0.33 | 0.000 |  |  |
| Not most deprived | Reference | Reference | Reference | Reference |  |  |
| Most deprived | 1.44 | 1.40 | 1.47 | 0.000 |  |  |
| Mother's age at booking (years) | 0.98 | 0.98 | 0.98 | 0.000 |  |  |
| White | Reference | Reference | Reference | Reference |  |  |
| Mixed | 0.90 | 0.85 | 0.96 | 0.001 |  |  |
| Asian | 0.31 | 0.29 | 0.32 | 0.000 |  |  |
| Black | 0.42 | 0.40 | 0.45 | 0.000 |  |  |
| Other | 0.35 | 0.33 | 0.38 | 0.000 |  |  |
| Missing ethnicity | 0.65 | 0.63 | 0.67 | 0.000 |  |  |
| **Model 3a** | | | | | | |
| Unlikely migrant (English as first language without complex social factors) | Reference | Reference | Reference | Reference | 652,871 | 0.046 |
| Probable migrant (English not their first language and complex social factors) | 0.27 | 0.26 | 0.29 | 0.000 |  |  |
| Possible migrant (only English not their first language) | 0.42 | 0.40 | 0.43 | 0.000 |  |  |
| Possible migrant (only complex social factors) | 1.42 | 1.38 | 1.46 | 0.000 |  |  |
| Missing migration status (first language or complex social factor data missing) | 0.31 | 0.30 | 0.32 | 0.000 |  |  |
| Not most deprived | Reference | Reference | Reference | Reference |  |  |
| Most deprived | 1.35 | 1.32 | 1.39 | 0.000 |  |  |
| Mother's age at booking (years) | 0.98 | 0.98 | 0.98 | 0.000 |  |  |
| **Model 4a** | | | | | | |
| Unlikely migrant (English as first language without complex social factors) | Reference | Reference | Reference | Reference | 373,367 | 0.041 |
| Probable migrant (English not their first language and complex social factors) | 0.38 | 0.35 | 0.40 | 0.000 |  |  |
| Possible migrant (only English not their first language) | 0.55 | 0.52 | 0.57 | 0.000 |  |  |
| Possible migrant (only complex social factors) | 1.42 | 1.37 | 1.46 | 0.000 |  |  |
| Not most deprived | Reference | Reference | Reference | Reference |  |  |
| Most deprived | 1.60 | 1.56 | 1.65 | 0.000 |  |  |
| Mother's age at booking (years) | 0.98 | 0.98 | 0.98 | 0.000 |  |  |
| White | Reference | Reference | Reference | Reference |  |  |
| Mixed | 0.88 | 0.82 | 0.94 | 0.000 |  |  |
| Asian | 0.33 | 0.31 | 0.34 | 0.000 |  |  |
| Black | 0.45 | 0.42 | 0.48 | 0.000 |  |  |
| Other | 0.37 | 0.34 | 0.40 | 0.000 |  |  |
| **Model 5a** | | | | | | |
| Unlikely migrant (English as first language without complex social factors) | Reference | Reference | Reference | Reference | 622,544 | 0.057 |
| Probable migrant (English not their first language and complex social factors) | 0.33 | 0.31 | 0.35 | 0.000 |  |  |
| Possible migrant (only English not their first language) | 0.53 | 0.51 | 0.55 | 0.000 |  |  |
| Possible migrant (only complex social factors) | 1.48 | 1.42 | 1.53 | 0.000 |  |  |
| Missing migration status (first language or complex social factor data missing) | 0.32 | 0.32 | 0.33 | 0.000 |  |  |
| Not most deprived | Reference | Reference | Reference | Reference |  |  |
| Most deprived | 1.48 | 1.44 | 1.51 | 0.000 |  |  |
| Mother's age at booking (years) | 0.98 | 0.98 | 0.98 | 0.000 |  |  |
| White | Reference | Reference | Reference | Reference |  |  |
| Mixed | 0.90 | 0.84 | 0.96 | 0.001 |  |  |
| Asian | 0.31 | 0.30 | 0.32 | 0.000 |  |  |
| Black | 0.42 | 0.39 | 0.44 | 0.000 |  |  |
| Other | 0.35 | 0.33 | 0.38 | 0.000 |  |  |
| Missing ethnicity | 0.65 | 0.63 | 0.67 | 0.000 |  |  |

| **Outcome: at least one physical health condition** | | | | | | |
| --- | --- | --- | --- | --- | --- | --- |
| **Migration status** | **Odds ratio** | **95% LCI** | **95% UCI** | **P value** | **Number of observations** | **Pseudo R squared (Hosmer-Lemeshow)** |
| **Model 1** | | | | | | |
| Unlikely migrant (English as first language without complex social factors) | Reference | Reference | Reference | Reference | 652,880 | 0.042 |
| Probable migrant (English not their first language and complex social factors) | 0.50 | 0.48 | 0.52 | 0.000 |  |  |
| Possible migrant (only English not their first language) | 0.85 | 0.83 | 0.86 | 0.000 |  |  |
| Possible migrant (only complex social factors) | 0.69 | 0.67 | 0.71 | 0.000 |  |  |
| Missing migration status (first language or complex social factor data missing) | 0.28 | 0.28 | 0.29 | 0.000 |  |  |
| **Model 2a** | | | | | | |
| Unlikely migrant (English as first language without complex social factors) | Reference | Reference | Reference | Reference | 652,871 | 0.053 |
| Probable migrant (English not their first language and complex social factors) | 0.62 | 0.60 | 0.64 | 0.000 |  |  |
| Possible migrant (only English not their first language) | 0.94 | 0.92 | 0.96 | 0.000 |  |  |
| Possible migrant (only complex social factors) | 0.82 | 0.80 | 0.84 | 0.000 |  |  |
| Missing migration status (first language or complex social factor data missing) | 0.29 | 0.29 | 0.30 | 0.000 |  |  |
| Not most deprived | Reference | Reference | Reference | Reference |  |  |
| Most deprived | 1.16 | 1.14 | 1.18 | 0.000 |  |  |
| Mother's age at booking (years) | 1.04 | 1.04 | 1.04 | 0.000 |  |  |
| White | Reference | Reference | Reference | Reference |  |  |
| Mixed | 0.90 | 0.86 | 0.94 | 0.000 |  |  |
| Asian | 0.62 | 0.61 | 0.64 | 0.000 |  |  |
| Black | 0.81 | 0.78 | 0.84 | 0.000 |  |  |
| Other | 0.60 | 0.57 | 0.62 | 0.000 |  |  |
| Missing ethnicity | 0.66 | 0.65 | 0.67 | 0.000 |  |  |
| **Model 3a** | | | | | | |
| Unlikely migrant (English as first language without complex social factors) | Reference | Reference | Reference | Reference | 652,871 | 0.048 |
| Probable migrant (English not their first language and complex social factors) | 0.53 | 0.51 | 0.55 | 0.000 |  |  |
| Possible migrant (only English not their first language) | 0.83 | 0.81 | 0.84 | 0.000 |  |  |
| Possible migrant (only complex social factors) | 0.83 | 0.80 | 0.85 | 0.000 |  |  |
| Missing migration status (first language or complex social factor data missing) | 0.28 | 0.28 | 0.29 | 0.000 |  |  |
| Not most deprived | Reference | Reference | Reference | Reference |  |  |
| Most deprived | 1.13 | 1.11 | 1.15 | 0.000 |  |  |
| Mother's age at booking (years) | 1.04 | 1.03 | 1.04 | 0.000 |  |  |
| **Model 4a** | | | | | | |
| Unlikely migrant (English as first language without complex social factors) | Reference | Reference | Reference | Reference | 355,517 | 0.013 |
| Probable migrant (English not their first language and complex social factors) | 0.61 | 0.58 | 0.64 | 0.000 |  |  |
| Possible migrant (only English not their first language) | 0.95 | 0.93 | 0.97 | 0.000 |  |  |
| Possible migrant (only complex social factors) | 0.86 | 0.83 | 0.90 | 0.000 |  |  |
| Not most deprived | Reference | Reference | Reference | Reference |  |  |
| Most deprived | 1.28 | 1.26 | 1.31 | 0.000 |  |  |
| Mother's age at booking (years) | 1.04 | 1.04 | 1.04 | 0.000 |  |  |
| White | Reference | Reference | Reference | Reference |  |  |
| Mixed | 0.89 | 0.84 | 0.94 | 0.000 |  |  |
| Asian | 0.65 | 0.64 | 0.67 | 0.000 |  |  |
| Black | 0.85 | 0.82 | 0.88 | 0.000 |  |  |
| Other | 0.61 | 0.58 | 0.64 | 0.000 |  |  |
| **Model 5a** | | | | | | |
| Unlikely migrant (English as first language without complex social factors) | Reference | Reference | Reference | Reference | 622,544 | 0.051 |
| Probable migrant (English not their first language and complex social factors) | 0.62 | 0.60 | 0.64 | 0.000 |  |  |
| Possible migrant (only English not their first language) | 0.94 | 0.92 | 0.96 | 0.000 |  |  |
| Possible migrant (only complex social factors) | 0.86 | 0.83 | 0.89 | 0.000 |  |  |
| Missing migration status (first language or complex social factor data missing) | 0.30 | 0.29 | 0.30 | 0.000 |  |  |
| Not most deprived | Reference | Reference | Reference | Reference |  |  |
| Most deprived | 1.17 | 1.14 | 1.19 | 0.000 |  |  |
| Mother's age at booking (years) | 1.04 | 1.03 | 1.04 | 0.000 |  |  |
| White | Reference | Reference | Reference | Reference |  |  |
| Mixed | 0.90 | 0.85 | 0.94 | 0.000 |  |  |
| Asian | 0.63 | 0.61 | 0.64 | 0.000 |  |  |
| Black | 0.81 | 0.78 | 0.83 | 0.000 |  |  |
| Other | 0.60 | 0.57 | 0.62 | 0.000 |  |  |
| Missing ethnicity | 0.66 | 0.64 | 0.67 | 0.000 |  |  |

| **Outcome: diabetes** | | | | | | |
| --- | --- | --- | --- | --- | --- | --- |
| **Migration status** | **Odds ratio** | **95% LCI** | **95% UCI** | **P value** | **Number of observations** | **Pseudo R squared (Hosmer-Lemeshow)** |
| **Model 1** |  |  |  |  |  |  |
| Unlikely migrant (English as first language without complex social factors) | Reference | Reference | Reference | Reference | 652,880 | 0.016 |
| Probable migrant (English not their first language and complex social factors) | 1.46 | 1.31 | 1.61 | 0.000 |  |  |
| Possible migrant (only English not their first language) | 1.43 | 1.33 | 1.53 | 0.000 |  |  |
| Possible migrant (only complex social factors) | 0.93 | 0.83 | 1.03 | 0.174 |  |  |
| Missing migration status (first language or complex social factor data missing) | 0.41 | 0.39 | 0.44 | 0.000 |  |  |
| **Model 2a** | | | | | | |
| Unlikely migrant (English as first language without complex social factors) | Reference | Reference | Reference | Reference | 652,871 | 0.034 |
| Probable migrant (English not their first language and complex social factors) | 1.30 | 1.17 | 1.45 | 0.000 |  |  |
| Possible migrant (only English not their first language) | 1.18 | 1.10 | 1.27 | 0.000 |  |  |
| Possible migrant (only complex social factors) | 1.21 | 1.09 | 1.35 | 0.000 |  |  |
| Missing migration status (first language or complex social factor data missing) | 0.41 | 0.39 | 0.44 | 0.000 |  |  |
| Not most deprived | Reference | Reference | Reference | Reference |  |  |
| Most deprived | 1.68 | 1.57 | 1.78 | 0.000 |  |  |
| Mother's age at booking (years) | 1.06 | 1.06 | 1.07 | 0.000 |  |  |
| White | Reference | Reference | Reference | Reference |  |  |
| Mixed | 1.13 | 0.93 | 1.35 | 0.197 |  |  |
| Asian | 1.88 | 1.75 | 2.01 | 0.000 |  |  |
| Black | 1.40 | 1.26 | 1.56 | 0.000 |  |  |
| Other | 0.90 | 0.78 | 1.04 | 0.172 |  |  |
| Missing ethnicity | 0.94 | 0.87 | 1.02 | 0.125 |  |  |
| **Model 3a** | | | | | | |
| Unlikely migrant (English as first language without complex social factors) | Reference | Reference | Reference | Reference | 652,871 | 0.029 |
| Probable migrant (English not their first language and complex social factors) | 1.46 | 1.31 | 1.62 | 0.000 |  |  |
| Possible migrant (only English not their first language) | 1.33 | 1.24 | 1.42 | 0.000 |  |  |
| Possible migrant (only complex social factors) | 1.20 | 1.08 | 1.34 | 0.001 |  |  |
| Missing migration status (first language or complex social factor data missing) | 0.42 | 0.39 | 0.45 | 0.000 |  |  |
| Not most deprived | Reference | Reference | Reference | Reference |  |  |
| Most deprived | 1.75 | 1.65 | 1.87 | 0.000 |  |  |
| Mother's age at booking (years) | 1.07 | 1.06 | 1.07 | 0.000 |  |  |
| **Model 4a** | | | | | | |
| Unlikely migrant (English as first language without complex social factors) | Reference | Reference | Reference | Reference | 373,367 | 0.025 |
| Probable migrant (English not their first language and complex social factors) | 1.18 | 1.05 | 1.33 | 0.006 |  |  |
| Possible migrant (only English not their first language) | 1.13 | 1.04 | 1.22 | 0.003 |  |  |
| Possible migrant (only complex social factors) | 1.20 | 1.07 | 1.34 | 0.001 |  |  |
| Not most deprived | Reference | Reference | Reference | Reference |  |  |
| Most deprived | 1.83 | 1.70 | 1.97 | 0.000 |  |  |
| Mother's age at booking (years) | 1.06 | 1.06 | 1.07 | 0.000 |  |  |
| White | Reference | Reference | Reference | Reference |  |  |
| Mixed | 1.15 | 0.93 | 1.39 | 0.185 |  |  |
| Asian | 1.96 | 1.82 | 2.11 | 0.000 |  |  |
| Black | 1.52 | 1.35 | 1.70 | 0.000 |  |  |
| Other | 0.94 | 0.80 | 1.09 | 0.421 |  |  |
| **Model 5a** | | | | | | |
| Unlikely migrant (English as first language without complex social factors) | Reference | Reference | Reference | Reference | 622,544 | 0.051 |
| Probable migrant (English not their first language and complex social factors) | 1.33 | 1.20 | 1.48 | 0.000 |  |  |
| Possible migrant (only English not their first language) | 1.18 | 1.10 | 1.27 | 0.000 |  |  |
| Possible migrant (only complex social factors) | 1.21 | 1.06 | 1.37 | 0.003 |  |  |
| Missing migration status (first language or complex social factor data missing) | 0.42 | 0.39 | 0.45 | 0.000 |  |  |
| Not most deprived | Reference | Reference | Reference | Reference |  |  |
| Most deprived | 1.69 | 1.58 | 1.80 | 0.000 |  |  |
| Mother's age at booking (years) | 1.06 | 1.06 | 1.07 | 0.000 |  |  |
| White | Reference | Reference | Reference | Reference |  |  |
| Mixed | 1.15 | 0.95 | 1.38 | 0.149 |  |  |
| Asian | 1.89 | 1.76 | 2.02 | 0.000 |  |  |
| Black | 1.40 | 1.26 | 1.56 | 0.000 |  |  |
| Other | 0.90 | 0.77 | 1.04 | 0.175 |  |  |
| Missing ethnicity | 0.95 | 0.88 | 1.03 | 0.203 |  |  |

| **Outcome: hypertension** | | | | | | |
| --- | --- | --- | --- | --- | --- | --- |
| **Migration status** | **Odds ratio** | **95% LCI** | **95% UCI** | **P value** | **Number of observations** | **Pseudo R squared (Hosmer-Lemeshow)** |
| **Model 1** |  |  |  |  |  |  |
| Unlikely migrant (English as first language without complex social factors) | Reference | Reference | Reference | Reference | 652,880 | 0.02 |
| Probable migrant (English not their first language and complex social factors) | 0.60 | 0.52 | 0.68 | 0.000 |  |  |
| Possible migrant (only English not their first language) | 0.89 | 0.82 | 0.95 | 0.001 |  |  |
| Possible migrant (only complex social factors) | 0.62 | 0.55 | 0.69 | 0.000 |  |  |
| Missing migration status (first language or complex social factor data missing) | 0.30 | 0.28 | 0.32 | 0.000 |  |  |
| **Model 2a** | | | | | | |
| Unlikely migrant (English as first language without complex social factors) | Reference | Reference | Reference | Reference | 652,871 | 0.039 |
| Probable migrant (English not their first language and complex social factors) | 0.67 | 0.58 | 0.77 | 0.000 |  |  |
| Possible migrant (only English not their first language) | 0.89 | 0.82 | 0.95 | 0.002 |  |  |
| Possible migrant (only complex social factors) | 0.83 | 0.74 | 0.93 | 0.001 |  |  |
| Missing migration status (first language or complex social factor data missing) | 0.31 | 0.29 | 0.33 | 0.000 |  |  |
| Not most deprived | Reference | Reference | Reference | Reference |  |  |
| Most deprived | 1.44 | 1.34 | 1.54 | 0.000 |  |  |
| Mother's age at booking (years) | 1.07 | 1.07 | 1.08 | 0.000 |  |  |
| White | Reference | Reference | Reference | Reference |  |  |
| Mixed | 1.02 | 0.86 | 1.21 | 0.792 |  |  |
| Asian | 0.85 | 0.78 | 0.92 | 0.000 |  |  |
| Black | 1.68 | 1.53 | 1.84 | 0.000 |  |  |
| Other | 0.72 | 0.62 | 0.84 | 0.000 |  |  |
| Missing ethnicity | 0.57 | 0.52 | 0.62 | 0.000 |  |  |
| **Model 3a** | | | | | | |
| Unlikely migrant (English as first language without complex social factors) | Reference | Reference | Reference | Reference | 652,871 | 0.034 |
| Probable migrant (English not their first language and complex social factors) | 0.63 | 0.55 | 0.72 | 0.000 |  |  |
| Possible migrant (only English not their first language) | 0.84 | 0.78 | 0.90 | 0.000 |  |  |
| Possible migrant (only complex social factors) | 0.85 | 0.75 | 0.95 | 0.005 |  |  |
| Missing migration status (first language or complex social factor data missing) | 0.30 | 0.28 | 0.32 | 0.000 |  |  |
| Not most deprived | Reference | Reference | Reference | Reference |  |  |
| Most deprived | 1.47 | 1.38 | 1.57 | 0.000 |  |  |
| Mother's age at booking (years) | 1.07 | 1.07 | 1.08 | 0.000 |  |  |
| **Model 4a** | | | | | | |
| Unlikely migrant (English as first language without complex social factors) | Reference | Reference | Reference | Reference | 373,367 | 0.018 |
| Probable migrant (English not their first language and complex social factors) | 0.62 | 0.53 | 0.72 | 0.000 |  |  |
| Possible migrant (only English not their first language) | 0.89 | 0.82 | 0.96 | 0.004 |  |  |
| Possible migrant (only complex social factors) | 0.82 | 0.73 | 0.92 | 0.001 |  |  |
| Not most deprived | Reference | Reference | Reference | Reference |  |  |
| Most deprived | 1.53 | 1.41 | 1.64 | 0.000 |  |  |
| Mother's age at booking (years) | 1.07 | 1.06 | 1.07 | 0.000 |  |  |
| White | Reference | Reference | Reference | Reference |  |  |
| Mixed | 0.91 | 0.74 | 1.10 | 0.341 |  |  |
| Asian | 0.87 | 0.80 | 0.96 | 0.003 |  |  |
| Black | 1.70 | 1.54 | 1.87 | 0.000 |  |  |
| Other | 0.73 | 0.62 | 0.86 | 0.000 |  |  |
| **Model 5a** | | | | | | |
| Unlikely migrant (English as first language without complex social factors) | Reference | Reference | Reference | Reference | 622,544 | 0.036 |
| Probable migrant (English not their first language and complex social factors) | 0.69 | 0.59 | 0.79 | 0.000 |  |  |
| Possible migrant (only English not their first language) | 0.89 | 0.82 | 0.96 | 0.002 |  |  |
| Possible migrant (only complex social factors) | 0.88 | 0.77 | 1.00 | 0.051 |  |  |
| Missing migration status (first language or complex social factor data missing) | 0.32 | 0.30 | 0.34 | 0.000 |  |  |
| Not most deprived | Reference | Reference | Reference | Reference |  |  |
| Most deprived | 1.45 | 1.35 | 1.55 | 0.000 |  |  |
| Mother's age at booking (years) | 1.07 | 1.06 | 1.07 | 0.000 |  |  |
| White | Reference | Reference | Reference | Reference |  |  |
| Mixed | 1.02 | 0.85 | 1.21 | 0.847 |  |  |
| Asian | 0.85 | 0.78 | 0.92 | 0.000 |  |  |
| Black | 1.69 | 1.54 | 1.85 | 0.000 |  |  |
| Other | 0.70 | 0.60 | 0.82 | 0.000 |  |  |
| Missing ethnicity | 0.57 | 0.52 | 0.62 | 0.000 |  |  |

| **Outcome: cardiac disease** | | | | | | |
| --- | --- | --- | --- | --- | --- | --- |
| **Migration status** | **Odds ratio** | **95% LCI** | **95% UCI** | **P value** | **Number of observations** | **Pseudo R squared (Hosmer-Lemeshow)** |
| **Model 1** | | | | | | |
| Unlikely migrant (English as first language without complex social factors) | Reference | Reference | Reference | Reference | 652,880 | 0.012 |
| Probable migrant (English not their first language and complex social factors) | 0.50 | 0.42 | 0.59 | 0.000 |  |  |
| Possible migrant (only English not their first language) | 0.82 | 0.75 | 0.90 | 0.000 |  |  |
| Possible migrant (only complex social factors) | 0.84 | 0.75 | 0.94 | 0.003 |  |  |
| Missing migration status (first language or complex social factor data missing) | 0.41 | 0.38 | 0.44 | 0.000 |  |  |
| **Model 2a** | | | | | | |
| Unlikely migrant (English as first language without complex social factors) | Reference | Reference | Reference | Reference | 652,871 | 0.019 |
| Probable migrant (English not their first language and complex social factors) | 0.61 | 0.51 | 0.72 | 0.000 |  |  |
| Possible migrant (only English not their first language) | 1.00 | 0.91 | 1.09 | 0.994 |  |  |
| Possible migrant (only complex social factors) | 0.79 | 0.70 | 0.89 | 0.000 |  |  |
| Missing migration status (first language or complex social factor data missing) | 0.42 | 0.39 | 0.45 | 0.000 |  |  |
| Not most deprived | Reference | Reference | Reference | Reference |  |  |
| Most deprived | 1.14 | 1.05 | 1.24 | 0.001 |  |  |
| Mother's age at booking (years) | 1.00 | 0.99 | 1.00 | 0.227 |  |  |
| White | Reference | Reference | Reference | Reference |  |  |
| Mixed | 0.71 | 0.56 | 0.88 | 0.002 |  |  |
| Asian | 0.35 | 0.30 | 0.40 | 0.000 |  |  |
| Black | 0.56 | 0.47 | 0.66 | 0.000 |  |  |
| Other | 0.60 | 0.49 | 0.71 | 0.000 |  |  |
| Missing ethnicity | 0.63 | 0.58 | 0.69 | 0.000 |  |  |
| **Model 3a** | | | | | | |
| Unlikely migrant (English as first language without complex social factors) | Reference | Reference | Reference | Reference | 652,871 | 0.012 |
| Probable migrant (English not their first language and complex social factors) | 0.48 | 0.40 | 0.57 | 0.000 |  |  |
| Possible migrant (only English not their first language) | 0.82 | 0.75 | 0.89 | 0.000 |  |  |
| Possible migrant (only complex social factors) | 0.80 | 0.70 | 0.89 | 0.000 |  |  |
| Missing migration status (first language or complex social factor data missing) | 0.40 | 0.37 | 0.43 | 0.000 |  |  |
| Not most deprived | Reference | Reference | Reference | Reference |  |  |
| Most deprived | 1.08 | 1.00 | 1.17 | 0.052 |  |  |
| Mother's age at booking (years) | 0.99 | 0.99 | 1.00 | 0.005 |  |  |
| **Model 4a** | | | | | | |
| Unlikely migrant (English as first language without complex social factors) | Reference | Reference | Reference | Reference | 373,367 | 0.009 |
| Probable migrant (English not their first language and complex social factors) | 0.59 | 0.48 | 0.71 | 0.000 |  |  |
| Possible migrant (only English not their first language) | 1.00 | 0.90 | 1.10 | 0.956 |  |  |
| Possible migrant (only complex social factors) | 0.77 | 0.68 | 0.87 | 0.000 |  |  |
| Not most deprived | Reference | Reference | Reference | Reference |  |  |
| Most deprived | 1.29 | 1.17 | 1.41 | 0.000 |  |  |
| Mother's age at booking (years) | 1.00 | 0.99 | 1.00 | 0.198 |  |  |
| White | Reference | Reference | Reference | Reference |  |  |
| Mixed | 0.75 | 0.59 | 0.95 | 0.020 |  |  |
| Asian | 0.37 | 0.32 | 0.43 | 0.000 |  |  |
| Black | 0.63 | 0.52 | 0.75 | 0.000 |  |  |
| Other | 0.64 | 0.52 | 0.77 | 0.000 |  |  |
| **Model 5a** | | | | | | |
| Unlikely migrant (English as first language without complex social factors) | Reference | Reference | Reference | Reference | 622,544 | 0.019 |
| Probable migrant (English not their first language and complex social factors) | 0.59 | 0.48 | 0.71 | 0.000 |  |  |
| Possible migrant (only English not their first language) | 1.00 | 0.91 | 1.10 | 0.957 |  |  |
| Possible migrant (only complex social factors) | 0.75 | 0.64 | 0.88 | 0.000 |  |  |
| Missing migration status (first language or complex social factor data missing) | 0.43 | 0.40 | 0.46 | 0.000 |  |  |
| Not most deprived | Reference | Reference | Reference | Reference |  |  |
| Most deprived | 1.16 | 1.06 | 1.25 | 0.001 |  |  |
| Mother's age at booking (years) | 1.00 | 0.99 | 1.00 | 0.093 |  |  |
| White | Reference | Reference | Reference | Reference |  |  |
| Mixed | 0.72 | 0.57 | 0.90 | 0.005 |  |  |
| Asian | 0.34 | 0.30 | 0.39 | 0.000 |  |  |
| Black | 0.53 | 0.44 | 0.63 | 0.000 |  |  |
| Other | 0.61 | 0.50 | 0.73 | 0.000 |  |  |
| Missing ethnicity | 0.63 | 0.57 | 0.69 | 0.000 |  |  |

| **Outcome: thromboembolic condition** | | | | | | |
| --- | --- | --- | --- | --- | --- | --- |
| **Migration status** | **Odds ratio** | **95% LCI** | **95% UCI** | **P value** | **Number of observations** | **Pseudo R squared (Hosmer-Lemeshow)** |
| **Model 1** | | | | | | |
| Unlikely migrant (English as first language without complex social factors) | Reference | Reference | Reference | Reference | 652,880 | 0.017 |
| Probable migrant (English not their first language and complex social factors) | 0.59 | 0.49 | 0.70 | 0.000 |  |  |
| Possible migrant (only English not their first language) | 0.95 | 0.87 | 1.05 | 0.333 |  |  |
| Possible migrant (only complex social factors) | 0.69 | 0.60 | 0.79 | 0.000 |  |  |
| Missing migration status (first language or complex social factor data missing) | 0.31 | 0.29 | 0.34 | 0.000 |  |  |
| **Model 2a** | | | | | | |
| Unlikely migrant (English as first language without complex social factors) | Reference | Reference | Reference | Reference | 652,871 | 0.032 |
| Probable migrant (English not their first language and complex social factors) | 0.80 | 0.66 | 0.96 | 0.017 |  |  |
| Possible migrant (only English not their first language) | 1.12 | 1.02 | 1.24 | 0.019 |  |  |
| Possible migrant (only complex social factors) | 0.88 | 0.76 | 1.02 | 0.091 |  |  |
| Missing migration status (first language or complex social factor data missing) | 0.34 | 0.31 | 0.37 | 0.000 |  |  |
| Not most deprived | Reference | Reference | Reference | Reference |  |  |
| Most deprived | 1.36 | 1.24 | 1.49 | 0.000 |  |  |
| Mother's age at booking (years) | 1.06 | 1.05 | 1.07 | 0.000 |  |  |
| White | Reference | Reference | Reference | Reference |  |  |
| Mixed | 0.69 | 0.53 | 0.89 | 0.005 |  |  |
| Asian | 0.37 | 0.32 | 0.43 | 0.000 |  |  |
| Black | 0.55 | 0.45 | 0.65 | 0.000 |  |  |
| Other | 0.71 | 0.58 | 0.85 | 0.000 |  |  |
| Missing ethnicity | 0.49 | 0.44 | 0.55 | 0.000 |  |  |
| **Model 3a** | | | | | | |
| Unlikely migrant (English as first language without complex social factors) | Reference | Reference | Reference | Reference | 652,871 | 0.024 |
| Probable migrant (English not their first language and complex social factors) | 0.62 | 0.51 | 0.74 | 0.000 |  |  |
| Possible migrant (only English not their first language) | 0.92 | 0.83 | 1.01 | 0.074 |  |  |
| Possible migrant (only complex social factors) | 0.89 | 0.77 | 1.03 | 0.132 |  |  |
| Missing migration status (first language or complex social factor data missing) | 0.32 | 0.29 | 0.35 | 0.000 |  |  |
| Not most deprived | Reference | Reference | Reference | Reference |  |  |
| Most deprived | 1.28 | 1.17 | 1.40 | 0.000 |  |  |
| Mother's age at booking (years) | 1.06 | 1.05 | 1.06 | 0.000 |  |  |
| **Model 4a** | | | | | | |
| Unlikely migrant (English as first language without complex social factors) | Reference | Reference | Reference | Reference | 373,367 | 0.016 |
| Probable migrant (English not their first language and complex social factors) | 0.80 | 0.65 | 0.97 | 0.031 |  |  |
| Possible migrant (only English not their first language) | 1.14 | 1.03 | 1.27 | 0.012 |  |  |
| Possible migrant (only complex social factors) | 0.88 | 0.75 | 1.02 | 0.092 |  |  |
| Not most deprived | Reference | Reference | Reference | Reference |  |  |
| Most deprived | 1.42 | 1.28 | 1.57 | 0.000 |  |  |
| Mother's age at booking (years) | 1.06 | 1.05 | 1.07 | 0.000 |  |  |
| White | Reference | Reference | Reference | Reference |  |  |
| Mixed | 0.70 | 0.52 | 0.91 | 0.010 |  |  |
| Asian | 0.38 | 0.33 | 0.45 | 0.000 |  |  |
| Black | 0.54 | 0.44 | 0.65 | 0.000 |  |  |
| Other | 0.70 | 0.57 | 0.85 | 0.000 |  |  |
| **Model 5a** | | | | | | |
| Unlikely migrant (English as first language without complex social factors) | Reference | Reference | Reference | Reference | 622,544 | 0.03 |
| Probable migrant (English not their first language and complex social factors) | 0.81 | 0.66 | 0.98 | 0.031 |  |  |
| Possible migrant (only English not their first language) | 1.13 | 1.03 | 1.25 | 0.012 |  |  |
| Possible migrant (only complex social factors) | 0.91 | 0.77 | 1.08 | 0.304 |  |  |
| Missing migration status (first language or complex social factor data missing) | 0.35 | 0.31 | 0.38 | 0.000 |  |  |
| Not most deprived | Reference | Reference | Reference | Reference |  |  |
| Most deprived | 1.35 | 1.23 | 1.48 | 0.000 |  |  |
| Mother's age at booking (years) | 1.06 | 1.05 | 1.06 | 0.000 |  |  |
| White | Reference | Reference | Reference | Reference |  |  |
| Mixed | 0.69 | 0.53 | 0.89 | 0.006 |  |  |
| Asian | 0.37 | 0.32 | 0.43 | 0.000 |  |  |
| Black | 0.55 | 0.45 | 0.65 | 0.000 |  |  |
| Other | 0.71 | 0.58 | 0.85 | 0.000 |  |  |
| Missing ethnicity | 0.48 | 0.43 | 0.54 | 0.000 |  |  |

| **Outcome: renal disease** | | | | | | |
| --- | --- | --- | --- | --- | --- | --- |
| **Migration status** | **Odds ratio** | **95% LCI** | **95% UCI** | **P value** | **Number of observations** | **Pseudo R squared (Hosmer-Lemeshow)** |
| **Model 1** | | | | | | |
| Unlikely migrant (English as first language without complex social factors) | Reference | Reference | Reference | Reference | 652,880 | 0.004 |
| Probable migrant (English not their first language and complex social factors) | 0.79 | 0.68 | 0.92 | 0.002 |  |  |
| Possible migrant (only English not their first language) | 1.02 | 0.93 | 1.11 | 0.677 |  |  |
| Possible migrant (only complex social factors) | 0.87 | 0.77 | 0.98 | 0.022 |  |  |
| Missing migration status (first language or complex social factor data missing) | 0.61 | 0.57 | 0.65 | 0.000 |  |  |
| **Model 2a** |  |  |  |  |  |  |
| Unlikely migrant (English as first language without complex social factors) | Reference | Reference | Reference | Reference | 652,871 | 0.012 |
| Probable migrant (English not their first language and complex social factors) | 0.97 | 0.83 | 1.13 | 0.714 |  |  |
| Possible migrant (only English not their first language) | 1.25 | 1.14 | 1.36 | 0.000 |  |  |
| Possible migrant (only complex social factors) | 0.79 | 0.69 | 0.89 | 0.000 |  |  |
| Missing migration status (first language or complex social factor data missing) | 0.63 | 0.59 | 0.68 | 0.000 |  |  |
| Not most deprived | Reference | Reference | Reference | Reference |  |  |
| Most deprived | 0.83 | 0.76 | 0.91 | 0.000 |  |  |
| Mother's age at booking (years) | 0.98 | 0.98 | 0.99 | 0.000 |  |  |
| White | Reference | Reference | Reference | Reference |  |  |
| Mixed | 0.76 | 0.60 | 0.94 | 0.013 |  |  |
| Asian | 0.42 | 0.37 | 0.48 | 0.000 |  |  |
| Black | 0.37 | 0.30 | 0.46 | 0.000 |  |  |
| Other | 0.74 | 0.63 | 0.87 | 0.000 |  |  |
| Missing ethnicity | 0.58 | 0.53 | 0.64 | 0.000 |  |  |
| **Model 3a** | | | | | | |
| Unlikely migrant (English as first language without complex social factors) | Reference | Reference | Reference | Reference | 652,871 | 0.005 |
| Probable migrant (English not their first language and complex social factors) | 0.79 | 0.67 | 0.91 | 0.002 |  |  |
| Possible migrant (only English not their first language) | 1.04 | 0.95 | 1.14 | 0.345 |  |  |
| Possible migrant (only complex social factors) | 0.79 | 0.70 | 0.90 | 0.000 |  |  |
| Missing migration status (first language or complex social factor data missing) | 0.61 | 0.57 | 0.65 | 0.000 |  |  |
| Not most deprived | Reference | Reference | Reference | Reference |  |  |
| Most deprived | 0.79 | 0.72 | 0.86 | 0.000 |  |  |
| Mother's age at booking (years) | 0.98 | 0.98 | 0.99 | 0.000 |  |  |
| **Model 4a** |  |  |  |  |  |  |
| Unlikely migrant (English as first language without complex social factors) | Reference | Reference | Reference | Reference | 373,367 | 0.009 |
| Probable migrant (English not their first language and complex social factors) | 0.95 | 0.79 | 1.12 | 0.523 |  |  |
| Possible migrant (only English not their first language) | 1.25 | 1.13 | 1.37 | 0.000 |  |  |
| Possible migrant (only complex social factors) | 0.79 | 0.70 | 0.90 | 0.001 |  |  |
| Not most deprived | Reference | Reference | Reference | Reference |  |  |
| Most deprived | 0.80 | 0.72 | 0.89 | 0.000 |  |  |
| Mother's age at booking (years) | 0.99 | 0.98 | 0.99 | 0.000 |  |  |
| White | Reference | Reference | Reference | Reference |  |  |
| Mixed | 0.70 | 0.53 | 0.89 | 0.006 |  |  |
| Asian | 0.42 | 0.36 | 0.48 | 0.000 |  |  |
| Black | 0.34 | 0.27 | 0.43 | 0.000 |  |  |
| Other | 0.76 | 0.63 | 0.91 | 0.003 |  |  |
| **Model 5a** |  |  |  |  |  |  |
| Unlikely migrant (English as first language without complex social factors) | Reference | Reference | Reference | Reference | 622,544 | 0.013 |
| Probable migrant (English not their first language and complex social factors) | 1.01 | 0.86 | 1.19 | 0.867 |  |  |
| Possible migrant (only English not their first language) | 1.25 | 1.14 | 1.36 | 0.000 |  |  |
| Possible migrant (only complex social factors) | 0.83 | 0.70 | 0.97 | 0.026 |  |  |
| Missing migration status (first language or complex social factor data missing) | 0.64 | 0.59 | 0.68 | 0.000 |  |  |
| Not most deprived | Reference | Reference | Reference | Reference |  |  |
| Most deprived | 0.85 | 0.77 | 0.93 | 0.000 |  |  |
| Mother's age at booking (years) | 0.98 | 0.98 | 0.99 | 0.000 |  |  |
| White | Reference | Reference | Reference | Reference |  |  |
| Mixed | 0.76 | 0.60 | 0.94 | 0.015 |  |  |
| Asian | 0.42 | 0.37 | 0.48 | 0.000 |  |  |
| Black | 0.37 | 0.30 | 0.46 | 0.000 |  |  |
| Other | 0.73 | 0.62 | 0.87 | 0.000 |  |  |
| Missing ethnicity | 0.58 | 0.53 | 0.63 | 0.000 |  |  |

| **Outcome: hepatitis b** | | | | | | |
| --- | --- | --- | --- | --- | --- | --- |
| **Migration status** | **Odds ratio** | **95% LCI** | **95% UCI** | **P value** | **Number of observations** | **Pseudo R squared (Hosmer-Lemeshow)** |
| **Model 1** | | | | | | |
| Unlikely migrant (English as first language without complex social factors) | Reference | Reference | Reference | Reference | 652,880 | 0.065 |
| Probable migrant (English not their first language and complex social factors) | 7.39 | 6.12 | 8.90 | 0.000 |  |  |
| Possible migrant (only English not their first language) | 5.88 | 5.05 | 6.86 | 0.000 |  |  |
| Possible migrant (only complex social factors) | 1.00 | 0.69 | 1.40 | 0.997 |  |  |
| Missing migration status (first language or complex social factor data missing) | 0.56 | 0.45 | 0.69 | 0.000 |  |  |
| **Model 2a** | | | | | | |
| Unlikely migrant (English as first language without complex social factors) | Reference | Reference | Reference | Reference | 652,871 | 0.099 |
| Probable migrant (English not their first language and complex social factors) | 5.40 | 4.43 | 6.58 | 0.000 |  |  |
| Possible migrant (only English not their first language) | 4.44 | 3.78 | 5.21 | 0.000 |  |  |
| Possible migrant (only complex social factors) | 1.20 | 0.83 | 1.69 | 0.307 |  |  |
| Missing migration status (first language or complex social factor data missing) | 0.54 | 0.44 | 0.67 | 0.000 |  |  |
| Not most deprived | Reference | Reference | Reference | Reference |  |  |
| Most deprived | 1.28 | 1.10 | 1.49 | 0.001 |  |  |
| Mother's age at booking (years) | 1.04 | 1.03 | 1.06 | 0.000 |  |  |
| White | Reference | Reference | Reference | Reference |  |  |
| Mixed | 1.82 | 1.12 | 2.79 | 0.009 |  |  |
| Asian | 2.03 | 1.69 | 2.44 | 0.000 |  |  |
| Black | 6.82 | 5.70 | 8.14 | 0.000 |  |  |
| Other | 2.55 | 1.99 | 3.24 | 0.000 |  |  |
| Missing ethnicity | 1.31 | 1.06 | 1.60 | 0.010 |  |  |
| **Model 3a** | | | | | | |
| Unlikely migrant (English as first language without complex social factors) | Reference | Reference | Reference | Reference | 652,871 | 0.073 |
| Probable migrant (English not their first language and complex social factors) | 7.55 | 6.23 | 9.13 | 0.000 |  |  |
| Possible migrant (only English not their first language) | 5.56 | 4.77 | 6.49 | 0.000 |  |  |
| Possible migrant (only complex social factors) | 1.27 | 0.88 | 1.79 | 0.183 |  |  |
| Missing migration status (first language or complex social factor data missing) | 0.56 | 0.45 | 0.69 | 0.000 |  |  |
| Not most deprived | Reference | Reference | Reference | Reference |  |  |
| Most deprived | 1.54 | 1.32 | 1.79 | 0.000 |  |  |
| Mother's age at booking (years) | 1.06 | 1.05 | 1.07 | 0.000 |  |  |
| **Model 4a** | | | | | | |
| Unlikely migrant (English as first language without complex social factors) | Reference | Reference | Reference | Reference | 373,367 | 0.1 |
| Probable migrant (English not their first language and complex social factors) | 5.85 | 4.72 | 7.24 | 0.000 |  |  |
| Possible migrant (only English not their first language) | 4.87 | 4.10 | 5.80 | 0.000 |  |  |
| Possible migrant (only complex social factors) | 1.34 | 0.92 | 1.90 | 0.112 |  |  |
| Not most deprived | Reference | Reference | Reference | Reference |  |  |
| Most deprived | 1.26 | 1.05 | 1.49 | 0.010 |  |  |
| Mother's age at booking (years) | 1.04 | 1.03 | 1.06 | 0.000 |  |  |
| White | Reference | Reference | Reference | Reference |  |  |
| Mixed | 1.82 | 1.09 | 2.84 | 0.014 |  |  |
| Asian | 1.96 | 1.61 | 2.38 | 0.000 |  |  |
| Black | 6.46 | 5.33 | 7.80 | 0.000 |  |  |
| Other | 2.40 | 1.85 | 3.09 | 0.000 |  |  |
| **Model 5a** | | | | | | |
| Unlikely migrant (English as first language without complex social factors) | Reference | Reference | Reference | Reference | 622,544 | 0.098 |
| Probable migrant (English not their first language and complex social factors) | 5.37 | 4.38 | 6.56 | 0.000 |  |  |
| Possible migrant (only English not their first language) | 4.46 | 3.80 | 5.23 | 0.000 |  |  |
| Possible migrant (only complex social factors) | 1.28 | 0.83 | 1.87 | 0.231 |  |  |
| Missing migration status (first language or complex social factor data missing) | 0.56 | 0.45 | 0.69 | 0.000 |  |  |
| Not most deprived | Reference | Reference | Reference | Reference |  |  |
| Most deprived | 1.28 | 1.09 | 1.49 | 0.002 |  |  |
| Mother's age at booking (years) | 1.04 | 1.03 | 1.05 | 0.000 |  |  |
| White | Reference | Reference | Reference | Reference |  |  |
| Mixed | 1.90 | 1.17 | 2.91 | 0.006 |  |  |
| Asian | 2.03 | 1.68 | 2.44 | 0.000 |  |  |
| Black | 6.86 | 5.72 | 8.20 | 0.000 |  |  |
| Other | 2.47 | 1.91 | 3.16 | 0.000 |  |  |
| Missing ethnicity | 1.32 | 1.07 | 1.62 | 0.008 |  |  |

| **Outcome: cancer** | | | | | | |
| --- | --- | --- | --- | --- | --- | --- |
| **Migration status** | **Odds ratio** | **95% LCI** | **95% UCI** | **P value** | **Number of observations** | **Pseudo R squared (Hosmer-Lemeshow)** |
| **Model 1** | | | | | | |
| Unlikely migrant (English as first language without complex social factors) | Reference | Reference | Reference | Reference | 652,880 | 0.015 |
| Probable migrant (English not their first language and complex social factors) | 0.33 | 0.20 | 0.49 | 0.000 |  |  |
| Possible migrant (only English not their first language) | 0.62 | 0.50 | 0.76 | 0.000 |  |  |
| Possible migrant (only complex social factors) | 0.58 | 0.44 | 0.76 | 0.000 |  |  |
| Missing migration status | 0.31 | 0.26 | 0.37 | 0.000 |  |  |
| **Model 2a** | | | | | | |
| Unlikely migrant (English as first language without complex social factors) | Reference | Reference | Reference | Reference | 652,871 | 0.035 |
| Probable migrant (English not their first language and complex social factors) | 0.53 | 0.33 | 0.81 | 0.005 |  |  |
| Possible migrant (only English not their first language) | 0.77 | 0.62 | 0.95 | 0.016 |  |  |
| Possible migrant (only complex social factors) | 0.87 | 0.64 | 1.14 | 0.327 |  |  |
| Missing migration status | 0.35 | 0.29 | 0.41 | 0.000 |  |  |
| Not most deprived | Reference | Reference | Reference | Reference |  |  |
| Most deprived | 0.84 | 0.68 | 1.03 | 0.102 |  |  |
| Mother's age at booking (years) | 1.08 | 1.07 | 1.09 | 0.000 |  |  |
| White | Reference | Reference | Reference | Reference |  |  |
| Mixed | 0.66 | 0.39 | 1.05 | 0.104 |  |  |
| Asian | 0.40 | 0.30 | 0.52 | 0.000 |  |  |
| Black | 0.25 | 0.15 | 0.41 | 0.000 |  |  |
| Other | 0.47 | 0.29 | 0.71 | 0.001 |  |  |
| Missing ethnicity | 0.49 | 0.39 | 0.60 | 0.000 |  |  |
| **Model 3a** | | | | | | |
| Unlikely migrant (English as first language without complex social factors) | Reference | Reference | Reference | Reference | 652,781 | 0.026 |
| Probable migrant (English not their first language and complex social factors) | 0.39 | 0.24 | 0.59 | 0.000 |  |  |
| Possible migrant (only English not their first language) | 0.62 | 0.50 | 0.76 | 0.000 |  |  |
| Possible migrant (only complex social factors) | 0.87 | 0.64 | 1.14 | 0.327 |  |  |
| Missing migration status | 0.33 | 0.28 | 0.39 | 0.000 |  |  |
| Not most deprived | Reference | Reference | Reference | Reference |  |  |
| Most deprived | 0.77 | 0.62 | 0.95 | 0.016 |  |  |
| Mother's age at booking (years) | 1.07 | 1.06 | 1.09 | 0.000 |  |  |
| **Model 4a** | | | | | | |
| Unlikely migrant (English as first language without complex social factors) | Reference | Reference | Reference | Reference | 373,367 | 0.021 |
| Probable migrant (English not their first language and complex social factors) | 0.52 | 0.31 | 0.82 | 0.008 |  |  |
| Possible migrant (only English not their first language) | 0.80 | 0.63 | 0.99 | 0.048 |  |  |
| Possible migrant (only complex social factors) | 0.89 | 0.66 | 1.18 | 0.440 |  |  |
| Not most deprived | Reference | Reference | Reference | Reference |  |  |
| Most deprived | 0.94 | 0.74 | 1.18 | 0.610 |  |  |
| Mother's age at booking (years) | 1.08 | 1.06 | 1.09 | 0.000 |  |  |
| White | Reference | Reference | Reference | Reference |  |  |
| Mixed | 0.73 | 0.42 | 1.17 | 0.226 |  |  |
| Asian | 0.40 | 0.30 | 0.53 | 0.000 |  |  |
| Black | 0.29 | 0.17 | 0.47 | 0.000 |  |  |
| Other | 0.51 | 0.31 | 0.79 | 0.004 |  |  |
| **Model 5a** | | | | | | |
| Unlikely migrant (English as first language without complex social factors) | Reference | Reference | Reference | Reference | 622,544 | 0.033 |
| Probable migrant (English not their first language and complex social factors) | 0.48 | 0.28 | 0.75 | 0.003 |  |  |
| Possible migrant (only English not their first language) | 0.77 | 0.62 | 0.95 | 0.017 |  |  |
| Possible migrant (only complex social factors) | 0.88 | 0.62 | 1.20 | 0.442 |  |  |
| Missing migration status | 0.35 | 0.30 | 0.42 | 0.000 |  |  |
| Not most deprived | Reference | Reference | Reference | Reference |  |  |
| Most deprived | 0.84 | 0.67 | 1.04 | 0.114 |  |  |
| Mother's age at booking (years) | 1.08 | 1.07 | 1.09 | 0.000 |  |  |
| White | Reference | Reference | Reference | Reference |  |  |
| Mixed | 0.60 | 0.33 | 0.97 | 0.055 |  |  |
| Asian | 0.40 | 0.30 | 0.52 | 0.000 |  |  |
| Black | 0.26 | 0.15 | 0.41 | 0.000 |  |  |
| Other | 0.48 | 0.29 | 0.72 | 0.001 |  |  |
| Missing ethnicity | 0.48 | 0.39 | 0.60 | 0.000 |  |  |

| **Outcome: family history of inherited disease** | | | | | | |
| --- | --- | --- | --- | --- | --- | --- |
| **Migration status** | **Odds ratio** | **95% LCI** | **95% UCI** | **P value** | **Number of observations** | **Pseudo R squared (Hosmer-Lemeshow)** |
| **Model 1** | | | | | | |
| Unlikely migrant (English as first language without complex social factors) | Reference | Reference | Reference | Reference | 652,880 | 0.008 |
| Probable migrant (English not their first language and complex social factors) | 0.36 | 0.31 | 0.41 | 0.000 |  |  |
| Possible migrant (only English not their first language) | 0.71 | 0.67 | 0.75 | 0.000 |  |  |
| Possible migrant (only complex social factors) | 1.15 | 1.07 | 1.22 | 0.000 |  |  |
| Missing migration status (first language or complex social factor data missing) | 0.59 | 0.56 | 0.61 | 0.000 |  |  |
| **Model 2a** | | | | | | |
| Unlikely migrant (English as first language without complex social factors) | Reference | Reference | Reference | Reference | 652,871 | 0.015 |
| Probable migrant (English not their first language and complex social factors) | 0.44 | 0.39 | 0.51 | 0.000 |  |  |
| Possible migrant (only English not their first language) | 0.85 | 0.80 | 0.91 | 0.000 |  |  |
| Possible migrant (only complex social factors) | 1.08 | 1.00 | 1.15 | 0.039 |  |  |
| Missing migration status (first language or complex social factor data missing) | 0.61 | 0.58 | 0.64 | 0.000 |  |  |
| Not most deprived | Reference | Reference | Reference | Reference |  |  |
| Most deprived | 0.98 | 0.93 | 1.03 | 0.498 |  |  |
| Mother's age at booking (years) | 0.99 | 0.99 | 1.00 | 0.000 |  |  |
| White | Reference | Reference | Reference | Reference |  |  |
| Mixed | 0.83 | 0.73 | 0.95 | 0.006 |  |  |
| Asian | 0.53 | 0.49 | 0.57 | 0.000 |  |  |
| Black | 0.43 | 0.38 | 0.49 | 0.000 |  |  |
| Other | 0.41 | 0.36 | 0.48 | 0.000 |  |  |
| Missing ethnicity | 0.60 | 0.57 | 0.64 | 0.000 |  |  |
| **Model 3a** | | | | | | |
| Unlikely migrant (English as first language without complex social factors) | Reference | Reference | Reference | Reference | 652,871 | 0.008 |
| Probable migrant (English not their first language and complex social factors) | 0.35 | 0.31 | 0.40 | 0.000 |  |  |
| Possible migrant (only English not their first language) | 0.72 | 0.67 | 0.76 | 0.000 |  |  |
| Possible migrant (only complex social factors) | 1.08 | 1.01 | 1.15 | 0.033 |  |  |
| Missing migration status (first language or complex social factor data missing) | 0.58 | 0.56 | 0.61 | 0.000 |  |  |
| Not most deprived | Reference | Reference | Reference | Reference |  |  |
| Most deprived | 0.94 | 0.89 | 0.98 | 0.011 |  |  |
| Mother's age at booking (years) | 0.99 | 0.99 | 0.99 | 0.000 |  |  |
| **Model 4a** | | | | | | |
| Unlikely migrant (English as first language without complex social factors) | Reference | Reference | Reference | Reference | 373,367 | 0.009 |
| Probable migrant (English not their first language and complex social factors) | 0.46 | 0.40 | 0.53 | 0.000 |  |  |
| Possible migrant (only English not their first language) | 0.86 | 0.81 | 0.93 | 0.000 |  |  |
| Possible migrant (only complex social factors) | 1.08 | 1.01 | 1.17 | 0.030 |  |  |
| Not most deprived | Reference | Reference | Reference | Reference |  |  |
| Most deprived | 1.09 | 1.02 | 1.16 | 0.007 |  |  |
| Mother's age at booking (years) | 0.99 | 0.99 | 1.00 | 0.000 |  |  |
| White | Reference | Reference | Reference | Reference |  |  |
| Mixed | 0.84 | 0.72 | 0.97 | 0.020 |  |  |
| Asian | 0.61 | 0.56 | 0.66 | 0.000 |  |  |
| Black | 0.51 | 0.45 | 0.58 | 0.000 |  |  |
| Other | 0.45 | 0.38 | 0.52 | 0.000 |  |  |
| **Model 5a** | | | | | | |
| Unlikely migrant (English as first language without complex social factors) | Reference | Reference | Reference | Reference | 622,544 | 0.014 |
| Probable migrant (English not their first language and complex social factors) | 0.42 | 0.36 | 0.49 | 0.000 |  |  |
| Possible migrant (only English not their first language) | 0.85 | 0.80 | 0.91 | 0.000 |  |  |
| Possible migrant (only complex social factors) | 1.04 | 0.95 | 1.14 | 0.370 |  |  |
| Missing migration status (first language or complex social factor data missing) | 0.63 | 0.60 | 0.66 | 0.000 |  |  |
| Not most deprived | Reference | Reference | Reference | Reference |  |  |
| Most deprived | 0.99 | 0.94 | 1.04 | 0.658 |  |  |
| Mother's age at booking (years) | 0.99 | 0.99 | 1.00 | 0.000 |  |  |
| White | Reference | Reference | Reference | Reference |  |  |
| Mixed | 0.84 | 0.73 | 0.95 | 0.009 |  |  |
| Asian | 0.53 | 0.49 | 0.57 | 0.000 |  |  |
| Black | 0.43 | 0.38 | 0.49 | 0.000 |  |  |
| Other | 0.42 | 0.36 | 0.48 | 0.000 |  |  |
| Missing ethnicity | 0.61 | 0.57 | 0.64 | 0.000 |  |  |

| **Outcome: family history of diabetes** | | | | | | |
| --- | --- | --- | --- | --- | --- | --- |
| **Migration status** | **Odds ratio** | **95% LCI** | **95% UCI** | **P value** | **Number of observations** | **Pseudo R squared (Hosmer-Lemeshow)** |
| **Model 1** | | | | | | |
| Unlikely migrant (English as first language without complex social factors) | Reference | Reference | Reference | Reference | 652,880 | 0.052 |
| Probable migrant (English not their first language and complex social factors) | 1.13 | 1.10 | 1.17 | 0.000 |  |  |
| Possible migrant (only English not their first language) | 1.27 | 1.24 | 1.29 | 0.000 |  |  |
| Possible migrant (only complex social factors) | 0.97 | 0.94 | 0.99 | 0.007 |  |  |
| Missing migration status | 0.26 | 0.26 | 0.27 | 0.000 |  |  |
| **Model 2a** | | | | | | |
| Unlikely migrant (English as first language without complex social factors) | Reference | Reference | Reference | Reference | 652,871 | 0.072 |
| Probable migrant (English not their first language and complex social factors) | 0.89 | 0.86 | 0.92 | 0.000 |  |  |
| Possible migrant (only English not their first language) | 1.01 | 0.99 | 1.03 | 0.401 |  |  |
| Possible migrant (only complex social factors) | 1.00 | 0.97 | 1.02 | 0.888 |  |  |
| Missing migration status | 0.25 | 0.25 | 0.26 | 0.000 |  |  |
| Not most deprived | Reference | Reference | Reference | Reference |  |  |
| Most deprived | 1.28 | 1.26 | 1.31 | 0.000 |  |  |
| Mother's age at booking (years) | 1.00 | 1.00 | 1.01 | 0.000 |  |  |
| White | Reference | Reference | Reference | Reference |  |  |
| Mixed | 1.55 | 1.48 | 1.62 | 0.000 |  |  |
| Asian | 2.85 | 2.80 | 2.90 | 0.000 |  |  |
| Black | 1.41 | 1.37 | 1.45 | 0.000 |  |  |
| Other | 1.31 | 1.26 | 1.35 | 0.000 |  |  |
| Missing ethnicity | 0.99 | 0.97 | 1.00 | 0.123 |  |  |
| **Model 3a** | | | | | | |
| Unlikely migrant (English as first language without complex social factors) | Reference | Reference | Reference | Reference | 652,871 | 0.054 |
| Probable migrant (English not their first language and complex social factors) | 1.10 | 1.06 | 1.13 | 0.000 |  |  |
| Possible migrant (only English not their first language) | 1.23 | 1.21 | 1.26 | 0.000 |  |  |
| Possible migrant (only complex social factors) | 0.99 | 0.96 | 1.01 | 0.301 |  |  |
| Missing migration status | 0.26 | 0.26 | 0.27 | 0.000 |  |  |
| Not most deprived | Reference | Reference | Reference | Reference |  |  |
| Most deprived | 1.35 | 1.33 | 1.38 | 0.000 |  |  |
| Mother's age at booking (years) | 1.01 | 1.01 | 1.01 | 0.000 |  |  |
| **Model 4a** | | | | | | |
| Unlikely migrant (English as first language without complex social factors) | Reference | Reference | Reference | Reference | 373,367 | 0.032 |
| Probable migrant (English not their first language and complex social factors) | 0.83 | 0.80 | 0.86 | 0.000 |  |  |
| Possible migrant (only English not their first language) | 0.98 | 0.96 | 1.00 | 0.046 |  |  |
| Possible migrant (only complex social factors) | 1.02 | 0.99 | 1.05 | 0.129 |  |  |
| Not most deprived | Reference | Reference | Reference | Reference |  |  |
| Most deprived | 1.37 | 1.34 | 1.40 | 0.000 |  |  |
| Mother's age at booking (years) | 1.00 | 1.00 | 1.01 | 0.000 |  |  |
| White | Reference | Reference | Reference | Reference |  |  |
| Mixed | 1.58 | 1.51 | 1.66 | 0.000 |  |  |
| Asian | 3.21 | 3.15 | 3.28 | 0.000 |  |  |
| Black | 1.53 | 1.49 | 1.59 | 0.000 |  |  |
| Other | 1.38 | 1.33 | 1.43 | 0.000 |  |  |
| **Model 5a** | | | | | | |
| Unlikely migrant (English as first language without complex social factors) | Reference | Reference | Reference | Reference | 622,544 | 0.073 |
| Probable migrant (English not their first language and complex social factors) | 0.92 | 0.89 | 0.95 | 0.000 |  |  |
| Possible migrant (only English not their first language) | 1.01 | 0.99 | 1.03 | 0.458 |  |  |
| Possible migrant (only complex social factors) | 1.09 | 1.05 | 1.13 | 0.000 |  |  |
| Missing migration status | 0.26 | 0.25 | 0.26 | 0.000 |  |  |
| Not most deprived | Reference | Reference | Reference | Reference |  |  |
| Most deprived | 1.30 | 1.28 | 1.32 | 0.000 |  |  |
| Mother's age at booking (years) | 1.00 | 1.00 | 1.00 | 0.001 |  |  |
| White | Reference | Reference | Reference | Reference |  |  |
| Mixed | 1.56 | 1.49 | 1.63 | 0.000 |  |  |
| Asian | 2.86 | 2.80 | 2.91 | 0.000 |  |  |
| Black | 1.41 | 1.37 | 1.45 | 0.000 |  |  |
| Other | 1.32 | 1.27 | 1.37 | 0.000 |  |  |
| Missing ethnicity | 0.99 | 0.97 | 1.01 | 0.325 |  |  |

| **Outcome: booking after 10 weeks gestations** | | | | | | |
| --- | --- | --- | --- | --- | --- | --- |
| **Migration status** | **Odds ratio** | **95% LCI** | **95% UCI** | **P value** | **Number of observations** | **Pseudo R squared (Hosmer-Lemeshow)** |
| **Model 1** | | | | | | |
| Unlikely migrant (English as first language without complex social factors) | Reference | Reference | Reference | Reference | 652,309 | 0.008 |
| Probable migrant (English not their first language and complex social factors) | 2.66 | 2.59 | 2.73 | 0.000 |  |  |
| Possible migrant (only English not their first language) | 1.35 | 1.33 | 1.38 | 0.000 |  |  |
| Possible migrant (only complex social factors) | 1.44 | 1.41 | 1.47 | 0.000 |  |  |
| Missing migration status (first language or complex social factor data missing) | 1.19 | 1.18 | 1.21 | 0.000 |  |  |
| **Model 2b** | | | | | | |
| Unlikely migrant (English as first language without complex social factors) | Reference | Reference | Reference | Reference | 500,439 | 0.018 |
| Probable migrant (English not their first language and complex social factors) | 2.25 | 2.18 | 2.32 | 0.000 |  |  |
| Possible migrant (only English not their first language) | 1.22 | 1.20 | 1.24 | 0.000 |  |  |
| Possible migrant (only complex social factors) | 1.38 | 1.34 | 1.42 | 0.000 |  |  |
| Missing migration status (first language or complex social factor data missing) | 1.15 | 1.14 | 1.17 | 0.000 |  |  |
| Not most deprived | Reference | Reference | Reference | Reference |  |  |
| Most deprived | 1.07 | 1.06 | 1.09 | 0.000 |  |  |
| Mother's age at booking (years) | 1.00 | 1.00 | 1.00 | 0.000 |  |  |
| Number of previous live births | 1.14 | 1.13 | 1.14 | 0.000 |  |  |
| White | Reference | Reference | Reference | Reference |  |  |
| Mixed | 1.35 | 1.29 | 1.41 | 0.000 |  |  |
| Asian | 1.29 | 1.27 | 1.32 | 0.000 |  |  |
| Black | 1.99 | 1.94 | 2.05 | 0.000 |  |  |
| Other | 1.69 | 1.63 | 1.74 | 0.000 |  |  |
| Missing ethnicity | 1.18 | 1.16 | 1.20 | 0.000 |  |  |
| **Model 3b** | | | | | | |
| Unlikely migrant (English as first language without complex social factors) | Reference | Reference | Reference | Reference | 500,439 | 0.013 |
| Probable migrant (English not their first language and complex social factors) | 2.54 | 2.47 | 2.62 | 0.000 |  |  |
| Possible migrant (only English not their first language) | 1.33 | 1.31 | 1.35 | 0.000 |  |  |
| Possible migrant (only complex social factors) | 1.39 | 1.35 | 1.44 | 0.000 |  |  |
| Missing migration status (first language or complex social factor data missing) | 1.16 | 1.15 | 1.18 | 0.000 |  |  |
| Not most deprived | Reference | Reference | Reference | Reference |  |  |
| Most deprived | 1.12 | 1.10 | 1.14 | 0.000 |  |  |
| Mother's age at booking (years) | 1.01 | 1.00 | 1.01 | 0.000 |  |  |
| Number of previous live births | 1.14 | 1.13 | 1.14 | 0.000 |  |  |
| **Model 4b** | | | | | | |
| Unlikely migrant (English as first language without complex social factors) | Reference | Reference | Reference | Reference | 322,673 | 0.02 |
| Probable migrant (English not their first language and complex social factors) | 2.27 | 2.20 | 2.35 | 0.000 |  |  |
| Possible migrant (only English not their first language) | 1.21 | 1.18 | 1.23 | 0.000 |  |  |
| Possible migrant (only complex social factors) | 1.37 | 1.33 | 1.42 | 0.000 |  |  |
| Not most deprived | Reference | Reference | Reference | Reference |  |  |
| Most deprived | 1.00 | 0.98 | 1.02 | 0.801 |  |  |
| Mother's age at booking (years) | 1.00 | 1.00 | 1.01 | 0.000 |  |  |
| Number of previous live births | 1.13 | 1.12 | 1.13 | 0.000 |  |  |
| White | Reference | Reference | Reference | Reference |  |  |
| Mixed | 1.28 | 1.22 | 1.35 | 0.000 |  |  |
| Asian | 1.27 | 1.24 | 1.30 | 0.000 |  |  |
| Black | 1.92 | 1.86 | 1.98 | 0.000 |  |  |
| Other | 1.68 | 1.62 | 1.74 | 0.000 |  |  |
| **Model 5b** |  |  |  |  |  |  |
| Unlikely migrant (English as first language without complex social factors) | Reference | Reference | Reference | Reference | 500,439 | 0.018 |
| Probable migrant (English not their first language and complex social factors) | 2.25 | 2.18 | 2.32 | 0.000 |  |  |
| Possible migrant (only English not their first language) | 1.22 | 1.20 | 1.24 | 0.000 |  |  |
| Possible migrant (only complex social factors) | 1.38 | 1.34 | 1.42 | 0.000 |  |  |
| Missing migration status (first language or complex social factor data missing) | 1.15 | 1.14 | 1.17 | 0.000 |  |  |
| Not most deprived | Reference | Reference | Reference | Reference |  |  |
| Most deprived | 1.07 | 1.06 | 1.09 | 0.000 |  |  |
| Mother's age at booking (years) | 1.00 | 1.00 | 1.00 | 0.000 |  |  |
| Number of previous live births | 1.14 | 1.13 | 1.14 | 0.000 |  |  |
| White | Reference | Reference | Reference | Reference |  |  |
| Mixed | 1.35 | 1.29 | 1.41 | 0.000 |  |  |
| Asian | 1.29 | 1.27 | 1.32 | 0.000 |  |  |
| Black | 1.99 | 1.94 | 2.05 | 0.000 |  |  |
| Other | 1.69 | 1.63 | 1.74 | 0.000 |  |  |
| Missing ethnicity | 1.18 | 1.16 | 1.20 | 0.000 |  |  |

| **Outcome: booking after 16 weeks gestation** | | | | | | |
| --- | --- | --- | --- | --- | --- | --- |
| **Migration status** | **Odds ratio** | **95% LCI** | **95% UCI** | **P value** | **Number of observations** | **Pseudo R squared (Hosmer-Lemeshow)** |
| **Model 1** | | | | | | |
| Unlikely migrant (English as first language without complex social factors) | Reference | Reference | Reference | Reference | 652,309 | 0.026 |
| Probable migrant (English not their first language and complex social factors) | 5.04 | 4.88 | 5.22 | 0.000 |  |  |
| Possible migrant (only English not their first language) | 1.62 | 1.57 | 1.67 | 0.000 |  |  |
| Possible migrant (only complex social factors) | 2.43 | 2.35 | 2.52 | 0.000 |  |  |
| Missing migration status (first language or complex social factor data missing) | 1.92 | 1.88 | 1.96 | 0.000 |  |  |
| **Model 2b** | | | | | | |
| Unlikely migrant (English as first language without complex social factors) | Reference | Reference | Reference | Reference | 500,439 | 0.042 |
| Probable migrant (English not their first language and complex social factors) | 3.69 | 3.54 | 3.83 | 0.000 |  |  |
| Possible migrant (only English not their first language) | 1.43 | 1.38 | 1.48 | 0.000 |  |  |
| Possible migrant (only complex social factors) | 2.23 | 2.12 | 2.34 | 0.000 |  |  |
| Missing migration status (first language or complex social factor data missing) | 1.65 | 1.61 | 1.70 | 0.000 |  |  |
| Not most deprived | Reference | Reference | Reference | Reference |  |  |
| Most deprived | 1.09 | 1.06 | 1.13 | 0.000 |  |  |
| Mother's age at booking (years) | 0.97 | 0.97 | 0.97 | 0.000 |  |  |
| Number of previous live births | 1.19 | 1.18 | 1.20 | 0.000 |  |  |
| White | Reference | Reference | Reference | Reference |  |  |
| Mixed | 1.54 | 1.43 | 1.66 | 0.000 |  |  |
| Asian | 1.19 | 1.15 | 1.23 | 0.000 |  |  |
| Black | 2.28 | 2.18 | 2.37 | 0.000 |  |  |
| Other | 2.05 | 1.95 | 2.15 | 0.000 |  |  |
| Missing ethnicity | 1.71 | 1.66 | 1.76 | 0.000 |  |  |
| **Model 3b** | | | | | | |
| Unlikely migrant (English as first language without complex social factors) | Reference | Reference | Reference | Reference | 500,439 | 0.032 |
| Probable migrant (English not their first language and complex social factors) | 4.39 | 4.22 | 4.55 | 0.000 |  |  |
| Possible migrant (only English not their first language) | 1.61 | 1.55 | 1.66 | 0.000 |  |  |
| Possible migrant (only complex social factors) | 2.25 | 2.14 | 2.36 | 0.000 |  |  |
| Missing migration status (first language or complex social factor data missing) | 1.70 | 1.66 | 1.74 | 0.000 |  |  |
| Not most deprived | Reference | Reference | Reference | Reference |  |  |
| Most deprived | 1.16 | 1.13 | 1.19 | 0.000 |  |  |
| Mother's age at booking (years) | 0.98 | 0.97 | 0.98 | 0.000 |  |  |
| Number of previous live births | 1.18 | 1.17 | 1.19 | 0.000 |  |  |
| **Model 4b** | | | | | | |
| Unlikely migrant (English as first language without complex social factors) | Reference | Reference | Reference | Reference | 322,673 | 0.049 |
| Probable migrant (English not their first language and complex social factors) | 3.85 | 3.68 | 4.03 | 0.000 |  |  |
| Possible migrant (only English not their first language) | 1.47 | 1.42 | 1.53 | 0.000 |  |  |
| Possible migrant (only complex social factors) | 2.21 | 2.09 | 2.32 | 0.000 |  |  |
| Not most deprived | Reference | Reference | Reference | Reference |  |  |
| Most deprived | 1.02 | 0.98 | 1.06 | 0.433 |  |  |
| Mother's age at booking (years) | 0.97 | 0.97 | 0.97 | 0.000 |  |  |
| Number of previous live births | 1.19 | 1.17 | 1.20 | 0.000 |  |  |
| White | Reference | Reference | Reference | Reference |  |  |
| Mixed | 1.55 | 1.42 | 1.69 | 0.000 |  |  |
| Asian | 1.14 | 1.10 | 1.19 | 0.000 |  |  |
| Black | 2.31 | 2.20 | 2.43 | 0.000 |  |  |
| Other | 1.94 | 1.84 | 2.06 | 0.000 |  |  |
| **Model 5b** | | | | | | |
| Unlikely migrant (English as first language without complex social factors) | Reference | Reference | Reference | Reference | 500,439 | 0.042 |
| Probable migrant (English not their first language and complex social factors) | 3.69 | 3.54 | 3.83 | 0.000 |  |  |
| Possible migrant (only English not their first language) | 1.43 | 1.38 | 1.48 | 0.000 |  |  |
| Possible migrant (only complex social factors) | 2.23 | 2.12 | 2.34 | 0.000 |  |  |
| Missing migration status (first language or complex social factor data missing) | 1.65 | 1.61 | 1.70 | 0.000 |  |  |
| Not most deprived | Reference | Reference | Reference | Reference |  |  |
| Most deprived | 1.09 | 1.06 | 1.13 | 0.000 |  |  |
| Mother's age at booking (years) | 0.97 | 0.97 | 0.97 | 0.000 |  |  |
| Number of previous live births | 1.19 | 1.18 | 1.20 | 0.000 |  |  |
| White | Reference | Reference | Reference | Reference |  |  |
| Mixed | 1.54 | 1.43 | 1.66 | 0.000 |  |  |
| Asian | 1.19 | 1.15 | 1.23 | 0.000 |  |  |
| Black | 2.28 | 2.18 | 2.37 | 0.000 |  |  |
| Other | 2.05 | 1.95 | 2.15 | 0.000 |  |  |
| Missing ethnicity | 1.71 | 1.66 | 1.76 | 0.000 |  |  |

| **Outcome: booking after 20 weeks gestation** | | | | | | |
| --- | --- | --- | --- | --- | --- | --- |
| **Migration status** | **Odds ratio** | **95% LCI** | **95% UCI** | **P value** | **Number of observations** | **Pseudo R squared (Hosmer-Lemeshow)** |
| **Model 1** | | | | | | |
| Unlikely migrant (English as first language without complex social factors) | Reference | Reference | Reference | Reference | 652,309 | 0.025 |
| Probable migrant (English not their first language and complex social factors) | 4.69 | 4.50 | 4.88 | 0.000 |  |  |
| Possible migrant (only English not their first language) | 1.45 | 1.39 | 1.51 | 0.000 |  |  |
| Possible migrant (only complex social factors) | 2.39 | 2.29 | 2.49 | 0.000 |  |  |
| Missing migration status (first language or complex social factor data missing) | 2.17 | 2.11 | 2.22 | 0.000 |  |  |
| **Model 2b** | | | | | | |
| Unlikely migrant (English as first language without complex social factors) | Reference | Reference | Reference | Reference | 524,215 | 0.038 |
| Probable migrant (English not their first language and complex social factors) | 3.69 | 3.53 | 3.86 | 0.000 |  |  |
| Possible migrant (only English not their first language) | 1.30 | 1.24 | 1.35 | 0.000 |  |  |
| Possible migrant (only complex social factors) | 2.13 | 2.03 | 2.23 | 0.000 |  |  |
| Missing migration status (first language or complex social factor data missing) | 1.84 | 1.78 | 1.89 | 0.000 |  |  |
| Not most deprived | Reference | Reference | Reference | Reference |  |  |
| Most deprived | 1.03 | 1.00 | 1.07 | 0.057 |  |  |
| Mother's age at booking (years) | 0.98 | 0.97 | 0.98 | 0.000 |  |  |
| Number of previous live births | 1.11 | 1.10 | 1.12 | 0.000 |  |  |
| White | Reference | Reference | Reference | Reference |  |  |
| Mixed | 1.45 | 1.33 | 1.58 | 0.000 |  |  |
| Asian | 1.13 | 1.08 | 1.18 | 0.000 |  |  |
| Black | 2.10 | 2.00 | 2.20 | 0.000 |  |  |
| Other | 2.04 | 1.93 | 2.15 | 0.000 |  |  |
| Missing ethnicity | 1.81 | 1.75 | 1.87 | 0.000 |  |  |
| **Model 3b** | | | | | | |
| Unlikely migrant (English as first language without complex social factors) | Reference | Reference | Reference | Reference | 524,215 | 0.028 |
| Probable migrant (English not their first language and complex social factors) | 4.36 | 4.18 | 4.55 | 0.000 |  |  |
| Possible migrant (only English not their first language) | 1.44 | 1.39 | 1.51 | 0.000 |  |  |
| Possible migrant (only complex social factors) | 2.14 | 2.04 | 2.24 | 0.000 |  |  |
| Missing migration status (first language or complex social factor data missing) | 1.89 | 1.83 | 1.95 | 0.000 |  |  |
| Not most deprived | Reference | Reference | Reference | Reference |  |  |
| Most deprived | 1.09 | 1.05 | 1.13 | 0.000 |  |  |
| Mother's age at booking (years) | 0.98 | 0.98 | 0.98 | 0.000 |  |  |
| Number of previous live births | 1.10 | 1.09 | 1.11 | 0.000 |  |  |
| **Model 4b** | | | | | | |
| Unlikely migrant (English as first language without complex social factors) | Reference | Reference | Reference | Reference | 337,622 | 0.042 |
| Probable migrant (English not their first language and complex social factors) | 3.92 | 3.73 | 4.13 | 0.000 |  |  |
| Possible migrant (only English not their first language) | 1.33 | 1.26 | 1.39 | 0.000 |  |  |
| Possible migrant (only complex social factors) | 2.15 | 2.04 | 2.27 | 0.000 |  |  |
| Not most deprived | Reference | Reference | Reference | Reference |  |  |
| Most deprived | 0.96 | 0.92 | 1.00 | 0.076 |  |  |
| Mother's age at booking (years) | 0.98 | 0.97 | 0.98 | 0.000 |  |  |
| Number of previous live births | 1.11 | 1.09 | 1.12 | 0.000 |  |  |
| White | Reference | Reference | Reference | Reference |  |  |
| Mixed | 1.51 | 1.37 | 1.67 | 0.000 |  |  |
| Asian | 1.08 | 1.03 | 1.14 | 0.002 |  |  |
| Black | 2.14 | 2.02 | 2.26 | 0.000 |  |  |
| Other | 1.96 | 1.84 | 2.09 | 0.000 |  |  |
| **Model 5b** | | | | | | |
| Unlikely migrant (English as first language without complex social factors) | Reference | Reference | Reference | Reference | 500,439 | 0.035 |
| Probable migrant (English not their first language and complex social factors) | 3.52 | 3.36 | 3.69 | 0.000 |  |  |
| Possible migrant (only English not their first language) | 1.29 | 1.24 | 1.35 | 0.000 |  |  |
| Possible migrant (only complex social factors) | 2.36 | 2.23 | 2.50 | 0.000 |  |  |
| Missing migration status (first language or complex social factor data missing) | 1.82 | 1.77 | 1.88 | 0.000 |  |  |
| Not most deprived | Reference | Reference | Reference | Reference |  |  |
| Most deprived | 1.03 | 0.99 | 1.07 | 0.101 |  |  |
| Mother's age at booking (years) | 0.98 | 0.97 | 0.98 | 0.000 |  |  |
| Number of previous live births | 1.10 | 1.09 | 1.11 | 0.000 |  |  |
| White | Reference | Reference | Reference | Reference |  |  |
| Mixed | 1.51 | 1.38 | 1.65 | 0.000 |  |  |
| Asian | 1.16 | 1.11 | 1.21 | 0.000 |  |  |
| Black | 2.13 | 2.02 | 2.24 | 0.000 |  |  |
| Other | 2.06 | 1.94 | 2.18 | 0.000 |  |  |
| Missing ethnicity | 1.84 | 1.77 | 1.90 | 0.000 |  |  |
